# Epigenetic Exposure Signatures Distinguish Early-Onset from Later-Onset Colorectal Cancer in a Racially Diverse US Cohort: Findings from the Disparities and Cancer Epidemiology (DANCE) Study

**DOI:** 10.64898/2026.09.28.26363311

**Authors:** Sarah M Lima, Jaeil Ahn, Chiranjeev Dash, Mei-Chin Hseih, Rena R Jones, Batsirai Mabvakure, Reem Muhsen, Siddhi Patil, Alina Peluso, Candy Promprasert, Tingting Qin, Julie J Ruterbusch, Maureen A Sartor, Ann G Schwartz, Angela S Wenzlaff, Rui Zhang, Elena M Stoffel, Kristen S Purrington, Laura S Rozek

## Abstract

**Background:** The incidence of early-onset colorectal cancer (EOCRC), defined as CRC diagnosed age <50, is rising in the United States and is the leading cause of cancer death in among those with early-onset cancer. Epigenetic exposure signatures based on methylation risk scores (MRS) offer a novel approach to identify long-term exposures that may contribute to EOCRC risk. We sought to replicate findings from a recent study in a racially and geographically diverse CRC cohort.

**Methods:** We computed weighted MRS across lifestyle, air pollution, and pesticide exposome traits in 146 participants from the Disparities and Cancer Epidemiology (DANCE) cohort, a diverse population-based CRC study, applying the Maas *et al*. (2026) framework to tumor tissue. Associations between MRS and age at CRC onset (EOCRC, n=29; intermediate-onset (IOCRC) 50–64 years, n=69; late-onset, ≥65 years, n=48) were evaluated using linear and polytomous regression, with adjustment and stratification for race, geography, Area Deprivation Index (ADI), and Multi-Environmental Exposure Index (MEEI). Spearman correlations were calculated between MRSes and observed exposures.

**Results:** EOCRC cases were predominately Black (65.5% vs. 31.3% among LOCRC) and resided in higher-deprivation neighborhoods. We found higher odds of EOCRC associated with epigenetic exposure signatures for 2,4-D herbicide, heptachlor, coarse particulate matter (PM), Mediterranean Diet Score, and smoking, and borderline associations for fine PM and nitrogen dioxide. IOCRC was associated with signatures for 2,4-D, lindane, and obesity. Heterogeneity was observed by race and area-level factors, though associations were largely consistent across strata. ADI and MEEI were correlated with multiple pesticide and pollutant epigenetic signatures.

**Conclusions:** In a diverse US cohort, epigenetic exposure signatures distinguish CRC cohorts, particularly EOCRC from later-onset disease, with associations consistent with but not identical to previous research. EOCRC in DANCE was characterized by higher epigenetic exposure signatures for air pollution, smoking, and distinct pesticide exposure profiles. IOCRC had overlapping characteristics with early- and late-onset and were the only group with a significant obesity signature. These findings support the hypothesis that exposome-driven epigenetic changes contribute to EOCRC risk and highlight the importance of studying diverse populations to characterize the full spectrum of environmental risk factors.

**Graphical Abstract:** Summary results for associations between epigenetic exposure signatures and age of colorectal cancer onset in the DANCE cohort.

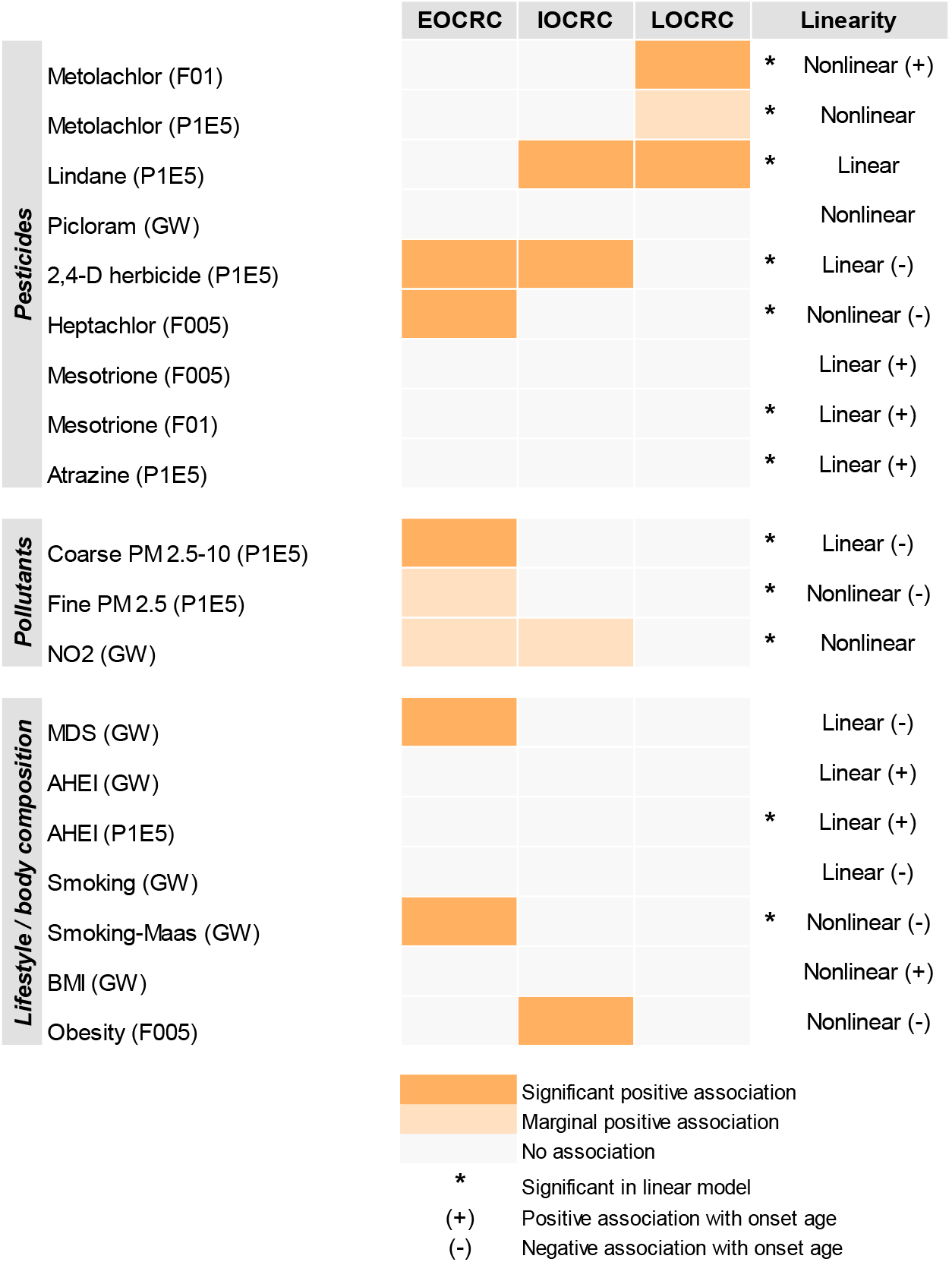

## Introduction

Colorectal cancer (CRC) has historically been considered a disease of older adults, but rates of early-onset CRC (EOCRC), defined as diagnosis before age 50, have increased by approximately 2% annually since the mid-1990s,^1,2^ leading the United States Preventive Services Task Force to lower its recommended screening age to 45.^3^ Notably, birth cohort analyses document a clear generational pattern: individuals born after 1960 face substantially higher CRC risk at any given age than preceding generations, climbing with each subsequent birth cohort.^2,4^ A generational shift of this magnitude, occurring over just a few decades, is not consistent with a genetic explanation, as germline susceptibility does not change fast enough to account for it. Birth cohort effects appear after the introduction or unprecedented change of a population-wide exposure. Consequently, environmental exposures that have undergone changes over the 20^th^ century may contribute to rises in EOCRC observed today.^5^ Moreover, persistent racial disparities in CRC further indicate environmental sources, as minoritized groups tend to have higher adverse exposure.^6^

Despite early-life exposures being repeatedly proposed as central to EOCRC, direct longitudinal evidence remains thin and no single exposure or mechanism has been established. The exposome, or the totality of exposures across a lifetime, has emerged as a critical framework for understanding EOCRC etiology,^7^ as the search for a unifying genetic or heritable cause has not explained this trend. Epigenetic exposure signatures have been proposed as a method to quantify exposomic effects across the lifecourse, acting as long-term biomarkers despite not being derived from longitudinal data.^8,9^ These epigenetic signatures provide a unique opportunity to efficiently study potential drivers of etiology, a significant benefit for CRC studies given its long latency period. In a groundbreaking study, Maas *et al*. validated DNA methylation risk scores (MRS) in a multi-cohort epigenome-wide study of EOCRC, identifying picloram, an herbicide, as a novel EOCRC risk factor.^10^ This finding is illustrative of the broader point; the responsible exposures are still largely unidentified but preliminary evidence indicates environmental rather than inherited causes. While groundbreaking, the Maas *et al*. study was conducted across predominantly white CRC cohorts without corresponding individual- or area-level exposure data. Given the racial, socioeconomic, and geographic disparities in (EO)CRC, it is critical to understand whether these epigenetic exposure signatures generalize to diverse US populations and whether different exposures emerge.

The Disparities and Cancer Epidemiology (DANCE) cohort is a rigorously characterized, population-based CRC study that is uniquely positioned to address this gap. DANCE is enriched for Black CRC cases (~46% of participants) and includes extensive epidemiologic, clinical, geographic, and socioeconomic characterization. Participants are recruited from metro-Detroit, Michigan and Louisiana, two areas in the US with significant socio-environmental burdens but varying urbanicity/rurality. In this study, we applied the Maas *et al*. epigenetic exposure signature framework to a DANCE pilot subset to characterize the landscape of epigenetic exposure signatures and evaluate relationships with age of CRC onset. We further expanded upon Maas *et al*.’s study to assess whether race, geography, area deprivation, and environmental exposures impact associations between epigenetic exposure signatures and CRC onset, and to examine whether epigenetic exposure signatures are correlated with measured exposures.

## Methods

### Study Design and Population

The DANCE cohort is a population-based CRC cohort that enrolled incident CRC cases from metro-Detroit, Michigan and Louisiana. The present case-case analysis used a pilot subset of 146 participants with available tumor DNA methylation data. Participants were classified into three onset (i.e., age-at-diagnosis) groups: EOCRC (<50 years; n=29), intermediate-onset CRC (IOCRC; 50–64 years; n=69), and late-onset CRC (LOCRC; ≥65 years; n=48). Race was self-reported and categorized as non-Hispanic Black or non-Hispanic White (hereon referred to as Black and White). All DANCE participants were non-Hispanic. Informed consent was obtained from all participants and IRB approval was obtained at participating sites.

### DNA Methylation and MRS Computation

Genome-wide DNA methylation was profiled using the Illumina Infinium MethylationEPIC BeadChip array from DNA extracted from formalin-fixed paraffin-embedded (FFPE) tumor tissue. Beta-values were processed using the sesame R package and adjusted for Horvath epigenetic age. Following the approach of Maas et al. (2026),^10^ weighted MRS were computed across exposome traits encompassing lifestyle factors,^11-13^ body size,^14,15^ air pollution measures,^16,17^ and pesticides.^18^ Effect sizes from relevant epigenome-wide association studies were applied to tumor methylation levels at corresponding CpG sites, across five marker selection thresholds (genome-wide [GW]: P<1.2×10^−7^; P1E5: P<1.0×10^−5^; FDR thresholds F01, F005, F001). MRS values were standardized to a mean of 0 and standard deviation (SD) of 1 across all DANCE participants.

### Reported & Measured Exposures

Baseline questionnaires collected self-reported information on current dietary factors, smoking habits, and body size. Dietary information included regular intake of fruits, vegetables, red meat, processed meat, fast food, and soda; answers were 6-level frequency selections, ranging from never to frequent (i.e., 5+ servings/day or >once/day). We created a diet score consistent with American Cancer Society (ACS) dietary guidelines by converting dietary intake data to quartiles and assigning points based on quartile.^19^ A higher diet score represents higher adherence to ACS dietary guidelines. Smoking variables included smoking status (never, former, current) and e-cigarette use (ever, never). We calculated current and pre-diagnosis body mass index (BMI; kg/m^2^). Neighborhood-level socioeconomic deprivation was quantified using the Area Deprivation Index (ADI) national rank at the census block group level based on residential address at diagnosis; ADI ranges 0 to 100, with higher score indicating greater socioeconomic deprivation.^20^ The Multi-Exposure Environmental Index (MEEI) is a measure of aggregate environmental exposures measured at the census tract-level.^21^ MEEI includes both beneficial and adverse environmental exposures and ranges −2 to 7; a negative number indicates beneficial environment, a higher positive number indicates multiple adverse exposures. We also included the individual components of MEEI, also measured at the census tract-level: percent greenspace and open areas, ultra-violet (UV) radiation, ozone, fine particulate matter (PM_2.5_, <2.5 µm in diameter), proportion of nighttime population within 2km of Toxics Release Inventory facility (variable hereon referred to as TRI), cold waves, heat waves, and heavy precipitation. The time frames for these exposures vary, but generally aggregated data from 2010-2020; more details on MEEI and its components has been published elsewhere.^21^ We used the Climate Vulnerability Index (CVI) national ranking of agricultural pesticide use at the census tract-level as a surrogate for pesticide exposure.^22^

### Statistical Analysis

Descriptive statistics were calculated for the sample according to CRC onset. Chi-square, Fisher’s exact (instances of small cell count), and analysis of variance (ANOVA) tests estimated statistical differences by CRC onset. We estimated group means of epigenetic exposure signatures according to age of onset and used ANOVAs to test for statistically significant differences.

We tested whether epigenetic exposure signatures were associated with age of CRC onset. Linear regression models estimated associations between SD-increase in MRS and difference in years for age of CRC onset (*β* coefficients, 95% confidence intervals, CIs). We evaluated crude associations and investigated race and ADI as separate covariates. We included sex as a potential covariate, but results were unchanged compared to crude models and thus were not included in reported results. Crude linear models were additionally stratified by race, ADI, state, and MEEI to assess for heterogeneity. We used polytomous regression to test whether MRS is associated with CRC onset category (early, intermediate, late). Polytomous models estimated odds ratios (ORs) and 95% CIs of EOCRC and IOCRC associated with SD-increase in MRS compared to LOCRC, the referent outcome. Polytomous models were run crudely and with separate adjustment for race and ADI. As a sensitivity analysis, we used generalized additive models (GAMs) to capture nonlinear associations between MRS and age of onset (continuous). Due to multiple comparisons, we used Benjamini-Hochberg correction for 10% and 20% false discovery rates (FDRs) for the main linear and polytomous regression models.

Spearman correlations were used to assess the relationship between epigenetic exposure signatures and observed individual- and area-level exposures. Correlational analyses were split by environmental vs lifestyle/body composition domains. Statistical analyses were conducted using SAS 9.4 (Cary, NC); DNA methylation processing, MRS computation, and figure generation were performed using R version 4.5.3.

## Results

The DANCE pilot subset included 146 CRC cases; 29 (19.9%) were early-onset, 69 (47.3%) were intermediate-onset, and 48 (32.9%) were late-onset (Table 1). Over 90% of participants were from metro-Detroit and 46% were Black. EOCRC cases were significantly more likely to be Black, never smokers, ever vaped, diagnosed at distant stage, and live in higher deprivation neighborhoods at diagnosis. Roughly 80% of participants were overweight or obese, but there were no significant differences by age of onset, although EOCRC cases had the highest obesity prevalence. We did not detect a significant difference in MEEI according to onset, though data indicate directionality, with EOCRC cases having the highest environmental burden and LOCRC having the lowest.

**Table 1.** Descriptive statistics by onset.

| **Characteristic** | **Total**<br>**N=146** | **CRC Onset** |  |  | **P-value** |
| --- | --- | --- | --- | --- | --- |
|  |  | **Early**<br>**29 (19.9%)** | **Intermediate**<br>**69 (47.3%)** | **Late**<br>**48 (32.9%)** |  |
| Sex |  |  |  |  | 0.837 |
| Female | 74 (50.7%) | 14 (48.3%) | 34 (49.3%) | 26 (54.2%) |  |
| Male | 72 (49.3%) | 15 (51.7%) | 35 (50.7%) | 22 (45.8%) |  |
| Race |  |  |  |  | **0.013** |
| Black | 67 (45.9%) | 19 (65.5%) | 33 (47.8%) | 15 (31.3%) |  |
| White | 79 (54.1%) | 10 (34.5%) | 36 (52.2%) | 33 (68.8%) |  |
| State |  |  |  |  | 0.800 |
| Louisiana | 13 (8.9%) | 3 (10.3%) | 5 (7.2%) | 5 (10.4%) |  |
| Michigan | 133 (91.1%) | 26 (89.7%) | 64 (92.8%) | 43 (89.6%) |  |
| Income |  |  |  |  | 0.292 |
| <\$20k | 33 (22.6%) | 7 (24.1%) | 20 (29.0%) | 6 (12.5%) | |
| \$20k - <\$60k | 37 (25.3%) | 8 (27.6%) | 13 (18.8%) | 16 (33.3%) | |
| ≥\$60k | 70 (48.0%) | 12 (41.4%) | 33 (47.8%) | 25 (52.1%) | |
| Unknown | 6 (4.1%) | 2 (6.9%) | 3 (4.3%) | 1 (2.1%) |  |
| BMI |  |  |  |  | 0.352 |
| Normal | 30 (20.8%) | 4 (13.8%) | 14 (20.9%) | 12 (25.0%) |  |
| Overweight | 49 (34.0%) | 7 (24.1%) | 23 (34.3%) | 19 (39.6%) |  |
| Obese | 64 (44.4%) | 18 (62.1%) | 29 (43.3%) | 17 (35.4%) |  |
| Smoking status |  |  |  |  | **0.006** |
| Never | 68 (60.7%) | 20 (83.3%) | 34 (64.2%) | 14 (40.0%) |  |
| Former | 36 (32.1%) | 3 (12.5%) | 14 (26.4%) | 19 (54.3%) |  |
| Current | 8 (7.4%) | 1 (4.2%) | 5 (9.4%) | 2 (5.7%) |  |
| Vaping status |  |  |  |  | **0.034** |
| Never | 9 (69.2%) | 34 (85.0%) | 32 (94.1%) | 14 (40.0%) |  |
| Ever | 4 (30.8%) | 6 (15.0%) | 1 (2.9%) | 19 (54.3%) |  |
| ACS diet score | 2.10 (0.9) | 1.97 (0.8) | 2.10 (0.9) | 2.20 (0.9) | 0.645 |
| ADI | 67.91 (29.3) | 79.96 (22.3) | 70.08 (29.7) | 57.22 (29.4) | **0.003** |
| MEEI | 2.28 (1.2) | 2.41 (0.9) | 2.34 (1.3) | 2.11 (1.3) | 0.473 |
| Stage |  |  |  |  | **0.006** |
| Local | 45 (30.8%) | 6 (20.7%) | 21 (30.4%) | 18 (37.5%) |  |
| Regional | 79 (54.1%) | 14 (48.3%) | 35 (50.7%) | 30 (62.5%) |  |
| Distant | 21 (14.4%) | 8 (27.6%) | 13 (18.8%) | 0 (0.0%) |  |
| Unknown | 1 (0.7%) | 1 (3.4%) | 0 (0.0%) | 0 (0.0%) |  |
| Tumor subsite |  |  |  |  | 0.304 |
| Left Colon | 54 (37.0%) | 14 (48.3%) | 26 (37.7%) | 14 (29.2%) |  |
| Rectum | 20 (13.7%) | 5 (17.2%) | 11 (15.9%) | 4 (8.3%) |  |
| Right Colon | 67 (45.9%) | 10 (34.5%) | 29 (42.0%) | 28 (58.3%) |  |
| Other | 5 (3.4%) | 0 (0.0%) | 3 (4.3%) | 2 (4.2%) |  |
| EOCRC = age <50, IOCRC = age 50-64, LOCRC = age 65+. ACS diet score = adherence to ACS dietary guidelines, range 0-4, higher score indicating more adherent (nutritious) diet. ADI = area deprivation index, range 0 (less deprivation) to 100 (more deprivation). MEEI = multi-exposure environmental index, range -2 (beneficial environment) to 7 (adverse environment). Fisher exact test used to estimate p-value for state, BMI, smoking status, vaping status, and tumor subsite. |  |  |  |  |  |

The distributions and group means of select MRSes are shown according to early-onset vs late-onset in Fig. 1. EOCRC cases had significantly higher scores for 2,4-D herbicide (P1E5), heptachlor (F005), coarse PM (PM_2.5-10_; P1E5), smoking-Maas (GW), and obesity (F005) than LOCRC cases; NO_2_ (GW) was marginally higher among EOCRC. Scores for metolachlor (F01) and lindane (P1E5) were higher among LOCRC cases than EOCRC. We did not detect significant differences in picloram (GW) by onset. MRSes not included in the figure did not show significant differences by onset.

**Figure 1.**
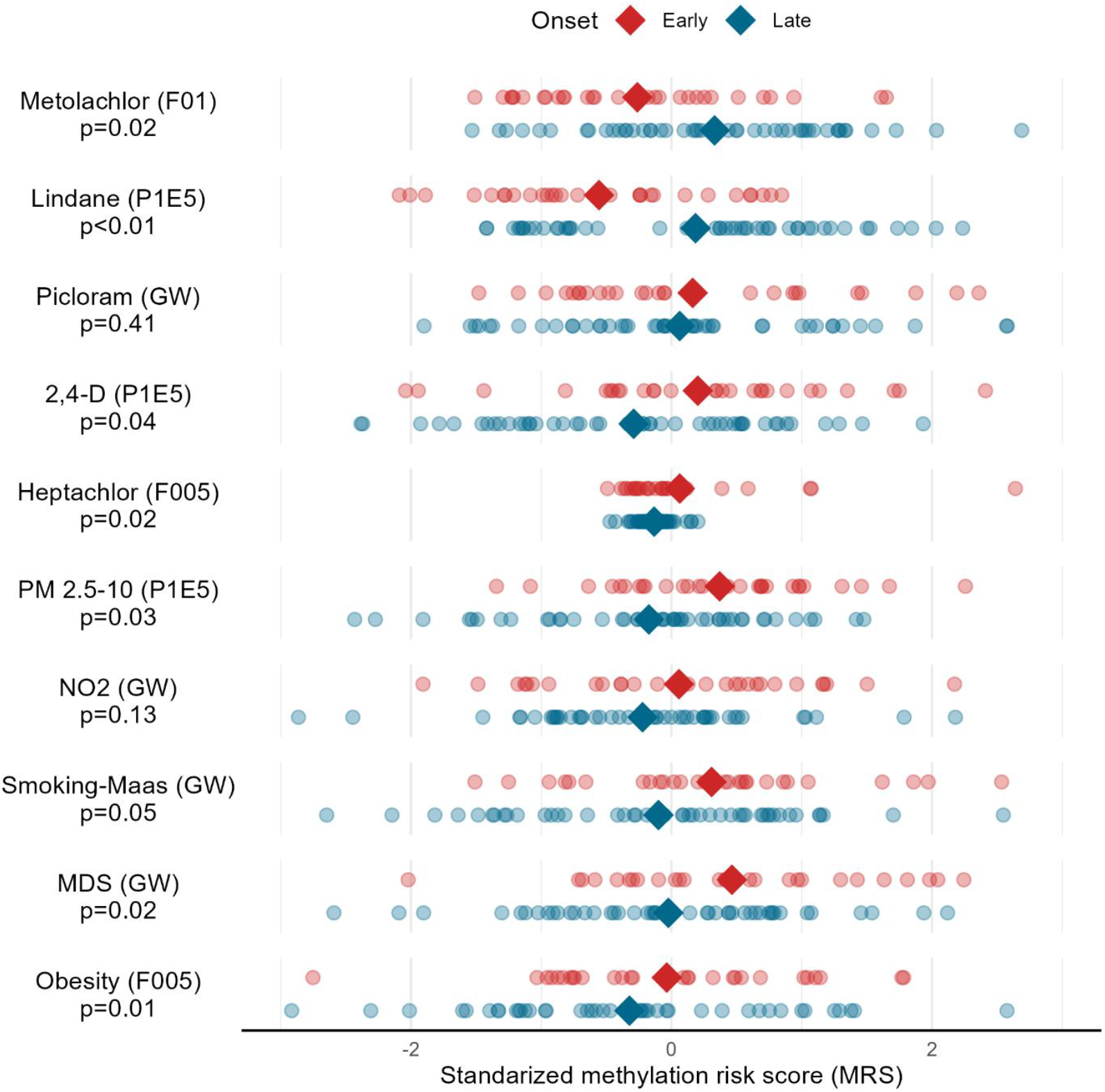
Mean and distribution of select epigenetic exposure signatures according to early vs late onset. Diamonds indicate group means. P-values estimated from ANOVA tests for differences in mean by early, intermediate, late onset; only early-onset vs late-onset distributions shown. Early-onset (age<50) n=29, late-onset (age 65+) n=48. MDS: Mediterranean Diet Score. Positive MRS (right side) suggests higher long-term exposure relative to the sample; negative MRS (left side) suggests lower longer-term exposure relative to the sample.

Table 2 reports associations between epigenetic exposure signatures and difference in years for age of CRC onset. Among pesticide-related signatures, we found a significantly younger age of onset associated with 2,4-D herbicide and heptachlor, with estimated effects of 2.47 (95% CI: −4.17, −0.76) and 2.13 (95% CI: −3.85, −0.41) years younger onset per SD-increase in respective signatures. A SD-increase in metolachlor, lindane, mesotrione, and atrazine was associated with significantly older age of onset, associations ranging 1.74 (95% CI: 0.01, 3.47; mesotrione F01) to 3.08 (95% CI: 1.41, 4.76; lindane) years older. We did not detect an association between picloram MRS and age of onset. Signatures for PM_2.5-10_, PM_2.5_, and NO_2_ were all associated with significantly younger age of onset, ranging 1.77 (95% CI: −3.50, −0.04; PM2.5 P1E5) to 2.51 (95% CI: −4.21, −0.80; PM2.5-10 P1E5) years younger per SD-increase. Among lifestyle and body composition MRSes, a SD-increase in smoking-Maas was associated with significantly younger onset (1.76 years, 95% CI: −3.49, −0.03), while Alternative Healthy Eating Index (AHEI) MRS was associated with significantly older age of onset (2.00 years, 95% IC: 0.23, 3.68). Separate adjustment for race and ADI tended to attenuate associations, but MRSes with significant crude associations generally maintained significant or borderline associations, with the exception of metolachlor (P1E5), mesotrione (F01), and smoking-Maas.

**Table 2.** Difference in years for age of CRC onset associated with epigenetic exposure signatures (MRS).

|  |  |  |  |
| --- | --- | --- | --- |
| **MRS (per SD-change)** | **Crude model**<br>**$\beta$ (95% CI)** | **Race adjusted**<br>**$\beta$ (95% CI)** | **ADI adjusted**<br>**$\beta$ (95% CI)** |
| ***Pesticides*** |  |  |  |
| Metolachlor (F01) | **2.57 (0.87, 4.27)** | **1.84 (0.11, 3.56)** | **2.00 (0.25, 3.76)** |
| Metolachlor (P1E5) | **1.77 (0.05, 3.50)** | 1.06 (-0.66, 2.79) | 1.38 (-0.38, 3.13) |
| Lindane (P1E5) | **3.08 (1.41, 4.76)** | **2.81 (1.19, 4.44)** | **2.65 (0.96, 4.34)** |
| Picloram (GW) | 0.06 (-1.69, 1.81) | 0.10 (-1.59, 1.78) | 0.16 (-1.58, 1.90) |
| 2,4-D herbicide (P1E5) | **-2.47 (-4.17, -0.76)** | **-2.25 (-3.89, -0.61)** | **-1.83 (-3.59, -0.07)** |
| Heptachlor (F005) | **-2.13 (-3.85, -0.41)** | **-1.93 (-3.58, -0.27)** | **-1.91 (-3.59, -0.22)** |
| Mesotrione (F005) | 1.63 (-0.10, 3.36) | 1.65 (-0.01, 3.31) | 1.46 (-0.28, 3.19) |
| Mesotrione (F01) | **1.74 (0.01, 3.47)** | 1.64 (-0.02, 3.30) | 1.43 (-0.33, 3.19) |
| Atrazine (P1E5) | **1.96 (0.23, 3.68)** | **1.85 (0.19, 3.50)** | **2.02 (0.33, 3.71)** |
| ***Pollutants*** |  |  |  |
| Coarse PM<sub>2.5-10</sub> (P1E5) | **-2.51 (-4.21, -0.80)** | **-2.22 (-3.89, -0.57)** | **-2.23 (-3.98, -0.49)** |
| Fine PM<sub>2.5</sub> (P1E5) | **-1.77 (-3.50, -0.04)** | **-1.99 (-3.64, -0.33)** | **-1.86 (-3.54, -0.18)** |
| NO<sub>2</sub> (GW) | **-2.03 (-3.75, -0.31)** | **-1.76 (-3.42, -0.10)** | -1.77 (-3.55, 0.02) |
| ***Lifestyle / body composition*** |  |  |  |
| MDS (GW) | -1.45 (-3.19, 0.29) | -1.56 (-3.22, 0.11) | -1.33 (-3.03, 0.37) |
| AHEI (GW) | 1.31 (-0.42, 3.05) | 1.63 (-0.03, 3.30) | 1.13 (-0.60, 2.86) |
| AHEI (P1E5) | **2.00 (0.23, 3.68)** | **2.20 (0.56, 3.84)** | 1.70 (-0.01, 3.41) |
| Smoking (GW) | -0.98 (-2.73, 0.76) | -0.55 (-2.25, 1.14) | -0.84 (-2.59, 0.90) |
| Smoking-Maas (GW) | **-1.76 (-3.49, -0.03)** | -1.06 (-2.79, 0.66) | -1.48 (-3.24, 0.29) |
| BMI (GW) | 1.51 (-0.22, 3.25) | 0.91 (-0.80, 2.62) | 0.91 (-0.84, 2.66) |
| Obesity (F005) | -1.49 (-3.22, 0.25) | -1.34 (-3.01, 0.33) | -1.41 (-3.13, 0.31) |
| *Beta coefficients represent the difference in years for age of onset associated with SD-change in MRS. Positive coefficients = older, negative coefficients = younger. **Bold:** P<0.05. Race modeled as White (ref), Black; ADI modeled continuously. MDS = Mediterranean Diet Score, AHEI = Alternative Healthy Eating Index. MRS thresholds: GW = genome wide, F005 = 5% false discovery rate, F01 = 10% false discovery rate, P1E5 = p-value<10<sup>-5</sup>.* |  |  |  |

Table 3 shows odds of EOCRC and IOCRC compared to LOCRC associated with a SD-increase in MRS, assessing potential cohort effects. Multiple epigenetic signatures were associated with EOCRC, including 2,4-D, coarse PM, Mediterranean Diet Score (MDS), and smoking-Maas. A SD-increase in 2,4-D was associated with 67% higher odds of being early-onset (95% CI: 1.03, 2.72) and 53% higher odds of intermediate-onset (95% CI: 1.04, 2.25) compared to late-onset. A SD-increase in PM_2.5-10_ was associated with roughly a 2-fold increase in odds of early-onset compared to late-onset (OR=1.94, 95% CI: 1.16, 3.24), but was not associated with IOCRC (OR=1.29, 95% CI: 0.89, 1.88). Signatures for MDS and smoking-Maas were both associated with approximately 70% higher odds EOCRC (OR=1.73, 95% CI: 1.04 2.86; OR=1.71, 95% CI: 1.05, 2.79, respectively) but showed null associations with IOCRC. The obesity MRS was associated with intermediate-onset (OR=1.84, 95% CI: 1.22, 2.78) but not early-onset. Our results suggest metolachlor MRS (F01) is associated with LOCRC, as we found lower odds of both EOCRC (OR=0.53, 95% CI: 0.33, 0.87) and IOCRC (OR=0.62, 95% CI: 0.42, 0.92) per SD-increase. We found lower odds of EOCRC associated with lindane MRS (OR=0.45, 95% CI: 0.27, 0.75) but no difference in odds of IOCRC compared to LOCRC. These results were largely consistent after separate adjustment for race and ADI. Notably, associations between heptachlor MRS and EOCRC strengthened and became significant after adjustment for race and ADI. In the ADI model, a SD-increase in heptachlor was associated with 8.53-fold higher odds of EOCRC (95% CI: 1.35, 54.0) compared to LOCRC.

**Table 3.** Odds of early and intermediate CRC onset compared to late-onset CRC associated with epigenetic exposure signatures.

| MRS (per SD-change) | Crude model |  |  | Race adjusted |  |  | ADI adjusted |  |  |
| --- | --- | --- | --- | --- | --- | --- | --- | --- | --- |
|  | Early<br>N=29 | Intermediate<br>N=48 | Late<br>N=69 | Early<br>N=29 | Intermediate<br>N=48 | Late<br>N=69 | Early<br>N=29 | Intermediate<br>N=48 | Late<br>N=69 |
|  | OR (95% CI) | OR (95% CI) | Ref | OR (95% CI) | OR (95% CI) | Ref | OR (95% CI) | OR (95% CI) | Ref |
| **Pesticides** |  |  |  |  |  |  |  |  |  |
| Metolachlor (F01) | **0.53 (0.33, 0.87)** | **0.62 (0.42, 0.92)** | 1.00 | 0.62 (0.37, 1.04) | **0.66 (0.44, 0.99)** | 1.00 | 0.61 (0.36, 1.04) | **0.64 (0.42, 0.97)** | 1.00 |
| Metolachlor (P1E5) | 0.67 (0.42, 1.09) | **0.63 (0.42, 0.93)** | 1.00 | 0.79 (0.47, 1.31) | 0.67 (0.45, 1.01) | 1.00 | 0.76 (0.45, 1.27) | **0.65 (0.43, 0.98)** | 1.00 |
| Lindane (P1E5) | **0.45 (0.27, 0.75)** | 0.92 (0.63, 1.35) | 1.00 | **0.44 (0.26, 0.76)** | 0.94 (0.64, 1.38) | 1.00 | **0.46 (0.27, 0.80)** | 0.94 (0.64, 1.40) | 1.00 |
| Picloram (GW) | 1.10 (0.70, 1.73) | 0.84 (0.57, 1.22) | 1.00 | 1.10 (0.69, 1.75) | 0.83 (0.57, 1.22) | 1.00 | 1.07 (0.66, 1.74) | 0.83 (0.56, 1.23) | 1.00 |
| 2,4-D herbicide (P1E5) | **1.67 (1.03, 2.72)** | **1.53 (1.04, 2.25)** | 1.00 | 1.64 (0.99, 2.71) | **1.51 (1.02, 2.23)** | 1.00 | 1.39 (0.83, 2.33) | 1.47 (0.98, 2.21) | 1.00 |
| Heptachlor (F005) | 5.19 (1.00, 26.9) | 1.78 (0.40, 7.93) | 1.00 | **7.84 (1.27, 48.6)** | 2.48 (0.49, 12.5) | 1.00 | **8.53 (1.35, 54.0)** | 2.40 (0.51, 11.3) | 1.00 |
| Mesotrione (F005) | 0.71 (0.44, 1.13) | 0.80 (0.54, 1.17) | 1.00 | 0.69 (0.42, 1.12) | 0.78 (0.53, 1.16) | 1.00 | 0.74 (0.45, 1.22) | 0.80 (0.53, 1.20) | 1.00 |
| Mesotrione (F01) | 0.71 (0.44, 1.14) | 0.82 (0.56, 1.19) | 1.00 | 0.71 (0.44, 1.16) | 0.82 (0.56, 1.20) | 1.00 | 0.78 (0.47, 1.30) | 0.83 (0.55, 1.24) | 1.00 |
| Atrazine (P1E5) | 0.66 (0.41, 1.06) | 0.73 (0.49, 1.08) | 1.00 | 0.66 (0.40, 1.07) | 0.72 (0.48, 1.08) | 1.00 | 0.61 (0.37, 1.01) | 0.68 (0.45, 1.05) | 1.00 |
| **Pollutants** |  |  |  |  |  |  |  |  |  |
| Coarse PM<sub>2.5-10</sub> (P1E5) | **1.94 (1.16, 3.24)** | 1.29 (0.89, 1.88) | 1.00 | **1.87 (1.11, 3.14)** | 1.28 (0.87, 1.87) | 1.00 | **1.84 (1.07, 3.18)** | 1.23 (0.82, 1.84) | 1.00 |
| Fine PM<sub>2.5</sub> (P1E5) | 1.42 (0.88, 2.30) | 1.12 (0.77, 1.62) | 1.00 | 1.53 (0.93, 2.53) | 1.16 (0.80, 1.69) | 1.00 | 1.51 (0.90, 2.51) | 1.13 (0.78, 1.65) | 1.00 |
| NO<sub>2</sub> (GW) | 1.57 (0.97, 2.55) | 1.37 (0.92, 2.03) | 1.00 | 1.52 (0.93, 2.48) | 1.36 (0.91, 2.03) | 1.00 | 1.47 (0.87, 2.48) | 1.41 (0.91, 2.17) | 1.00 |
| **Lifestyle / body composition** |  |  |  |  |  |  |  |  |  |
| MDS (GW) | **1.73 (1.04, 2.86)** | 0.85 (0.58, 1.24) | 1.00 | **1.83 (1.08, 3.10)** | 0.87 (0.60, 1.28) | 1.00 | 1.67 (0.99, 2.81) | 0.86 (0.58, 1.27) | 1.00 |
| AHEI (GW) | 0.82 (0.51, 1.31) | 0.74 (0.51, 1.09) | 1.00 | 0.75 (0.46, 1.22) | 0.71 (0.48, 1.05) | 1.00 | 0.87 (0.53, 1.44) | 0.73 (0.49, 1.08) | 1.00 |
| AHEI (P1E5) | 0.73 (0.45, 1.17) | 0.71 (0.49, 1.04) | 1.00 | 0.67 (0.41, 1.10) | 0.68 (0.46, 1.01) | 1.00 | 0.75 (0.46, 1.24) | 0.70 (0.47, 1.04) | 1.00 |
| Smoking (GW) | 1.16 (0.73, 1.85) | 1.20 (0.83, 1.74) | 1.00 | 1.06 (0.66, 1.72) | 1.15 (0.79, 1.68) | 1.00 | 1.15 (0.70, 1.88) | 1.31 (0.88, 1.94) | 1.00 |
| Smoking-Maas (GW) | **1.71 (1.05, 2.79)** | 1.00 (0.68, 1.46) | 1.00 | 1.48 (0.89, 2.47) | 0.92 (0.62, 1.37) | 1.00 | 1.65 (0.98, 2.79) | 1.06 (0.70, 1.59) | 1.00 |
| BMI (GW) | 0.70 (0.44, 1.12) | 0.80 (0.54, 1.17) | 1.00 | 0.79 (0.48, 1.29) | 0.84 (0.57, 1.25) | 1.00 | 0.81 (0.49, 1.34) | 0.87 (0.58, 1.30) | 1.00 |
| Obesity (F005) | 1.37 (0.84, 2.24) | **1.84 (1.22, 2.78)** | 1.00 | 1.38 (0.83, 2.28) | **1.86 (1.22, 2.83)** | 1.00 | 1.44 (0.85, 2.42) | **1.94 (1.26, 2.99)** | 1.00 |
| ORs represent odds of early- or intermediate-onset diagnosis compared to late-onset associated with SD-change in MRS. **Bold:** P<0.05. Race modeled as White (ref), Black; ADI modeled continuously. MDS = Mediterranean Diet Score, AHEI = Alternative Healthy Eating Index. MRS thresholds: GW = genome wide, F005 = 5% false discovery rate, F01 = 10% false discovery rate, P1E5 = p-value<10<sup>-5</sup>. |  |  |  |  |  |  |  |  |  |

We assessed for nonlinearity, with select signatures shown in Fig. 2. Notably, picloram—for which we observed null effects across primary models—was identified to have nonlinear pattern. Signatures where we identified associations specific to a certain onset group in polytomous models, including heptachlor, NO_2_, PM_2.5_, smoking-Maas (all EOCRC), obesity (IOCRC), were also found to display nonlinear patterns with onset age. Supplemental material 1 shows nonlinear patterns were largely consistent with linear and polytomous models.

**Figure 2.**
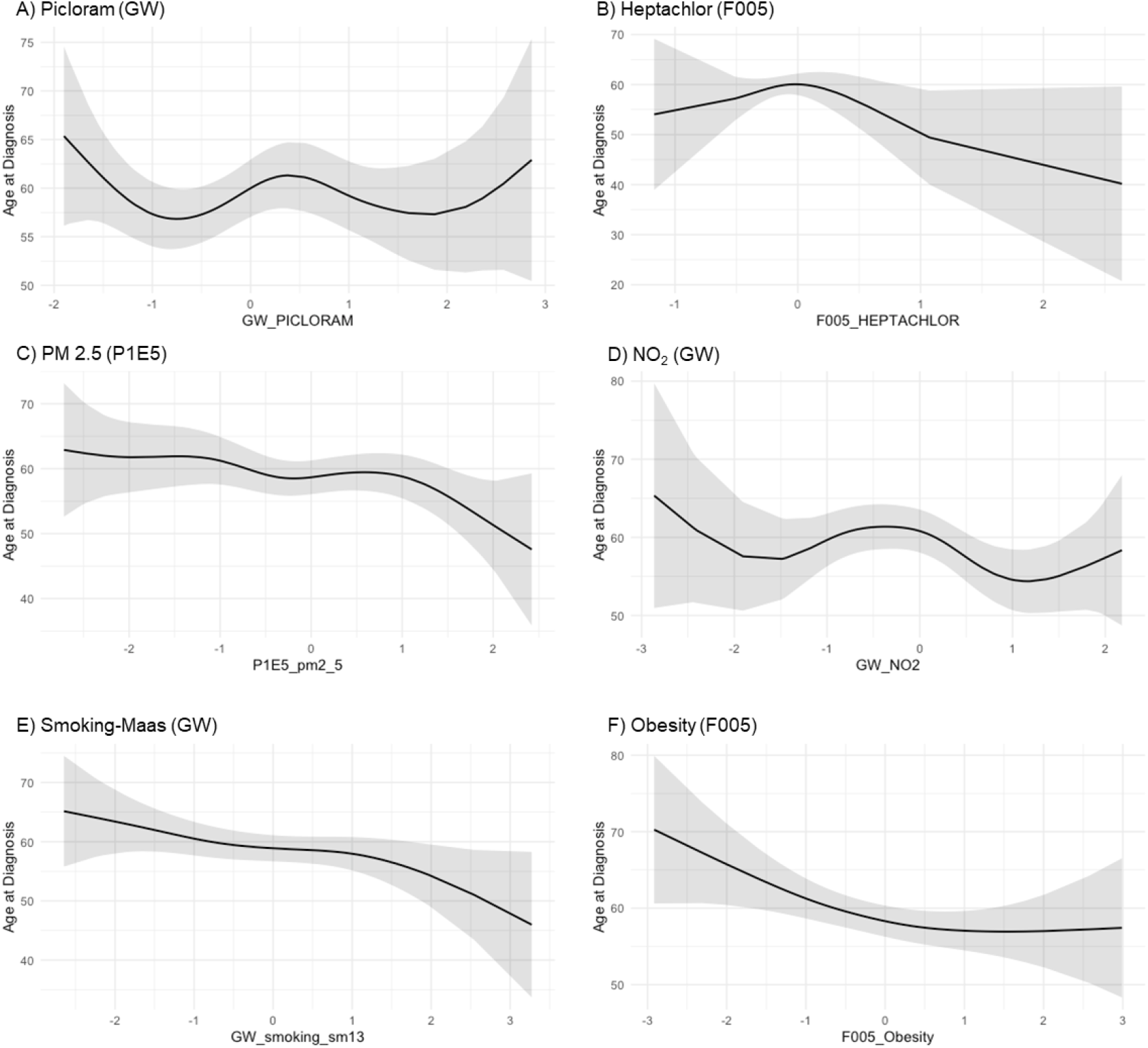
Nonlinear patterns between select epigenetic exposure signatures and age of CRC onset. Nonlinear patterns estimated by GAM. (A) picloram, (B) Heptachlor, (C) PM2.5, (D) NO_2_, (E) smoking-Maas, (F) obesity.

Fig. 3 shows stratified associations between signatures and onset age. We found potential heterogeneity according to race (Fig. 3a). NO_2_ was associated with younger onset and mesotrione (F005) was associated with older onset among Black participants specifically. In contrast, associations specific to White participants included heptachlor and obesity with younger onset and metolachlor (F01, P1E5) and atrazine with older onset. In high-deprivation areas (Fig. 3b), signatures for lindane and mesotrione (F005) were associated with older onset and the MDS signature was associated with younger onset. Heptachlor (F005) was more strongly associated with younger onset in low-deprivation areas compared to high-deprivation areas. Despite a limited sample from Louisiana (n=13), we found potential heterogeneity by state (Fig. 3c), where Louisiana cases appear to drive the associations between MRSes for picloram and MDS with younger onset. We found significant divergent heterogeneity in the association between mesotrione (F005) and onset by MEEI, associated with older onset in high-environmental burden areas and younger onset in low-environmental burden areas (Fig. 3d). Other associations specific to high-environmental burden areas included NO_2_ with younger onset and mesotrione (P1E5) and AHEI (GW) with older onset. In contrast, signatures for 2,4-D and smoking-Maas showed associations with younger onset specifically among low-environmental burden areas. Supplemental Material 2 shows stratified nonlinear patterns between signatures and age of onset.

**Figure 3.**
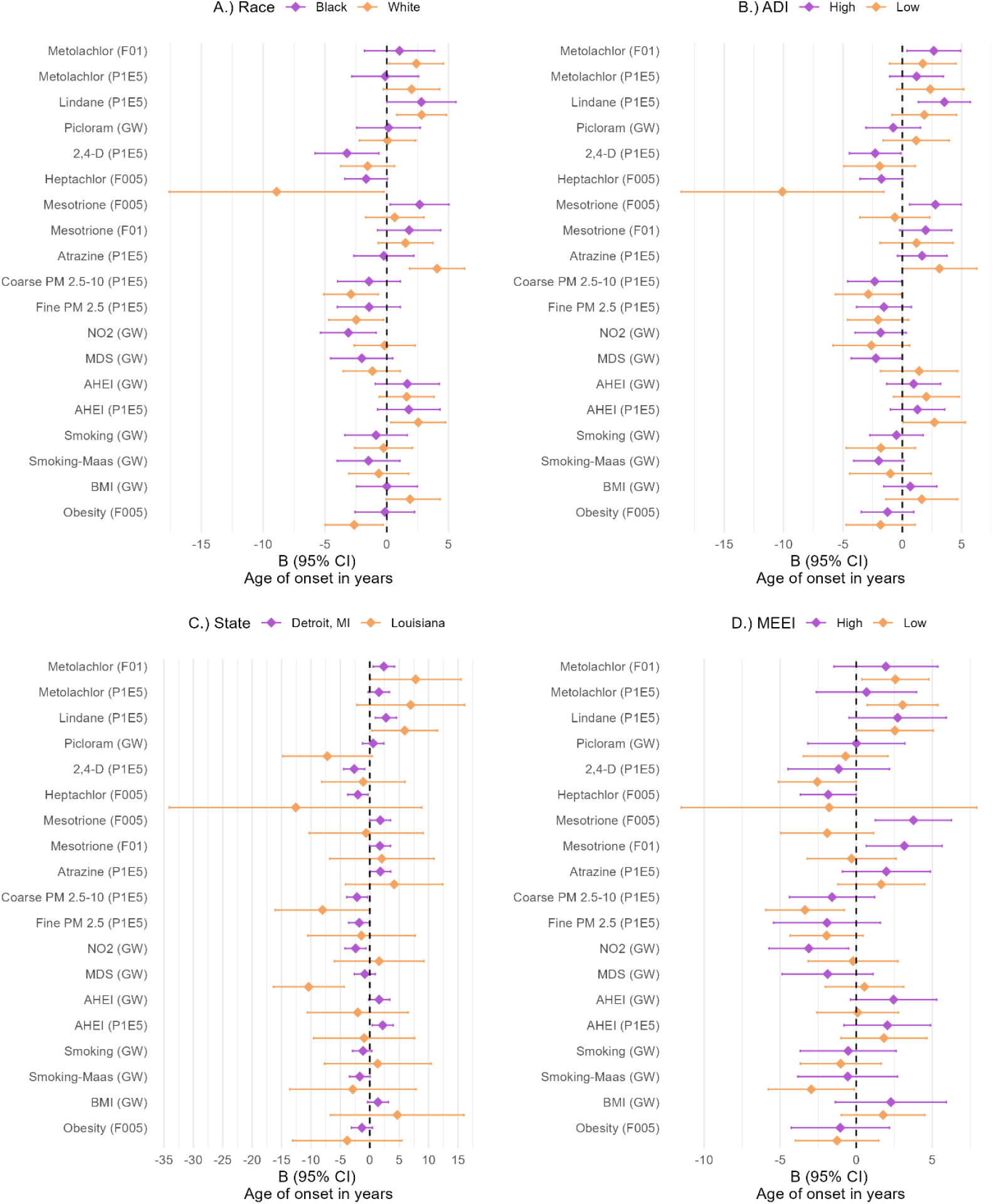
Associations between MRSes and age of onset in years, stratified by A) race, B) ADI, C) state, D) MEEI. Beta coefficients represent the difference in years for age of onset per SD-change in MRS. ADI= Area Deprivation Index; high n=96, low n=44. MEEI = Multi-Exposure Environmental Index; high n=52, low n=56.

Supplemental Material 3 reports p-values adjusted for Benjamini-Hochberg correction. Using a 20% FDR, we found signatures for metolachlor, lindane, 2,4-D and coarse PM maintained p<0.05 with age of CRC onset. Using a 20% FDR in the polytomous models, we found signatures for lindane, metolachlor, coarse PM, MDS, and smoking-Maas maintained significance with EOCRC and the signatures for 2,4-D, metolachlor, and obesity maintained significance with IOCRC.

Correlation heatmaps between MRSes and measured exposures for environmental factors and lifestyle factors are shown in Fig. 4. ADI and MEEI show weak but statistically significant correlations with MRSes (Fig. 4a), positively correlated with 2,4-D herbicide (P1E5) and PM_2.5-10_ signatures and negatively correlated with lindane, mesotrione (F01), and metolachlor (F01) signatures. The MRSes for 2,4-D, metolachlor (F01), and mesotrione (F005) showed the most significant correlations with observed environmental exposures. Notably, no air pollution signatures were correlated with census tract-level PM_2.5_. Census tract-level rank of pesticides was significantly positively correlated with metolachlor (F01, P1E5) but no other pesticide signatures. Lifestyle/body composition exposures were generally not correlated with corresponding MRSes (Fig. 4b).

**Figure 4.**
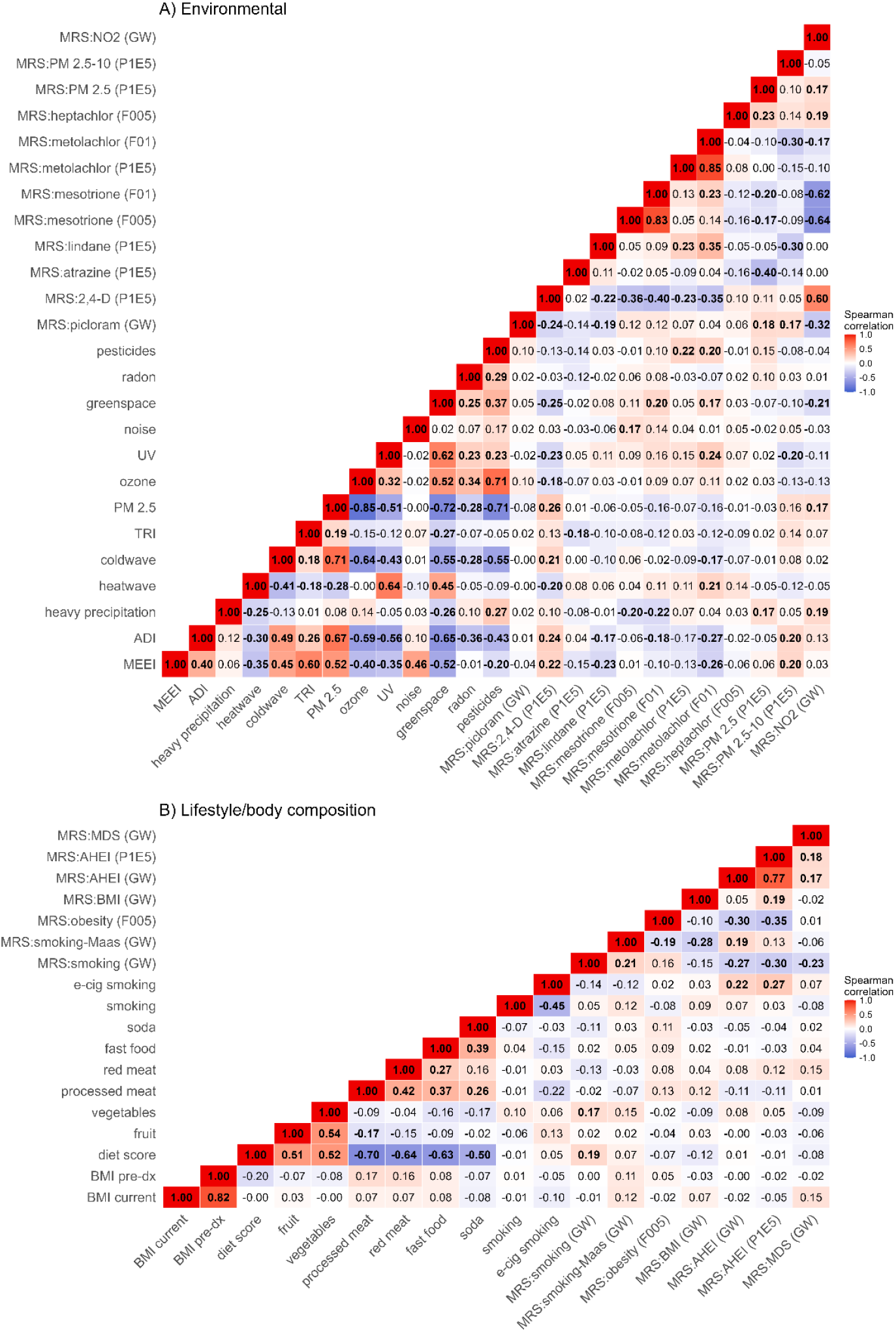
Correlation heatmap of MRSes and corresponding exposures. Panel A: environmental MRSes and exposures. Panel B: lifestyle / body composition MRSes and exposures.

## Discussion

In this racially, socioeconomically, and geographically diverse CRC cohort, we found EOCRC cases display distinct epigenetic exposure signature patterns from LOCRC. Moreover, epigenetic exposure signatures for IOCRC cases had overlapping features for both EOCRC and LOCRC. These results are supportive of birth cohort effects observed within CRC and provide preliminary evidence of potential drivers for the significant rise in EOCRC over time.

This study presents the first application of an epigenetic exposome fingerprinting framework to a racially representative CRC cohort, building directly on recent findings in predominantly White populations.^10^ The specific exposures driving the generational rise in EOCRC remain largely undefined, and the evidence-to-date points toward environmental and lifestyle factors.^23^ By design, we sought to directly replicate the initial findings in our CRC cohort to limit multiple testing from an agnostic approach. Our findings are broadly consistent with Maas *et al*.’s,^10^ showing evidence that age of CRC onset is associated with epigenetic exposure signatures for pesticides and pollutants, with relatively few associations with lifestyle or body size signatures. Specifically, we found epigenetic exposure signatures for 2,4-D herbicide, heptachlor, coarse PM, smoking-Maas, and Mediterranean diet were associated with EOCRC, with signatures for fine PM and NO_2_ marginally non-significant. We also showed that an obesity epigenetic exposure signature was associated with IOCRC but not EOCRC. Signatures for metolachlor may be specifically associated with LOCRC, and lindane may be associated with IOCRC and LOCRC. Results from this DANCE pilot cohort both replicate key associations from Maas *et al*. and reveal important divergences that highlight the value of studying diverse populations in this framework.

Given the likely environmental role in early-onset cancers, characterizing historical exposures is essential to characterizing novel risk factors that can lead to new avenues to prevent EOCRC.^24^ Somatic alterations, such as mutational and epigenetic exposure signatures, offer a distinct advantage over traditional exposure assessment methods, which typically rely on self-report, administrative records, or area-level proxies that are subject to bias. Because DNA methylation alterations are induced by the biologically active dose of a toxicant reaching target tissue rather than by estimated or modeled exposure alone, methylation-based signatures can capture inter-individual variation in absorption, metabolism, and detoxification that external exposure metrics cannot.^7^ This is particularly relevant for pesticides and air pollutants, where ambient or occupational exposure estimates may poorly reflect the internal dose actually experienced by an individual. Epigenetic exposure signatures have also demonstrated replication across diverse disease contexts,^25-27^ including cardiovascular disease, respiratory disease, and multiple cancer types, suggesting methylation marks capture generalizable biological responses to exposure rather than disease-specific artifacts. This cross-disease replication strengthens confidence that the signatures identified here reflect genuine exposure-related biology rather than CRC-specific confounding. Taken together, these properties make epigenetic exposure signatures a powerful tool for exposome research, particularly in diseases with long latency periods, where the exposures most relevant to carcinogenesis may have occurred decades before diagnosis and are no longer directly measurable.

The pesticide findings in DANCE are particularly intriguing in relation to the results from Maas and colleagues. Whereas Maas *et al*. found the picloram signature to be significantly elevated in EOCRC^10^—consistent with greater picloram exposure in individuals born after widespread agricultural adoption of this herbicide—DANCE generally showed no associations with picloram (other than among Louisiana cases) and instead identified 2,4-D and heptachlor signatures as significantly associated with earlier CRC onset. We also showed that metolachlor and lindane were associated with later onset. Given the rise of EOCRC started with 1960 birth cohorts,^4^ comparing birth cohorts to the timing of pesticide federal status (Fig. 5) may offer a useful heuristic. Lindane was introduced for US commercial use in 1945, phased out in 2002, and federally banned in 2007. As a result, more recent birth cohorts would likely have lower long-term exposure, consistent with our result showing lindane linked to later onset. In contrast, picloram was introduced in the US in 1964 with continued use today. This timing aligns with the start of the EOCRC epidemic, but we generally did not detect any associations with CRC onset. However, picloram is a restricted use pesticide, which may explain why results were null other than an association found among Louisiana cases. Further, we detected nonlinear patterns between the picloram signature and CRC onset age, which could also explain the discordance between our results and those from Maas *et al*.

**Figure 5.**
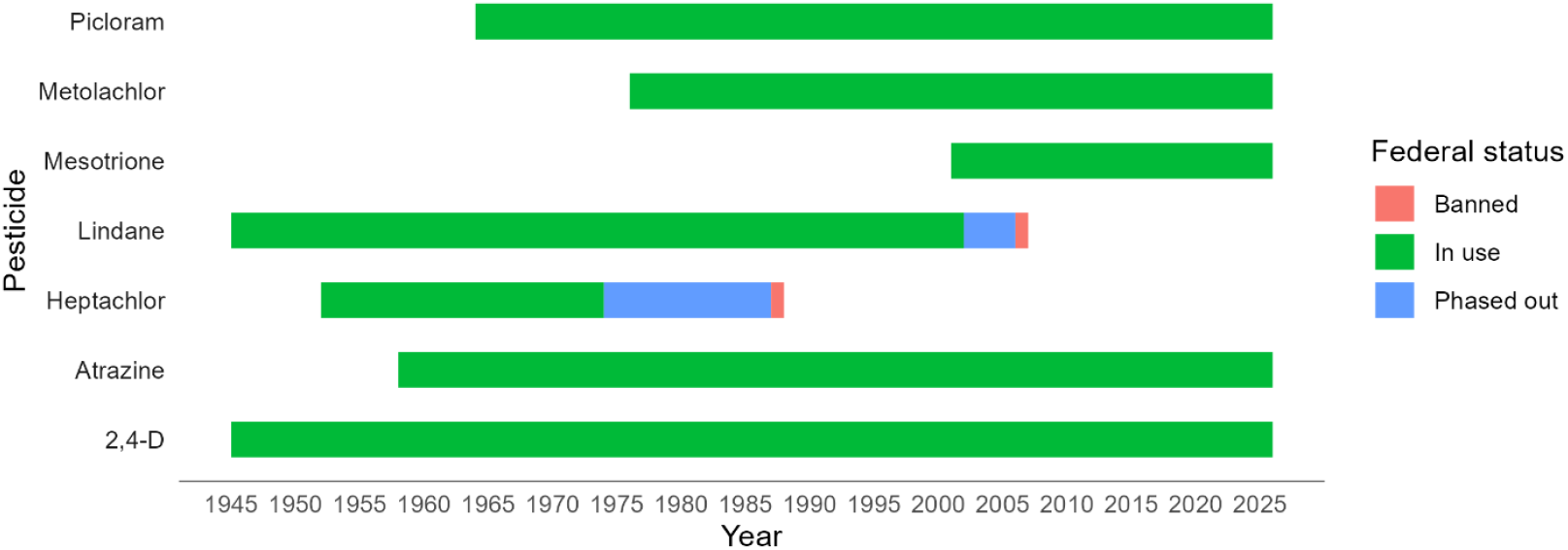
Timing of pesticide use and federal regulation status. Figure shows pesticide timing and US federal regulation status.

Our findings that signatures for 2,4-D and heptachlor are linked to EOCRC are surprising. 2,4-D herbicide has been used in the US since the 1940s on an array of agricultural crops and non-agricultural uses, thus exposure is not specific to more recent birth cohorts. However, US Geological Survey data show a marked increase in 2,4-D agricultural use over time, particularly since 2005.^28^ Further, the varied uses of 2,4-D present multiple possible exposure routes.

Heptachlor is an organochlorine insecticide that was phased out in 1974 and federally banned in 1988.^29^ While it has been identified as possibly carcinogenic, the majority of EOCRC cases would be from birth cohorts after heptachlor was banned. However, its main degradation byproduct, heptachlor epoxide, is environmentally persistent and bioaccumulates, including in the Great Lakes.^30^ Further, numerous pesticides, including organochlorines, have been shown to be detectable over time within house dust, thus heptachlor may present low-dose but persistent exposure well past its banning.^31,32^ Finally, heptachlor is heavily lipophilic. Given most EOCRC cases in DANCE were overweight or obese, younger participants may have higher exposure to heptachlor through adiposity reservoirs even if they were exogenously exposed for a relatively short time period. Such temporal exposure dynamics of pesticides, influenced by the specific agricultural history of the US Southeast and Midwest, may explain meaningful differences between DANCE and the Maas *et al*. cohorts.

Our study supports further investigation of pesticides as a potential driver of rapidly rising rates of EOCRC.^33^ According to a National Research Council report, pesticide treatment of crops skyrocketed from 3% in 1955 to 95% by the 1980’s,^34^ with a 10-fold increase in herbicide use during this period.^35^ Though pesticide use in the US peaked in 1981, it marginally plateaued at roughly 500 million pounds per year.^35^ While this is particularly concerning for high-exposure populations—such as farmers and residents of agricultural areas—pesticide exposure may occur through indirect routes, including runoff into water systems, long-distance transportation by wind, and diet, and thus may be a population-wide exposure.^36,37^

The coarse PM association with EOCRC is a novel finding not highlighted in Maas *et al*. and may reflect the specific geographic and residential histories of DANCE participants, who were recruited from metro-Detroit and Louisiana, areas with elevated industrial and traffic-related particulate pollution.^38,39^ Our results indicate a PM_2.5–10_ gradient declining across onset groups, consistent with a generational increase in coarse particulate exposure among younger patients, potentially linked to urban residential patterns and occupational factors.^40,41^ We also identified directional evidence that fine PM and NO_2_ signatures are higher among early-onset cases than late-onset and may be associated with EOCRC. While federal regulations under the Clean Air Act have led to reductions in air pollution in the US, not all pollutants have declined. Namely agricultural and energy sector pollutant concentrations have increased over time—sectors that are specifically relevant for DANCE geographies.^42^ Moreover, stratified results suggest these findings were most consistent among Detroit-based participants, consistent with the area’s history of auto manufacturing and resulting pollution.^43^ Outdoor air pollution as a broad mixture is classified as a Group 1 carcinogen.^44^ Further, as the prevalence of other major carcinogens decrease, like smoking, cancer risk due to air pollution may grow as its relative burden increases.^45^ Pollutants, including PM and NO_2_, are associated with higher CRC risk across, although with substantial heterogeneity.^46-49^ Our group analyzed data from the Metropolitan Detroit Cancer Surveillance System, noting the effect of PM_2.5_ on mortality was stronger among non-Hispanic Black CRC cases than non-Hispanic White and especially strong among non-Hispanic Black cases with EOCRC.^50^ While NO_2_ has not been specifically classified as a carcinogen, it may proxy traffic-related chemical components or ultrafine PM.^51^ The mechanistic link between air pollution and EOCRC in our study warrants further investigation with residential history data and geocoded pollution estimates in the full DANCE cohort.

The replication of the smoking-Maas signature associated with CRC onset in DANCE is a critical finding.^10^ Despite differences in cohort composition, geographic context, and tumor biospecimen type (FFPE in DANCE vs. fresh-frozen in most Maas *et al*. publicly available cohorts), we observed higher epigenetic signatures for smoking was associated with EOCRC. This consistency across cohorts and racial groups strengthens the evidence that smoking-related DNA methylation changes are a meaningful correlate of younger CRC onset, and that these epigenetic markers persist in tumor tissue from FFPE specimens. However, that we and Maas *et al* found epigenetic exposure signatures for smoking were higher among EOCRC is perplexing. While current and former smoking status has been associated with risk for both early- and late-onset CRC in observational studies, there has not been evidence of heterogeneity by age of onset.^52,53^ Further, the prevalence of cigarette smoking has declined over time, thus does not match the exposure pattern indicative of the EOCRC birth cohort effects, but vaping has risen among young adults and may explain our results.^54^ This pattern is consistent with our sample, where prevalence of never smoking and vaping was highest among early-onset cases. While, it is possible there is under-reporting of smoking, our results speak to the complexity of epigenetic exposure signatures and suggest smoking-Maas may proxy other exposures (historical or inherited), despite its validation.^12^

The overrepresentation of Black cases and cases from high-ADI neighborhoods among the early-onset group in DANCE (65.5% EOCRC vs. 31.3% LOCRC) underscores the intersection of racial health disparities, neighborhood deprivation, and environmental exposure in EOCRC risk. Prior epigenomic studies have focused on predominantly non-Hispanic White/European Ancestry cohorts and thus failed to capture this dimension of disease etiology.^55^ Notably, we found that epigenetic exposure signatures maintained significant effects even after adjustment for race or ADI, factors strongly associated with CRC onset.^56,57^

The racial, socioeconomic, and geographic diversity of DANCE also provides insights into contextual determinants of epigenetic signatures. We found associations between heptachlor and age of onset were particularly strong among non-Hispanic White participants and in low-deprivation neighborhoods. While unexpected, heptachlor was an insecticide often used indoors for pest control. Because housing in low-income, majority Black, or renter-occupied neighborhoods have been documented to face negligence and unmitigated pest presence,^58,59^ socioeconomic advantage may paradoxically accompany higher exposure to heptachlor or other insecticides and could explain the observed associations. Alternatively, gentrified neighborhoods are areas of recent upward socioeconomic transition that may concentrate young White individuals to historical buildings with residual heptachlor exposure.^60^ We observed the association between the NO_2_ signature and earlier CRC onset was specific to Black cases. This result could be indicative of racialized NO_2_ exposure gradients and a possible source of racial disparities in EOCRC risk.^61^ Moreover, ADI and MEEI were significantly correlated with epigenetic exposure signatures for pesticides and pollutants, but the direction of correlation was not always the same. For example, correlations for ADI and MEEI were positive with 2,4-D and coarse PM, but were negatively correlated with most other pesticides, suggesting some pesticide signatures were higher among low-deprivation and low-environmental exposure areas. While there are significant disparities in EOCRC,^57,62^ the burden of EOCRC exists across sociodemographic characteristics, thus the etiology behind rising rates is likely a complex interaction across social, environmental, and lifestyle factors. Our results attest to this complexity between socioenvironmental advantage/disadvantage and its potential influence on CRC onset and highlight the strengths of using a diverse cohort to investigate EOCRC.

Our study was further strengthened by using multiple statistical approaches that corroborated one another. Specifically, we used polytomous regression and GAM to detect associations linked to specific onset age groups. As a result, we found numerous signatures show linear associations with age of onset while other signatures may be particular to specific birth cohorts.

This study is limited by the pilot sample size (n=146) and reduced power. Due to the small sample size, we largely relied on crude or single-covariate models. Future analyses in the full DANCE cohort will incorporate multiple participant, clinical, and socioeconomic factors as covariates. We also relied on single-signature models, but individuals are exposed to environmental and exogenous factors as mixtures. Future studies should consider mixtures and interactions between exposures and epigenetic signatures. Further, while the DANCE pilot cohort introduces necessary diversity, only 10% of the pilot sample was from Louisiana. The full DANCE cohort will have greater representation of Louisiana cases. The use of FFPE tumor tissue introduces potential technical variation relative to fresh-frozen specimens, and the MRS framework was originally validated in fresh-frozen cohorts; however, the replication of the smoking signal in FFPE tissue is encouraging. Finally, the MRS methodology provides epigenetic proxies for exposure rather than direct measurements, and associations should be interpreted as hypothesis-generating pending validation with longitudinal measured exposure data. While we evaluated correlation between signatures and exposure data, the epigenetic exposure signatures are theorized to capture exposure across the lifecourse,^8^ whereas our data are cross-sectional, limiting our ability to evaluate agreement. This clarifies the need to incorporate residential histories and evaluate spatio-temporal-specific environmental exposures.

## Conclusion

We identified distinct epigenetic exposure signatures across CRC onset groups, consistent with cohort effects. These findings from the DANCE pilot cohort demonstrate that epigenetic exposure signatures meaningfully distinguish EOCRC from later-onset CRC in a racially, socioeconomically, and geographically diverse US population, with partial replication and important divergences from prior findings.^10^ The smoking epigenetic signature replicates robustly, while pesticide and pollution exposure profiles diverge in ways consistent with the distinct agricultural and industrial exposure histories of US populations specific to DANCE geographies. Future research is needed to replicate our and Maas *et al*.’s findings, particularly from cohorts with residential histories and longitudinal exposure data. Nonetheless, the pesticide and pollution results collectively implicate environmental policy as a potentially important tool to curb growing rates of EOCRC and reduce disparities. In contrast to individually-tailored strategies, such as primarily focusing on diet and lifestyle, policy intervention would lead to significant absolute reductions of EOCRC and improved population health.^63^ The disproportionate burden of EOCRC among Black participants residing in high-deprivation neighborhoods underscores the importance of environmental justice considerations in EOCRC research. Fully powered analyses in the complete DANCE cohort will enable definitive characterization of these associations and their interaction with race, socioeconomic status, and residential history, generating actionable insights for EOCRC prevention in a systematically underrepresented population.

## Acknowledgements

SM Lima is supported by NCI T32 Postdoctoral Fellowship, T32-CA261787. This work was supported, in part, by NCI Award Number R01-CA259420, U01-CA199240, the Epidemiology Research Core, and the National Cancer Institute Center Grant (P30-CA022453) awarded to the Barbara Ann Karmanos Cancer Institute at Wayne State University. This research was supported [in part] by the Intramural Research Program of the National Institutes of Health (NIH). The contributions of the NIH author(s) are considered Works of the United States Government. The findings and conclusions presented in this paper are those of the author(s) and do not necessarily reflect the views of the NIH or the U.S. Department of Health and Human Services.

## Disclosures

Authors have no conflicts to disclose.

## Data availability

Data on epigenetic exposure signatures used for replication are available at https://github.com/CancerCompBioLab/EOCRCexposome. DANCE data cannot be shared publicly due to privacy of individuals that participated in the study. The data will be shared on reasonable request to the corresponding authors.

## Notes

### Competing Interest Statement

The authors have declared no competing interest.

### Author Declarations

IRBs of Georgetown University, Wayne State University, and Louisiana State University gave ethical approval for this work.

