## Supplemental Material 1 for "Epigenetic Exposure Signatures Distinguish Early-Onset from Later-Onset Colorectal Cancer in a Racially Diverse US Cohort: Findings from the Disparities and Cancer Epidemiology (DANCE) Study"

**Supplemental Material 1. Results from generalized additive models (GAMs) capturing non-linear associations between epigenetic exposure signatures and age of CRC onset.**

*Pesticides*

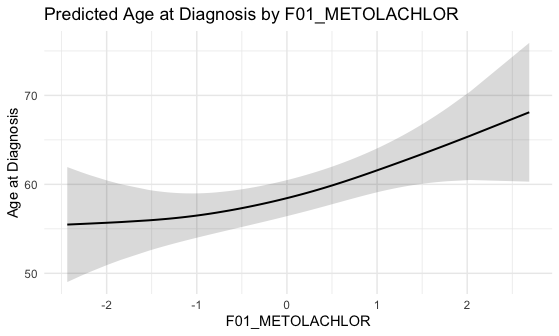

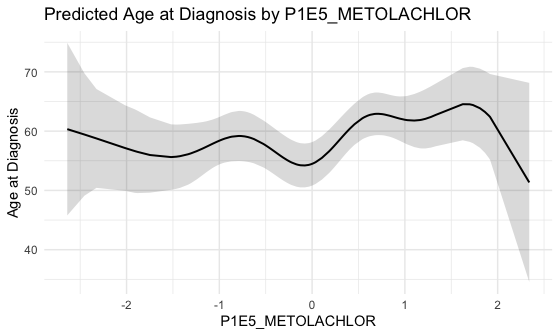

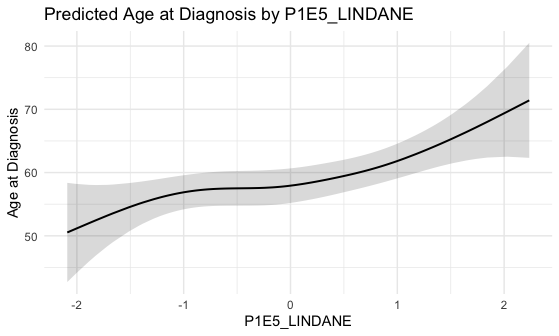

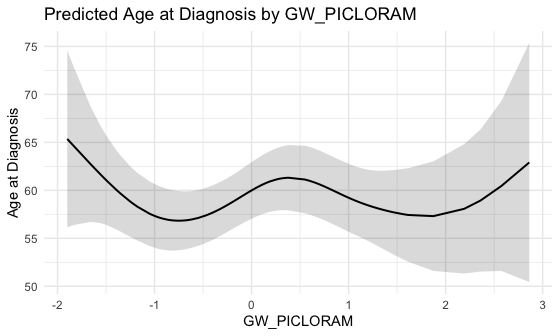

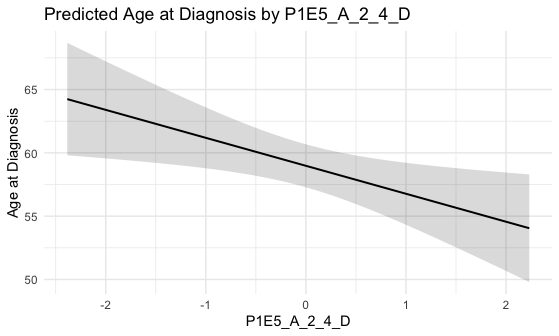

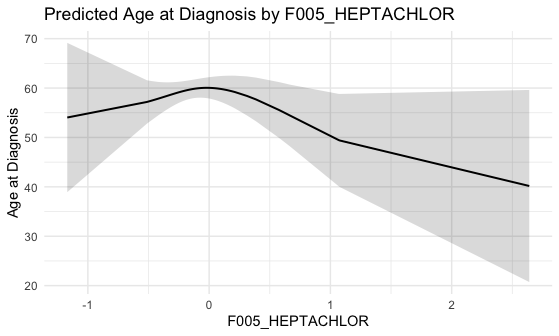

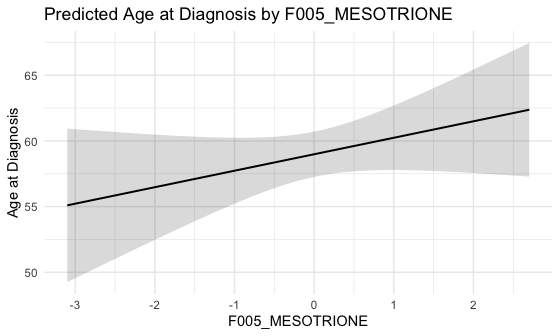

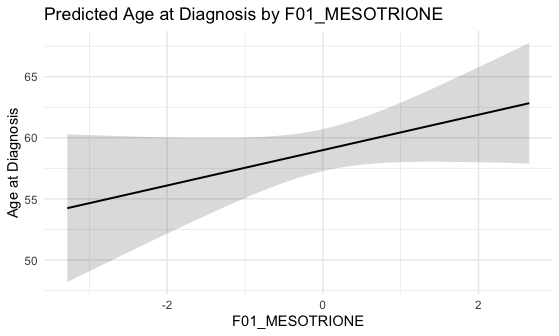

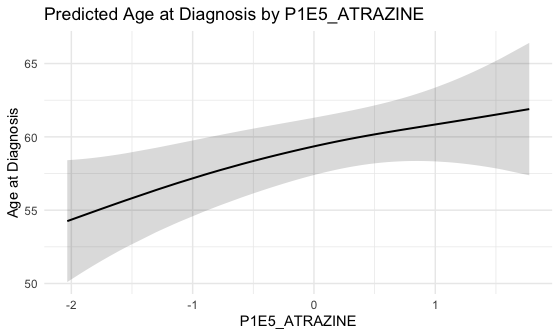

*Pollutants*

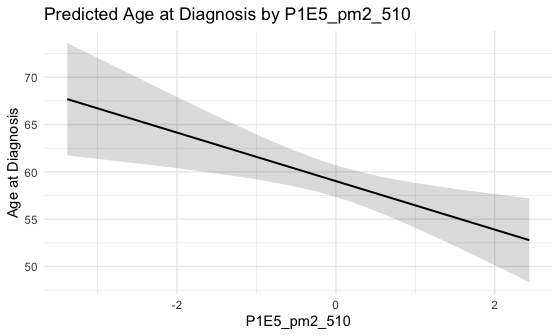

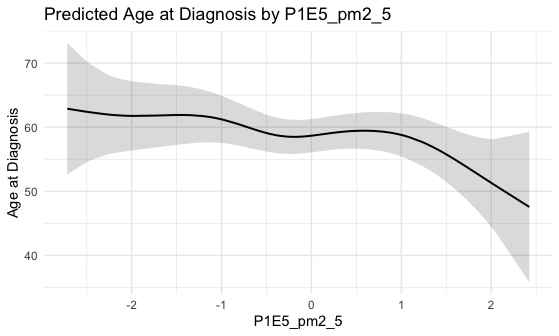

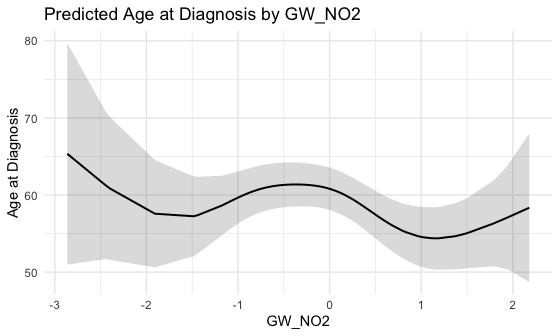

*Lifestyle & Body Size*

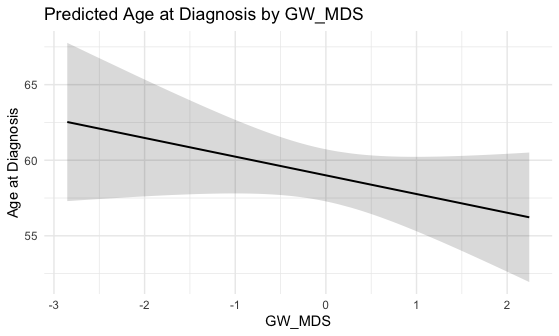

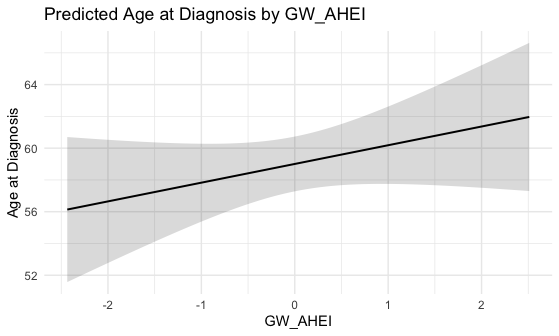

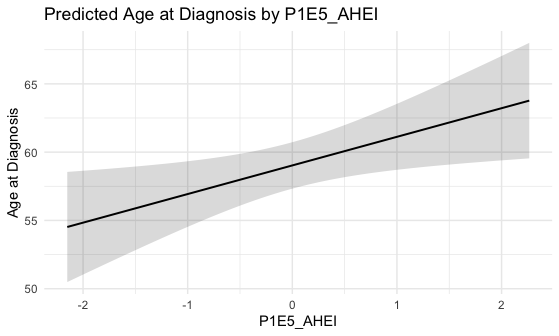

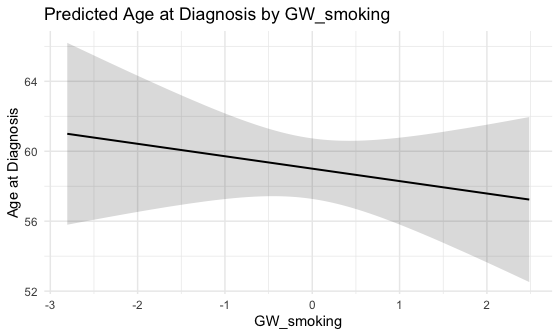

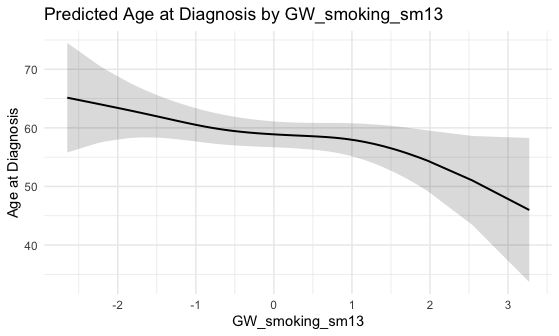

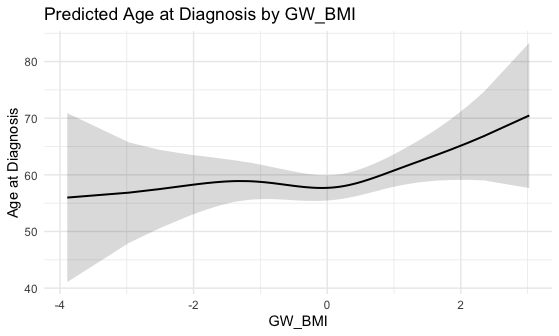

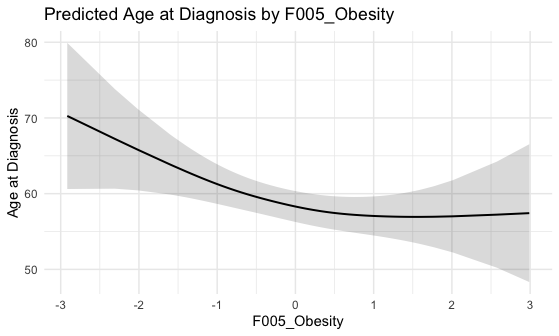
