## Supplemental Material 2 for "Epigenetic Exposure Signatures Distinguish Early-Onset from Later-Onset Colorectal Cancer in a Racially Diverse US Cohort: Findings from the Disparities and Cancer Epidemiology (DANCE) Study"

**Supplemental Material 2. Results from generalized additive models (GAMs) capturing non-linear associations between epigenetic exposure signatures and age of CRC onset, stratified by state, race, MEEI, and ADI.**

[Age of CRC onset vs MRS using GAM - By MEEI (L:Low, MEEI <=2, H:High, MEEI >2)	10](#_Toc238053075)

[Age of CRC onset vs MRS using GAM - By 2015 National ADI (L:Low, ADI <=50, H:High, ADI >50)	15](#_Toc238053076)

[Age of CRC onset vs MRS using GAM - By MEEI (L:Low, MEEI <=2, H:High, MEEI >2)	24](#_Toc238053080)

[Age of CRC onset vs MRS using GAM - By 2015 National ADI (L:Low, ADI <=50, H:High, ADI >50)	26](#_Toc238053081)

[Age of CRC onset vs MRS using GAM - By MEEI (L:Low, MEEI <=2, H:High, MEEI >2)	36](#_Toc238053085)

[Age of CRC onset vs MRS using GAM - By 2015 National ADI (L:Low, ADI <=50, H:High, ADI >50)	39](#_Toc238053086)

### *Pesticides*

#### Age of CRC onset vs MRS using GAM - By States (M:Detroit, Michigan, L:Louisiana)

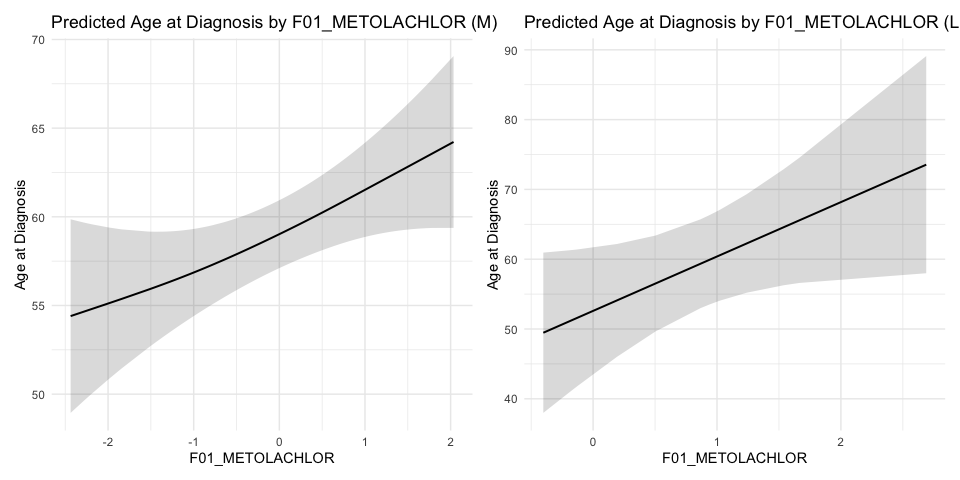

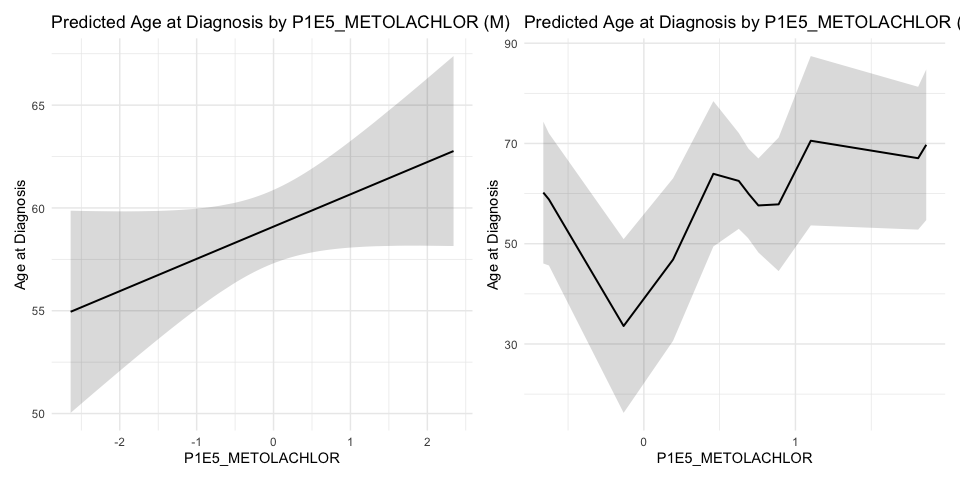

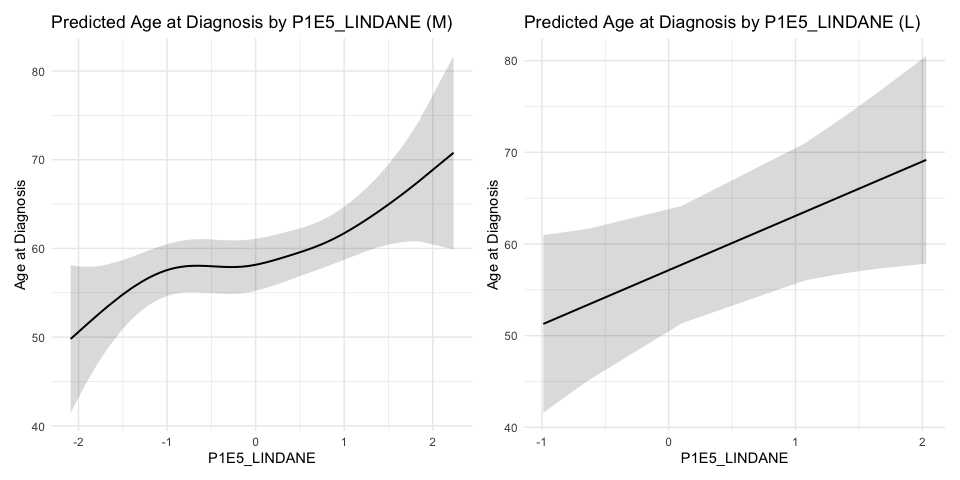

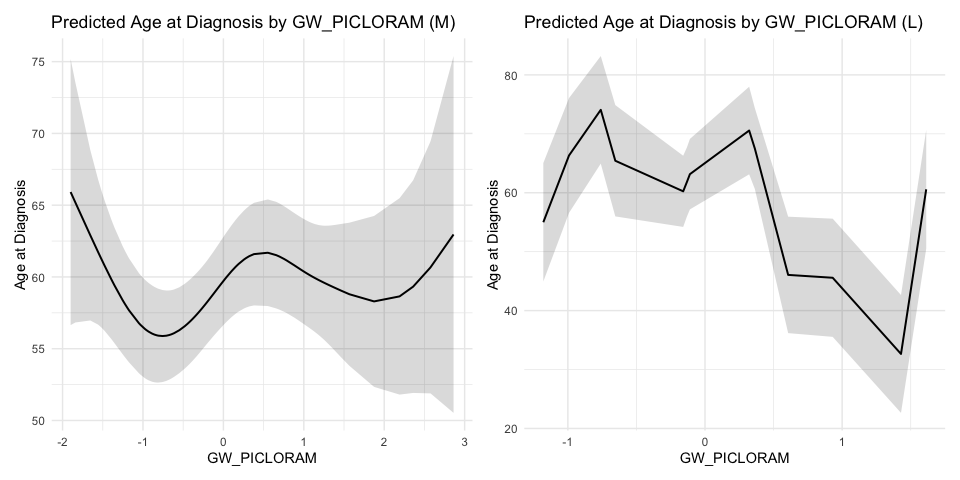

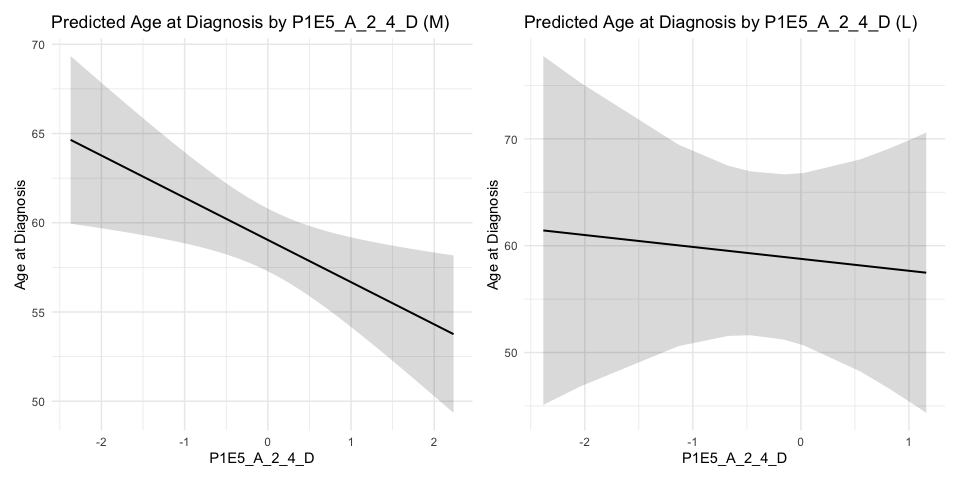

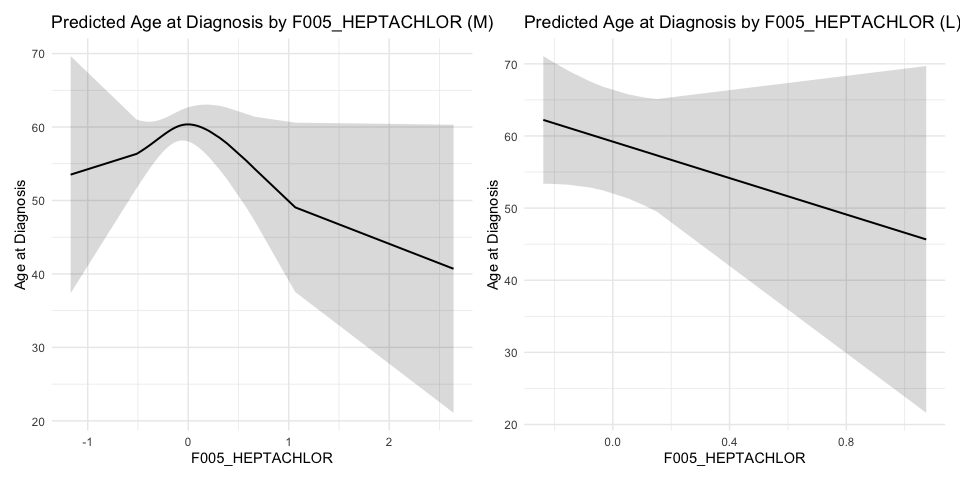

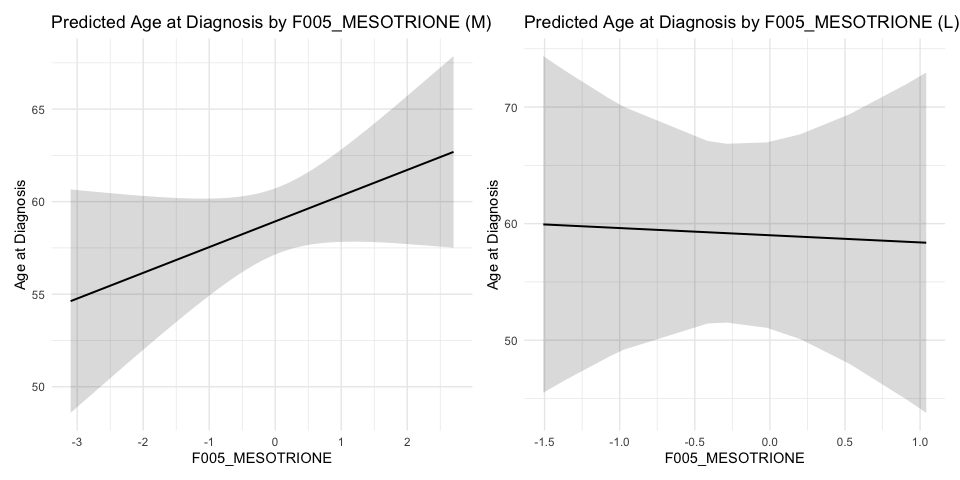

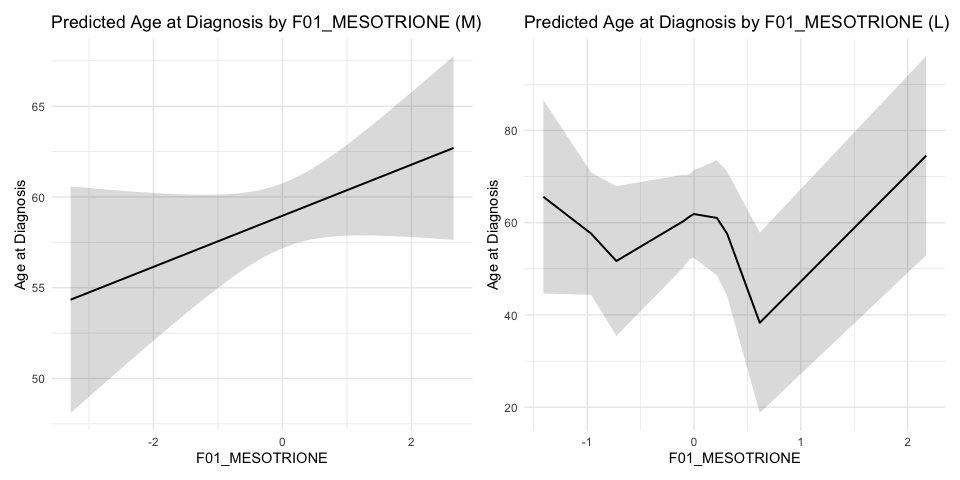

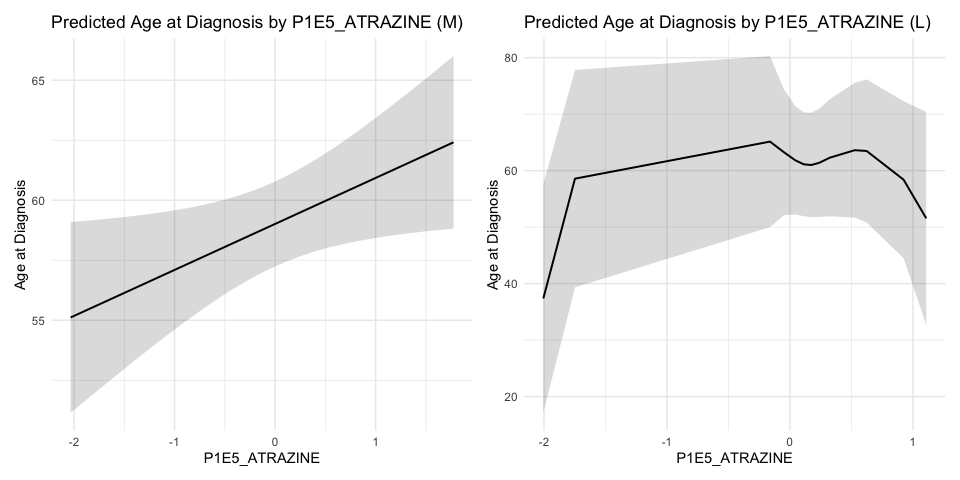

#### Age of CRC onset vs MRS using GAM - By RACE (AA:African American, NHW:NonHispanic White)

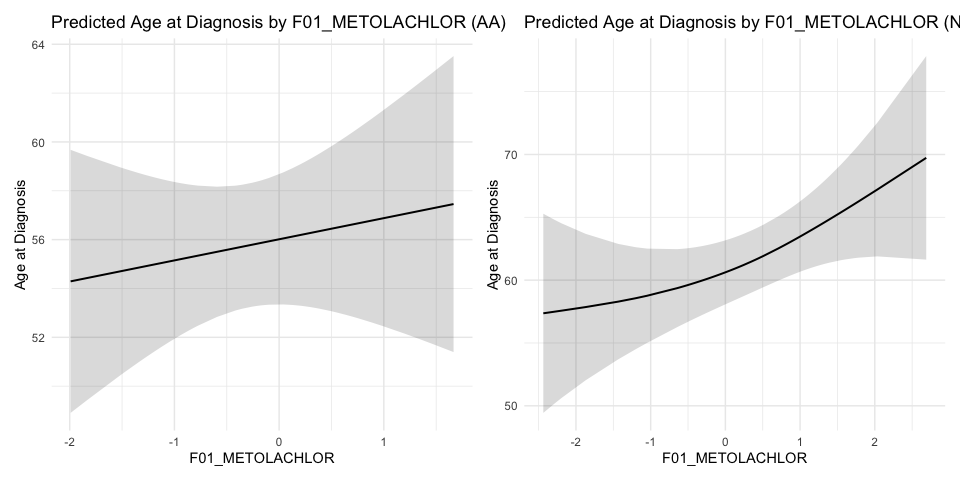

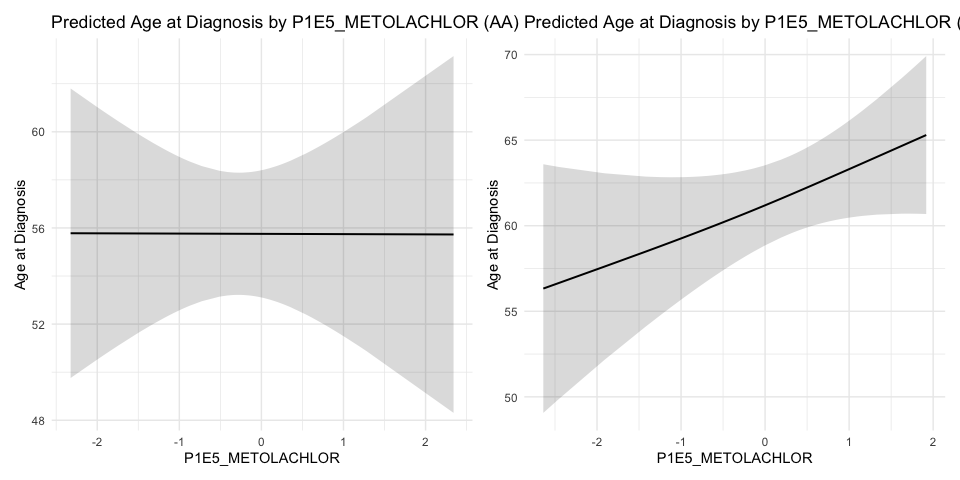

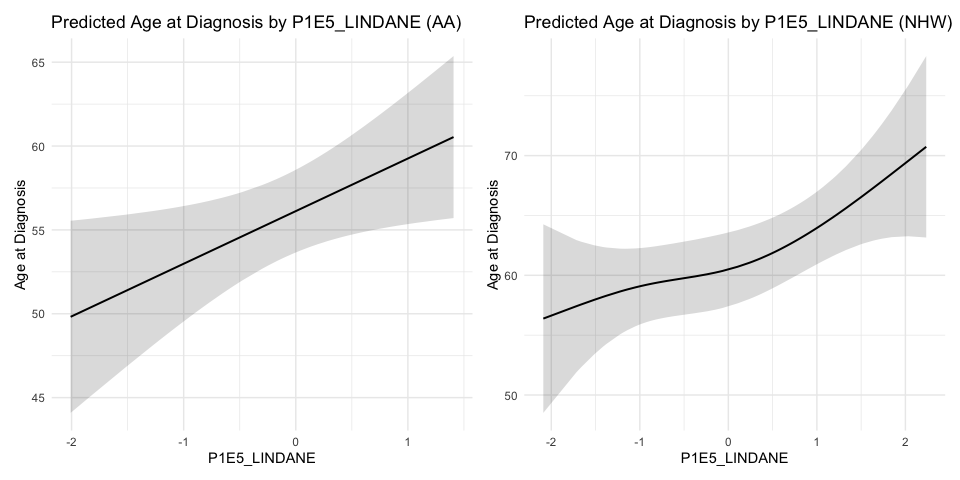

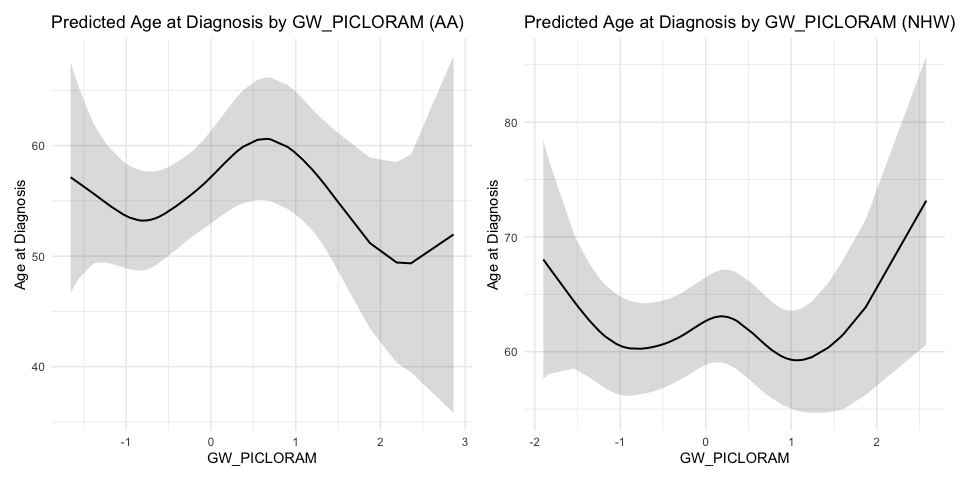

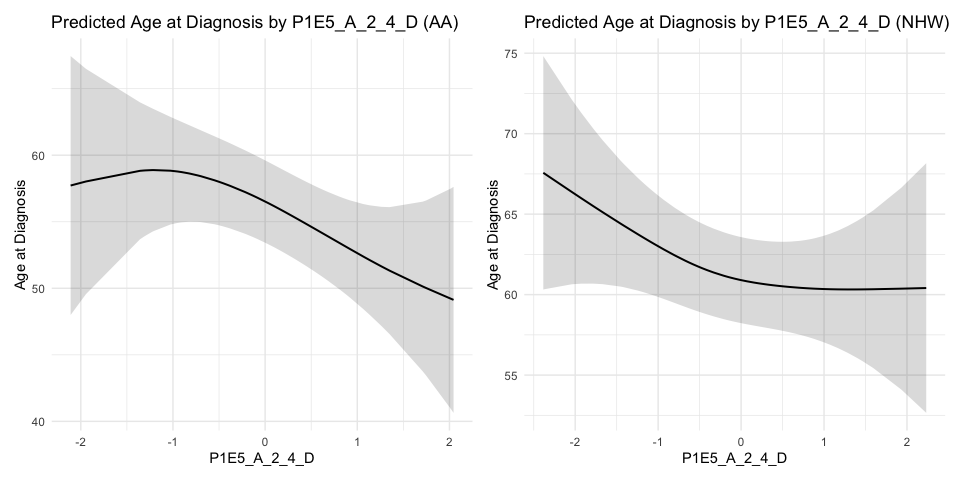

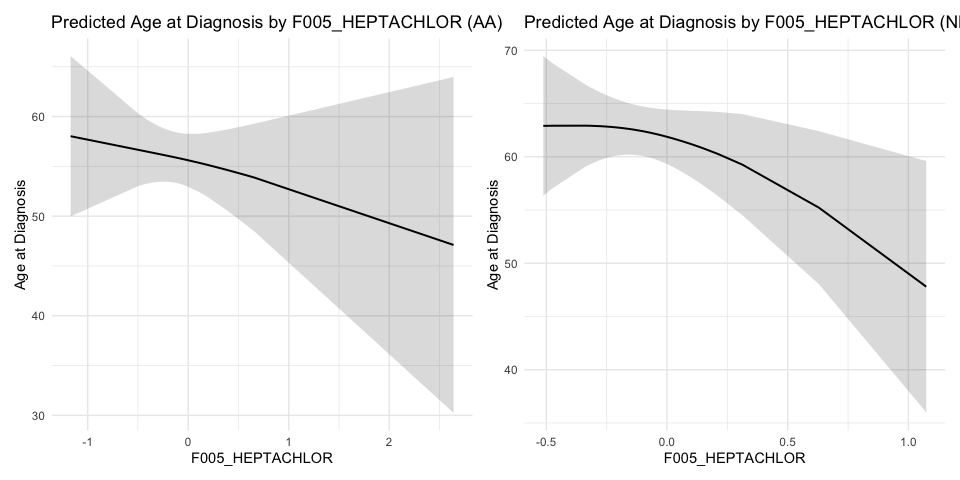

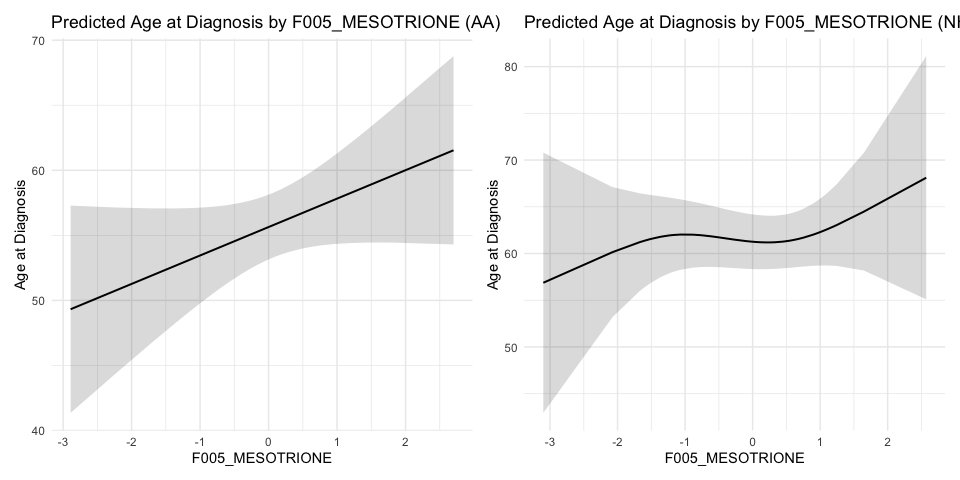

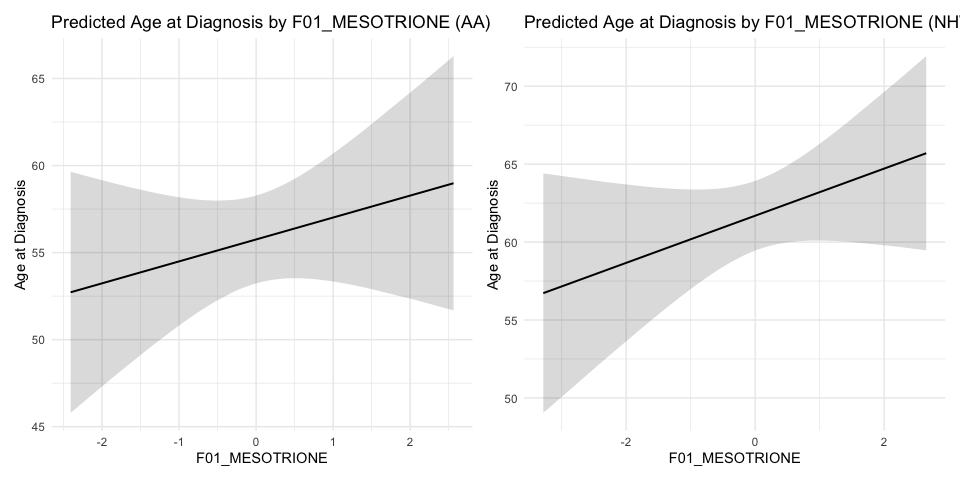

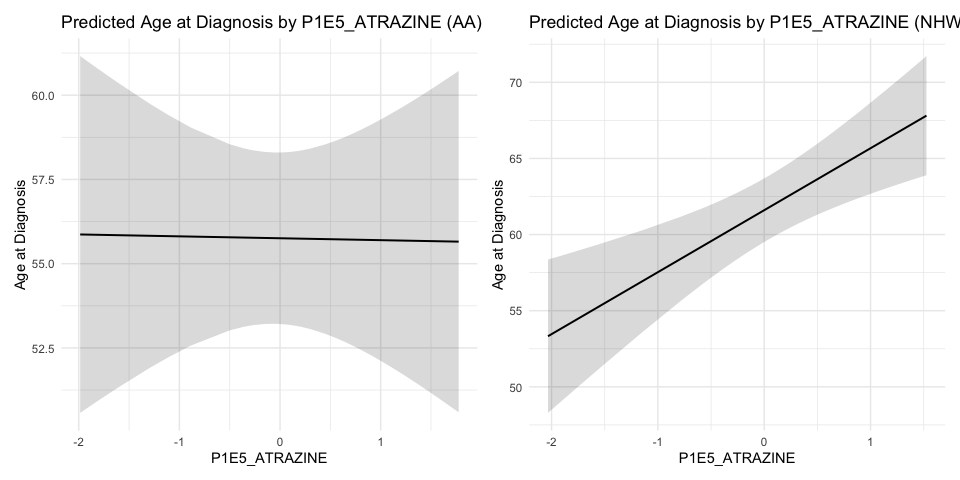

#### Age of CRC onset vs MRS using GAM - By MEEI (L:Low, MEEI <=2, H:High, MEEI >2)

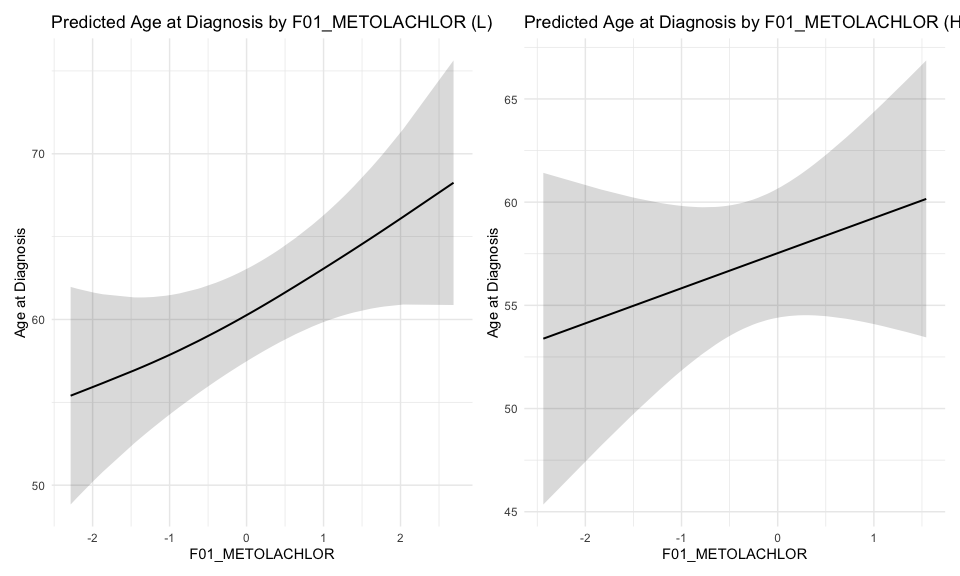

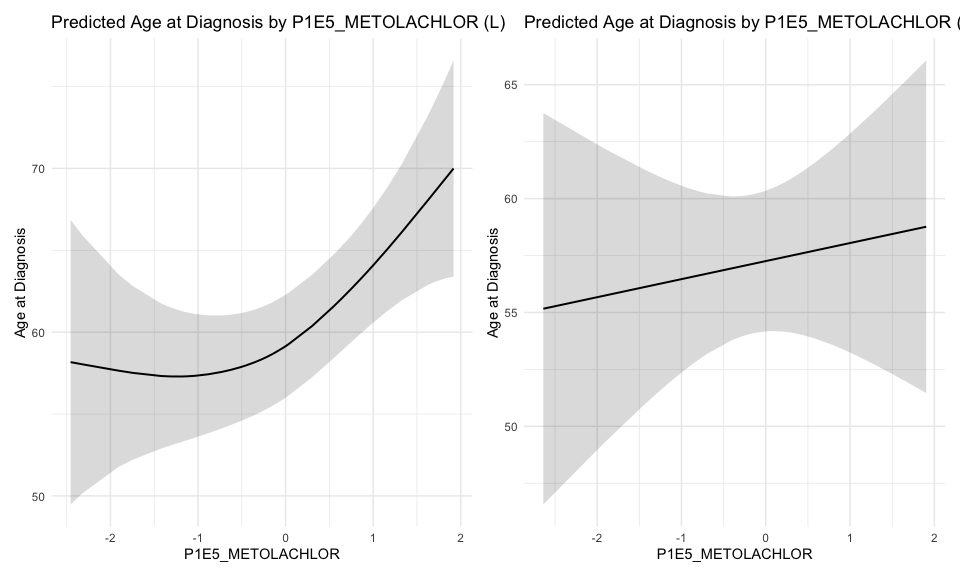

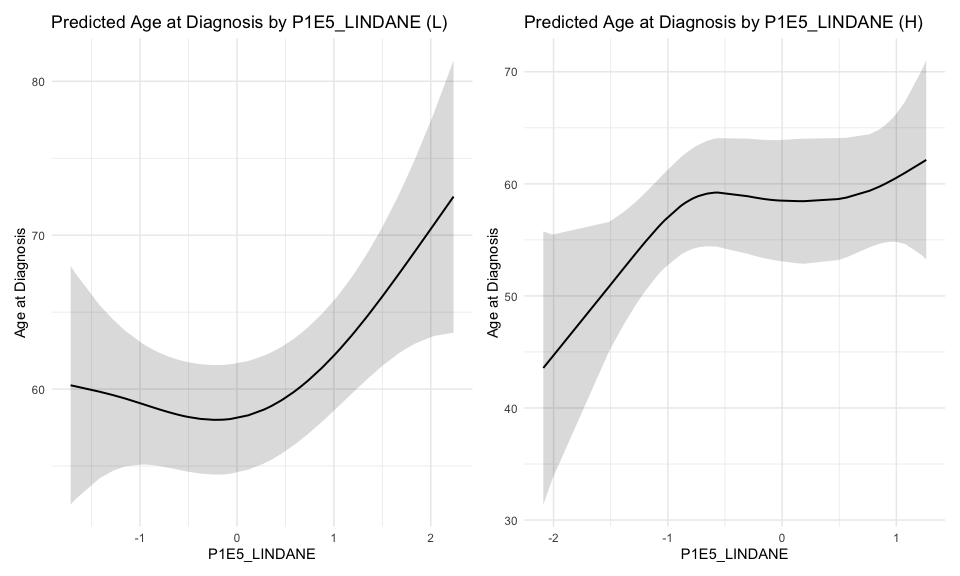

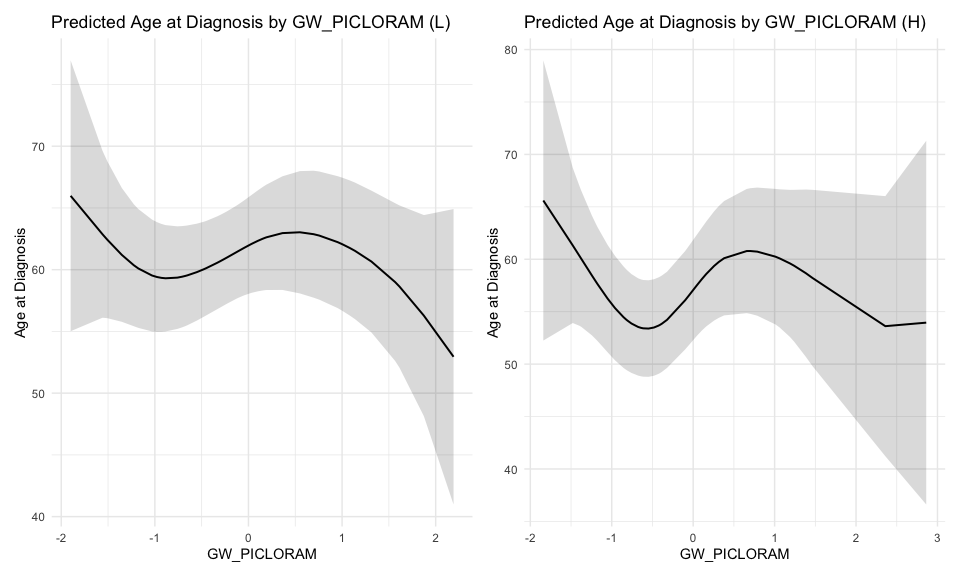

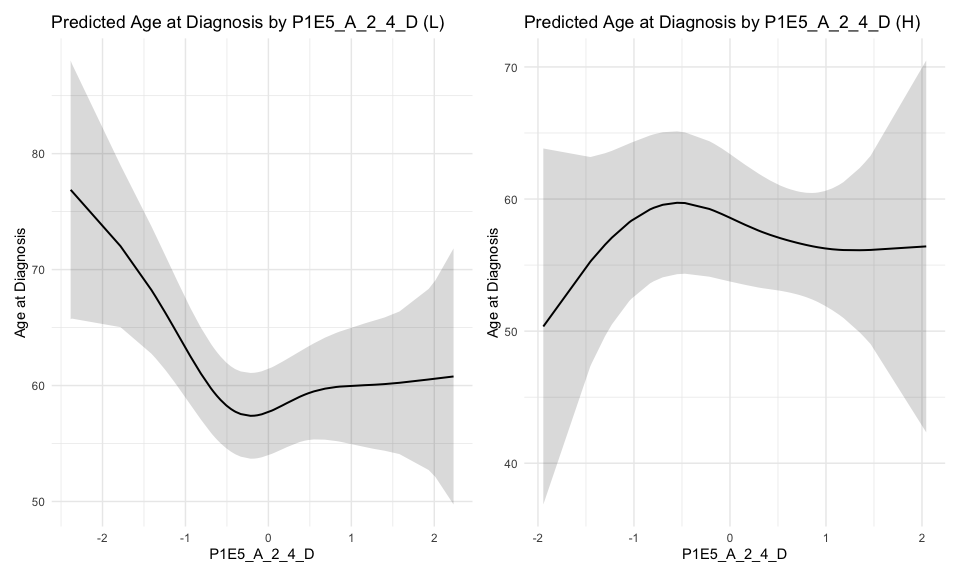

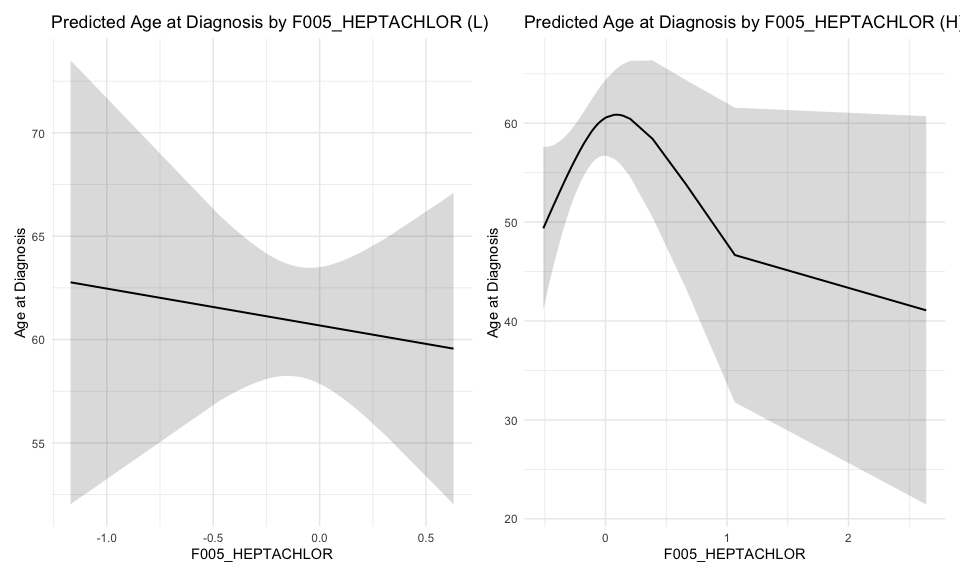

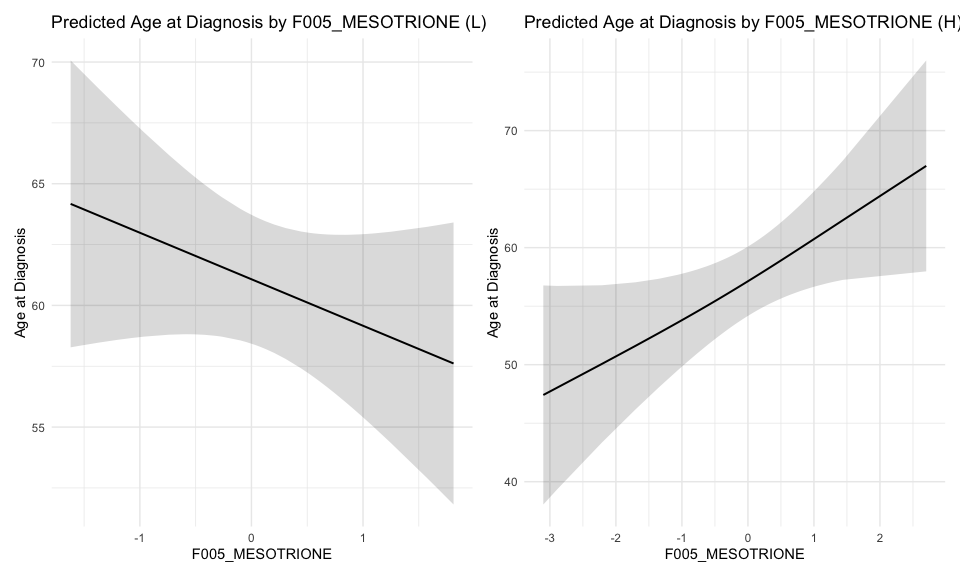

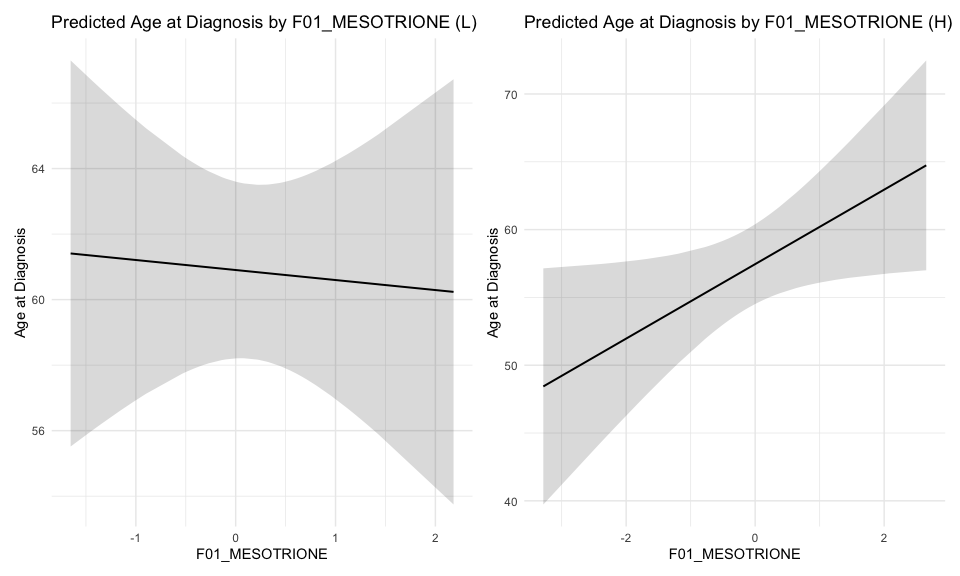

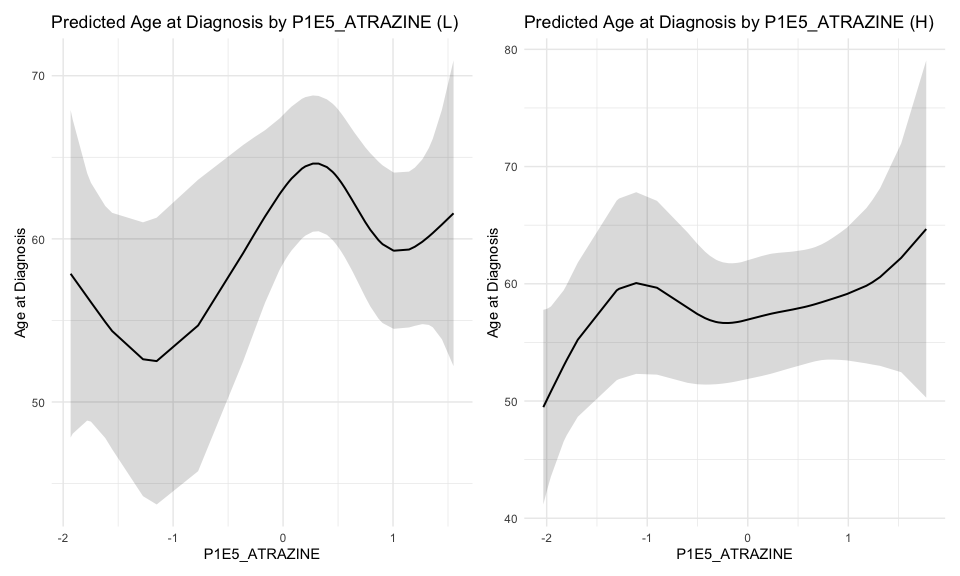

#### Age of CRC onset vs MRS using GAM - By 2015 National ADI (L:Low, ADI <=50, H:High, ADI >50)

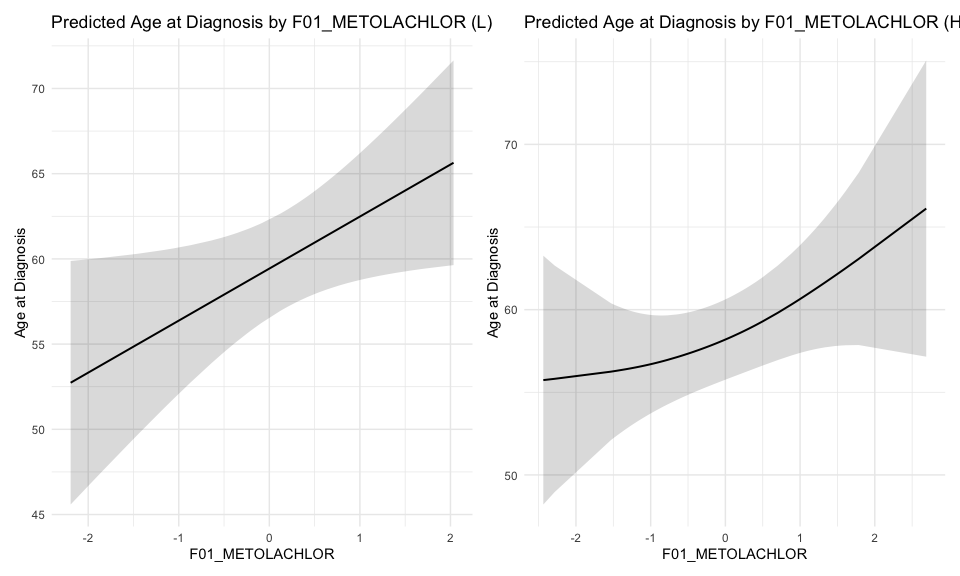

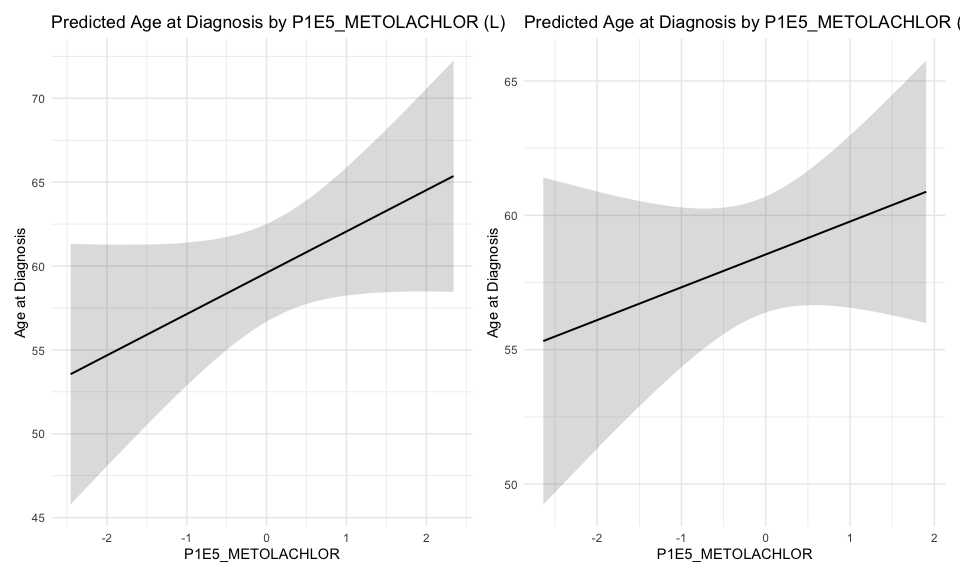

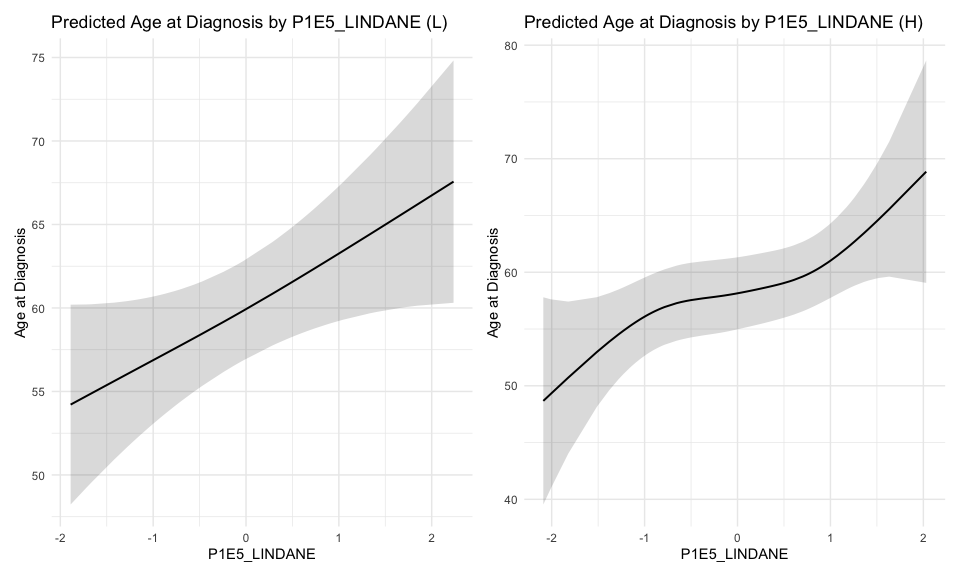

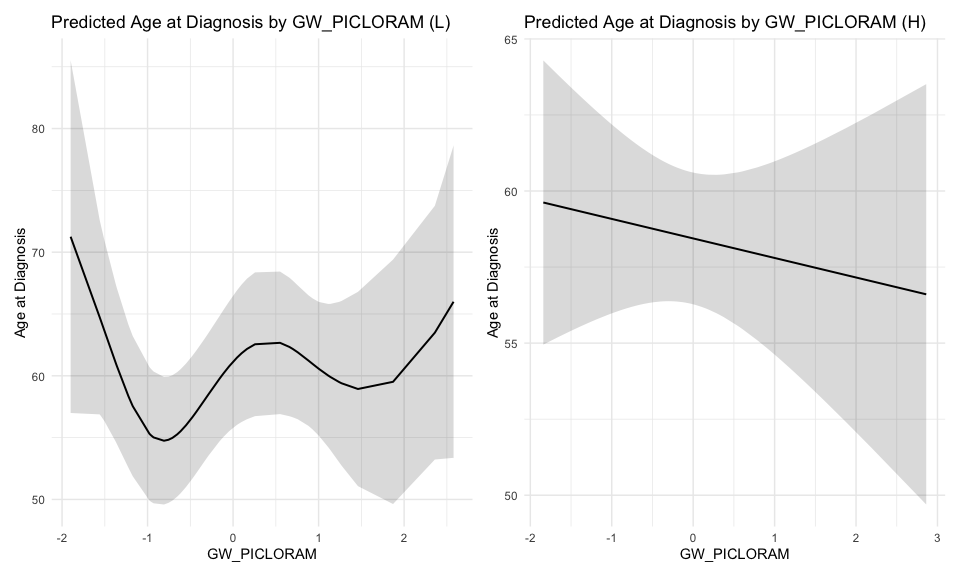

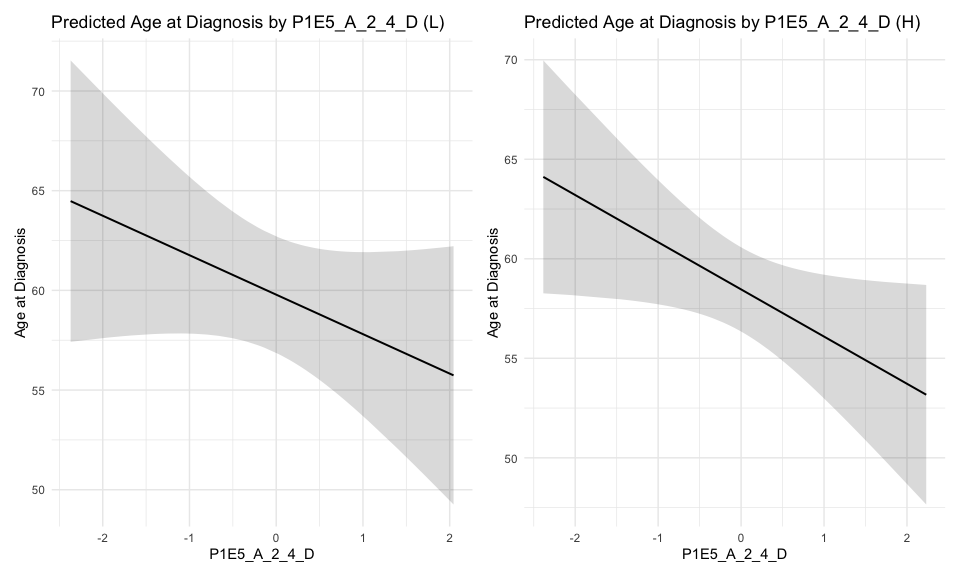

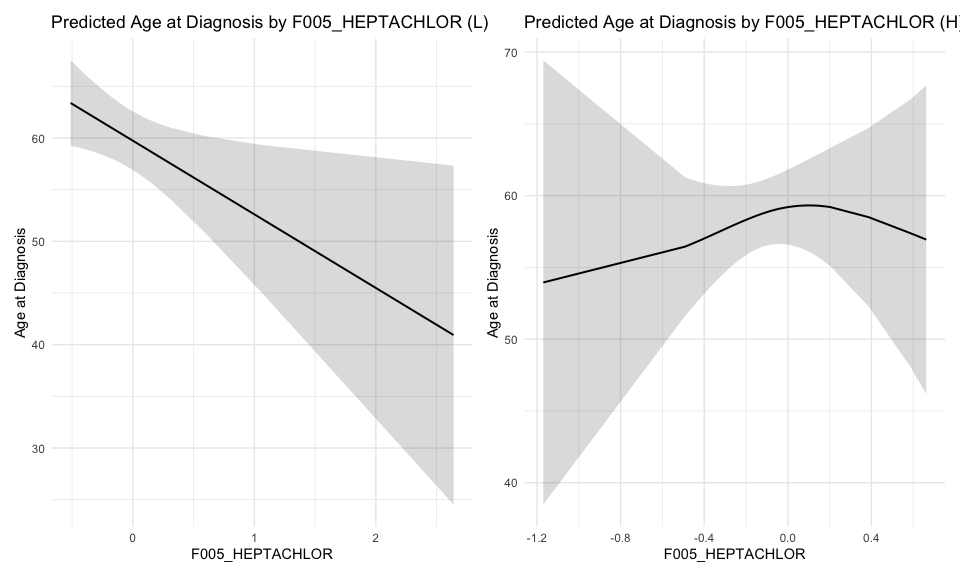

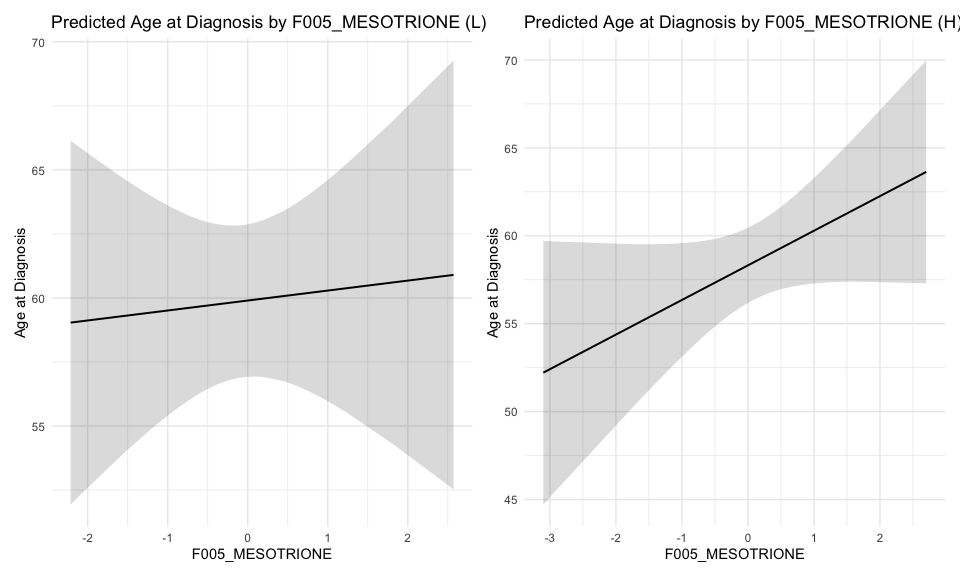

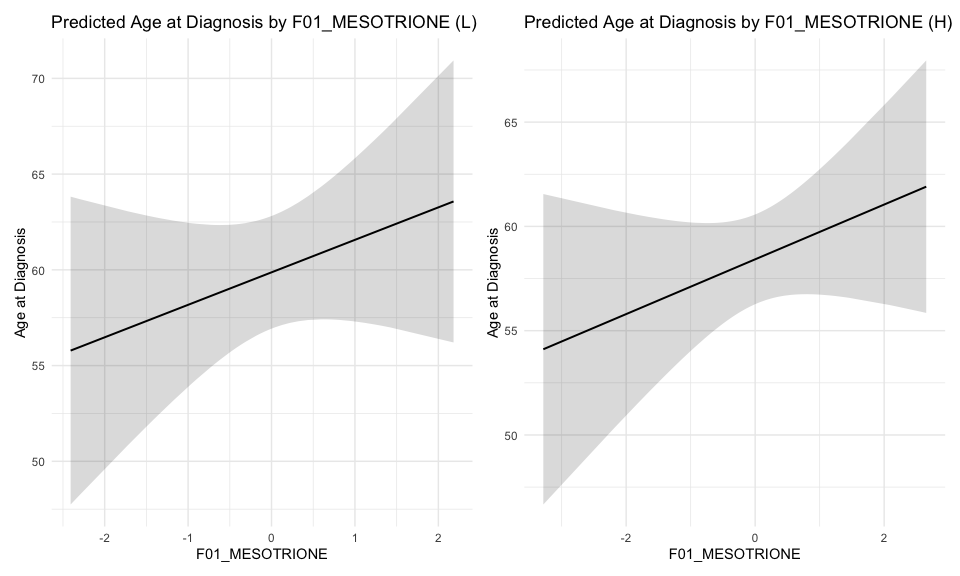

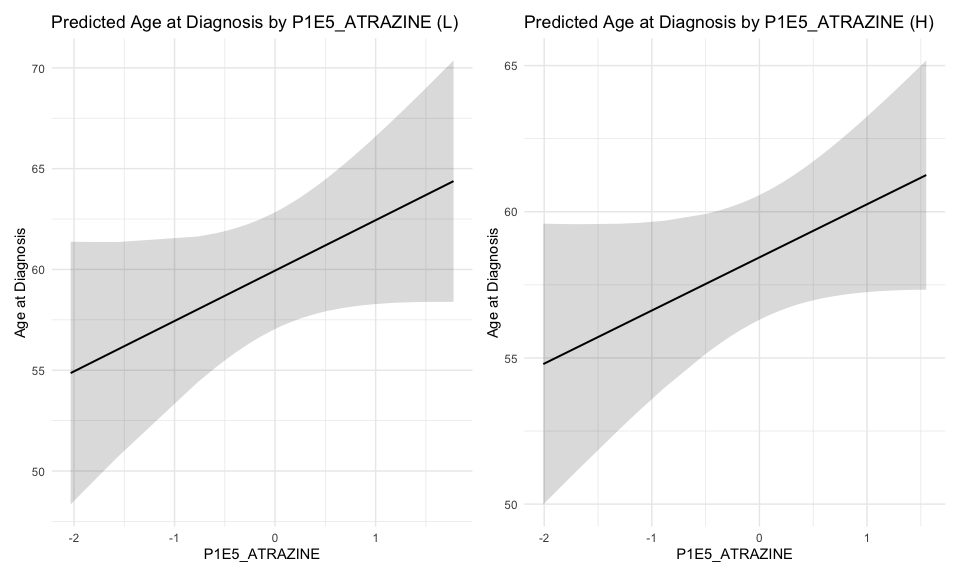

### *Pollutants*

#### Age of CRC onset vs MRS using GAM - By States (M:Detroit, Michigan, L:Louisiana)

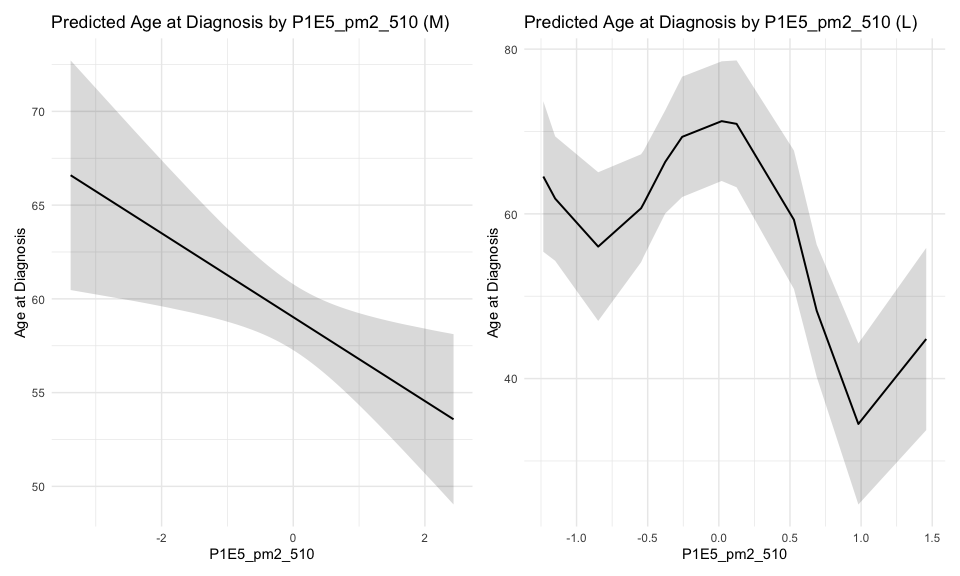

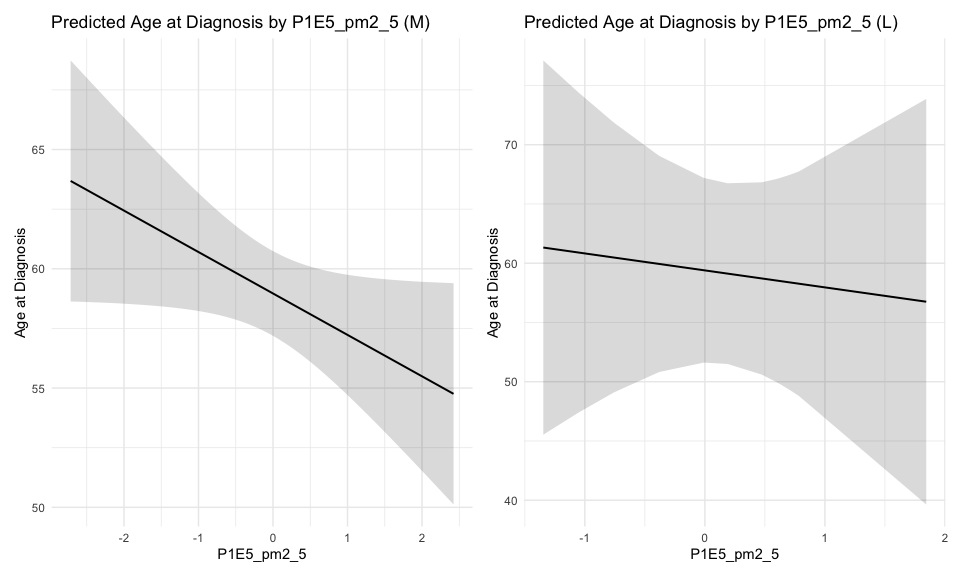

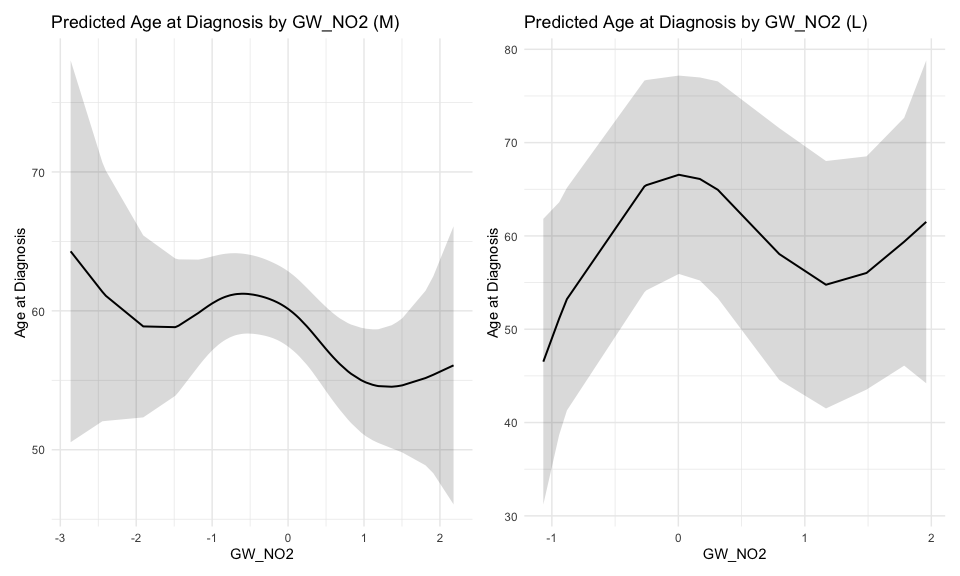

#### Age of CRC onset vs MRS using GAM - By RACE (AA:African American, NHW:NonHispanic White)

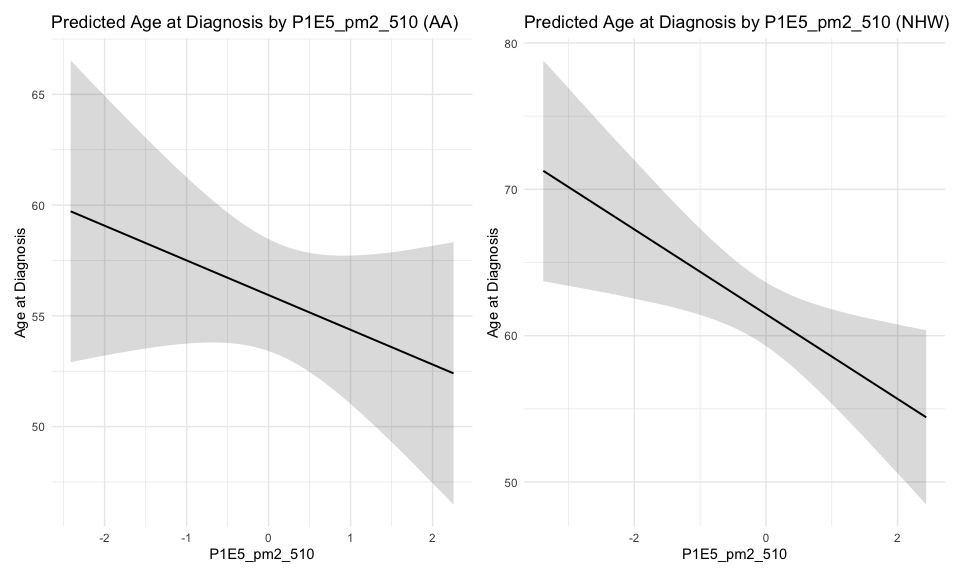

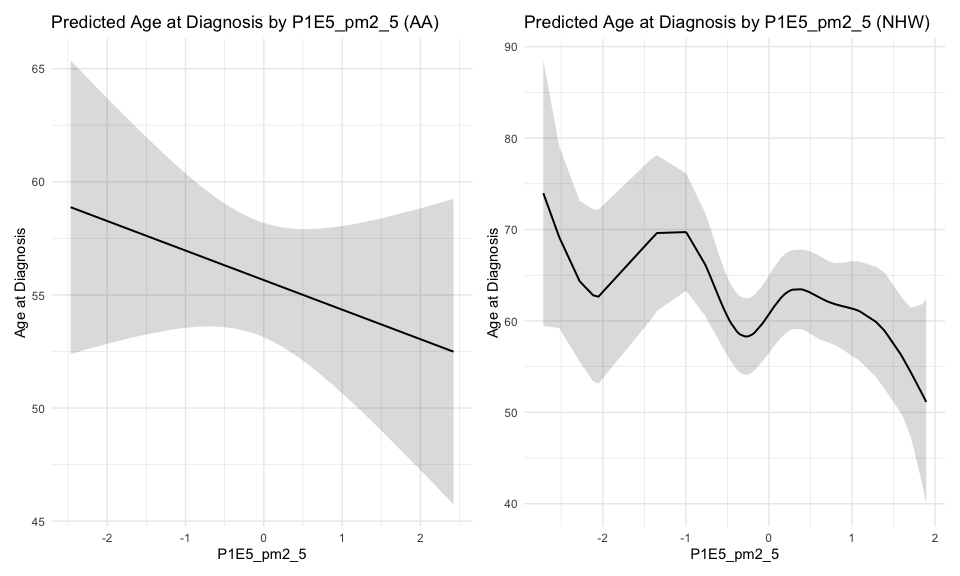

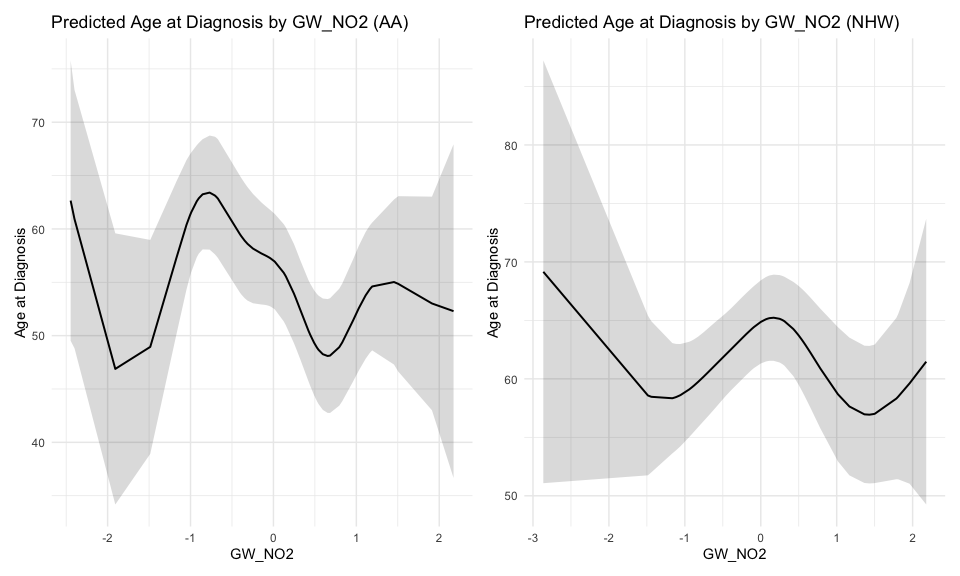

#### Age of CRC onset vs MRS using GAM - By MEEI (L:Low, MEEI <=2, H:High, MEEI >2)

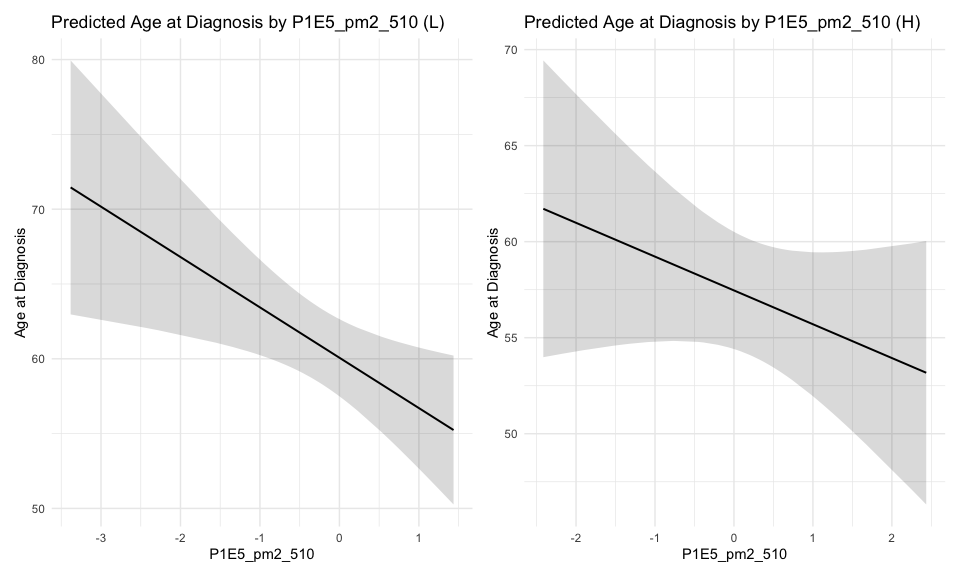

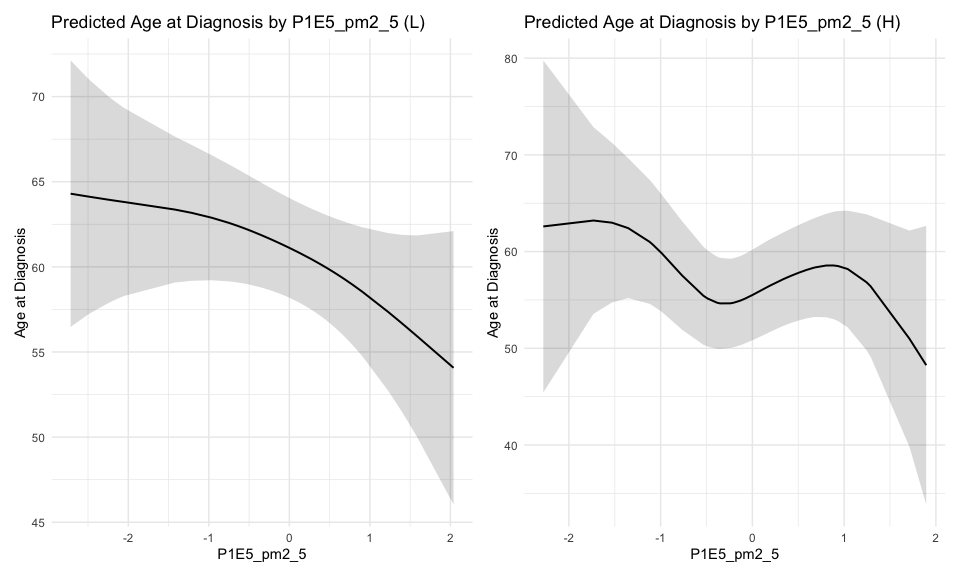

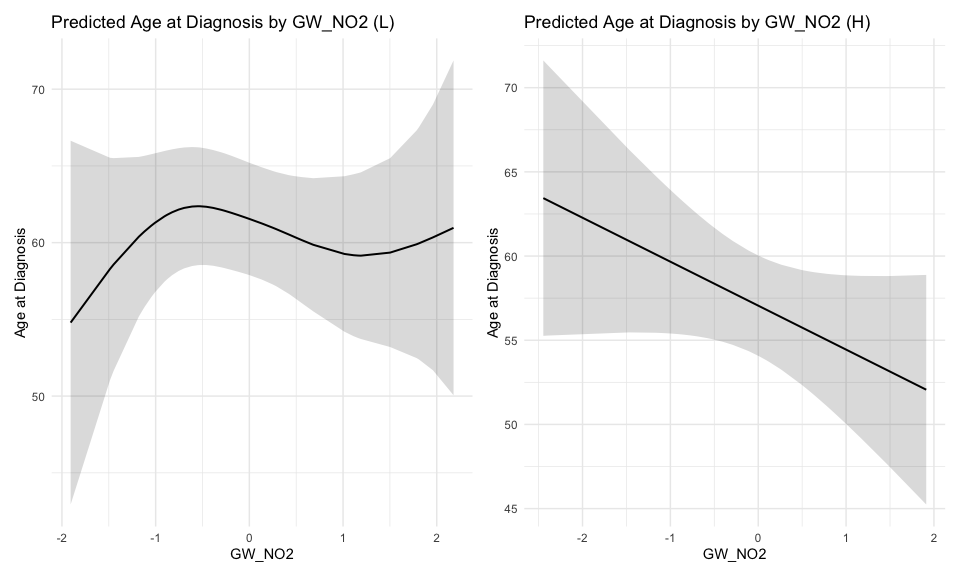

#### Age of CRC onset vs MRS using GAM - By 2015 National ADI (L:Low, ADI <=50, H:High, ADI >50)

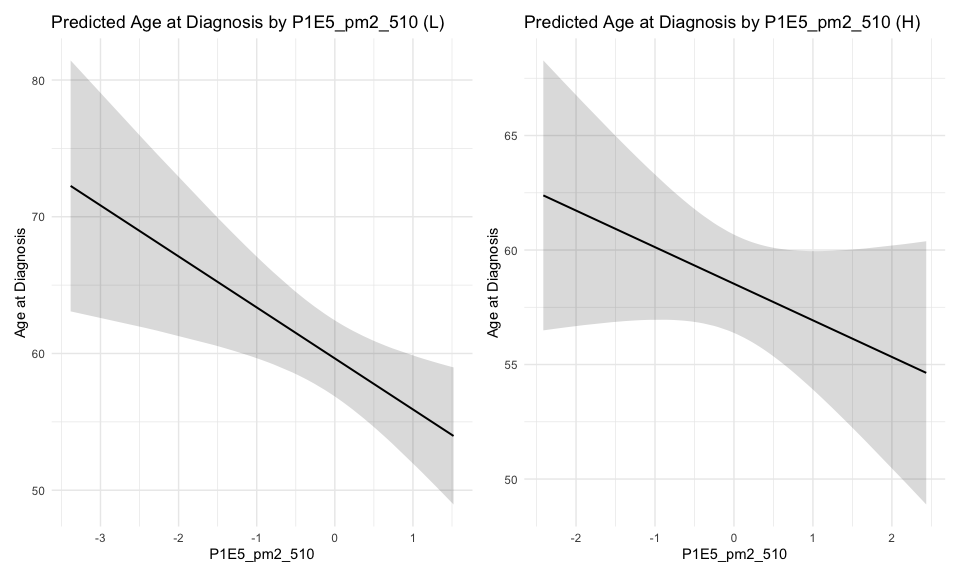

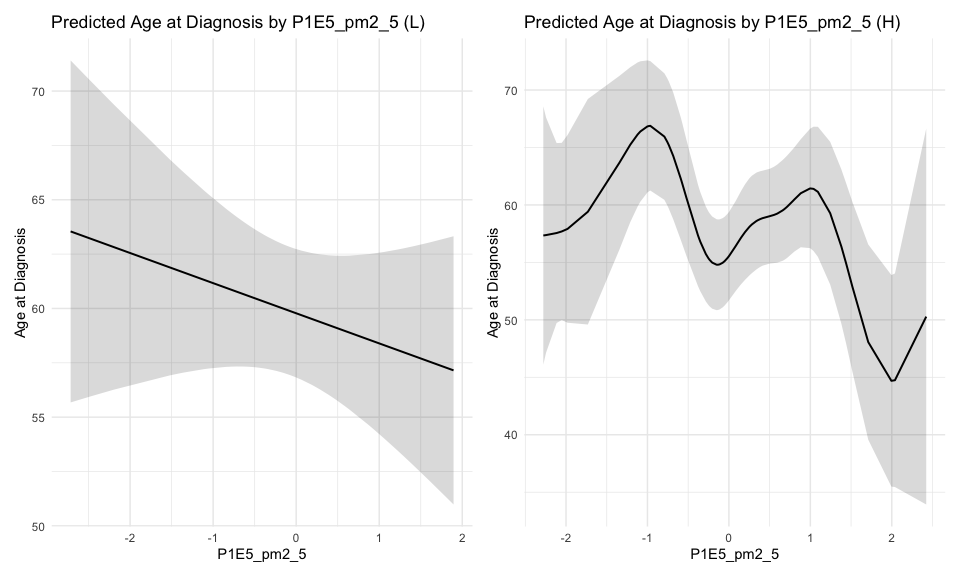

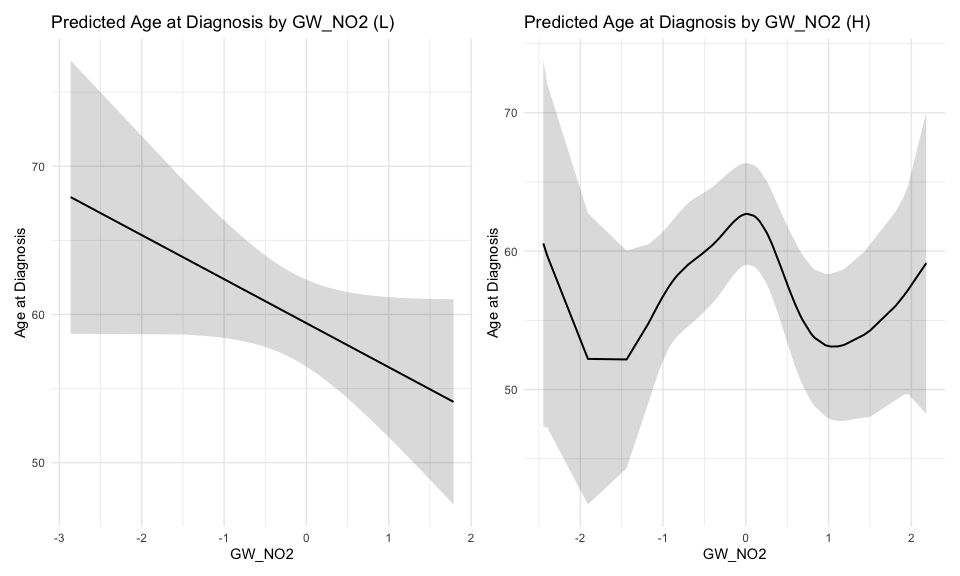

### *Lifestyle and Body Size*

#### Age of CRC onset vs MRS using GAM - By States (M:Detroit, Michigan, L:Louisiana)

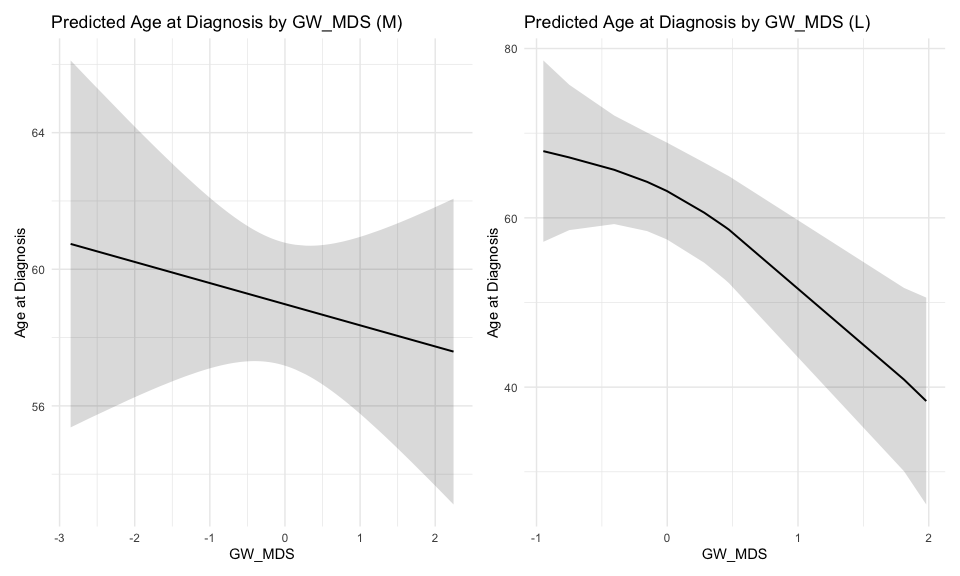

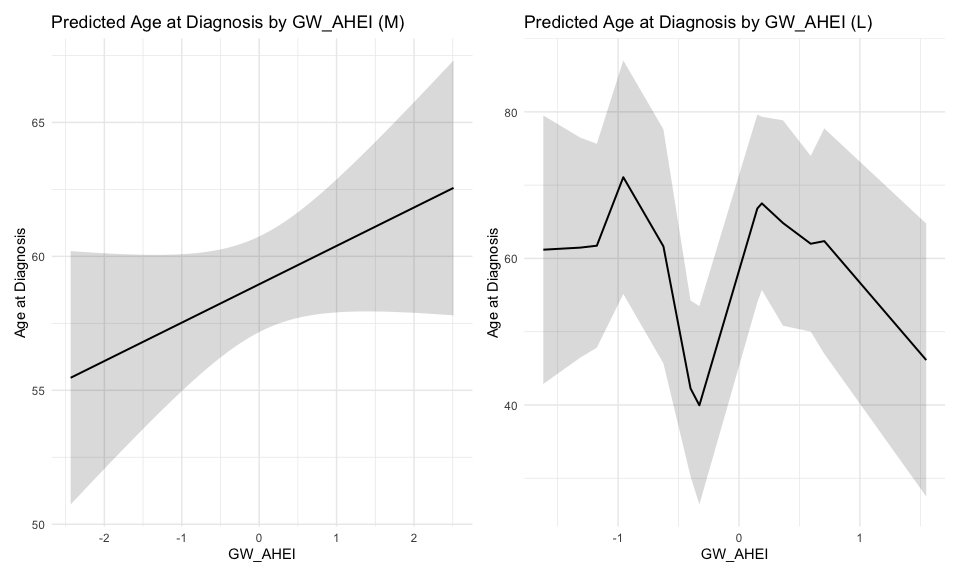

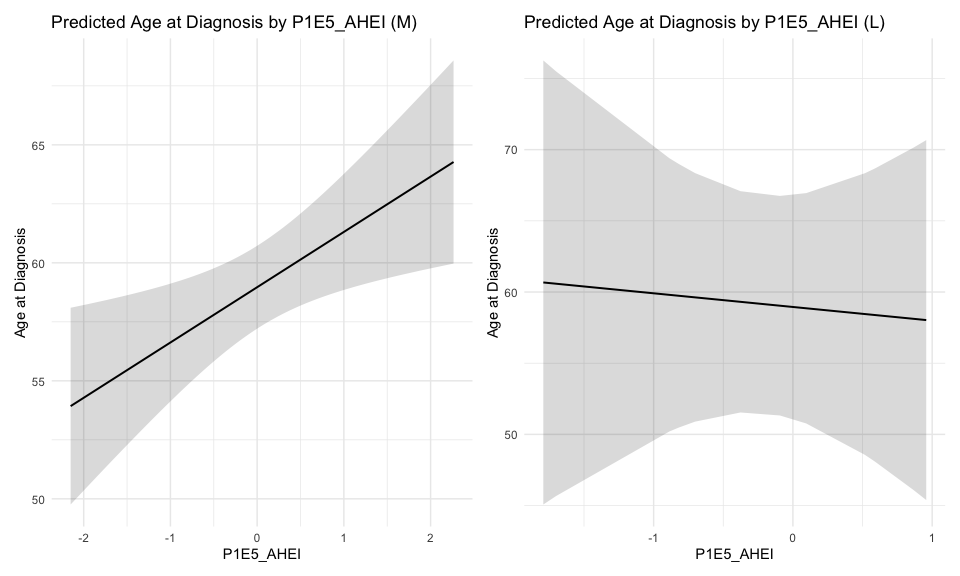

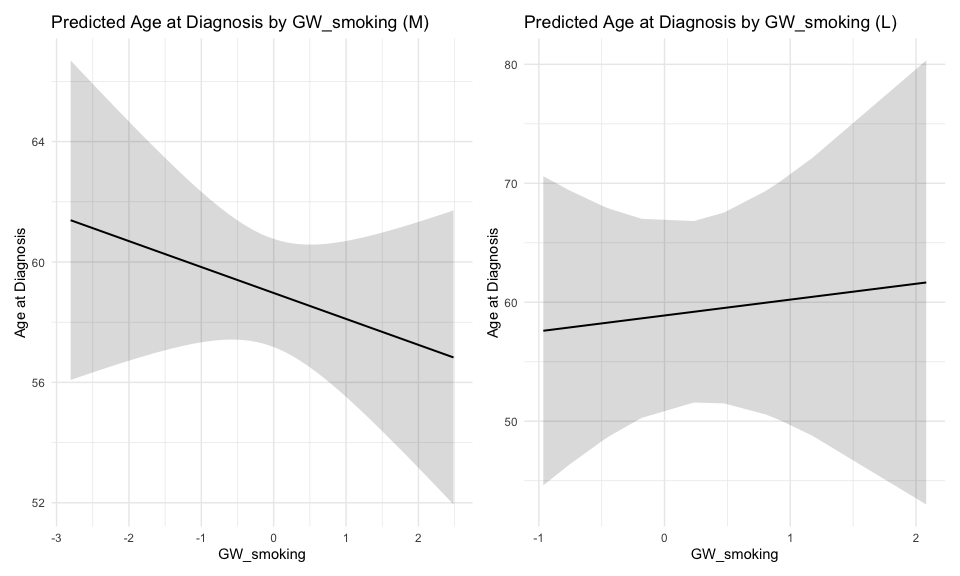

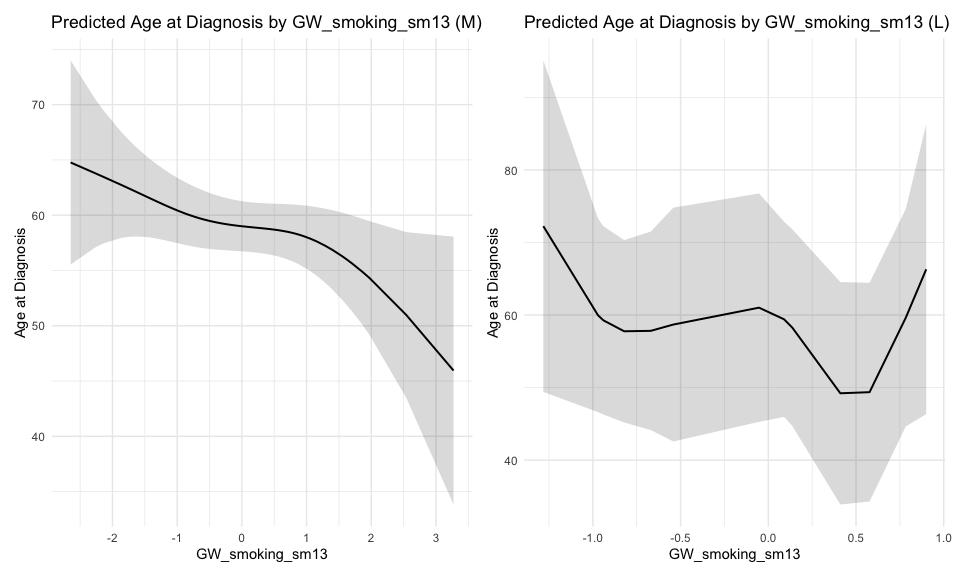

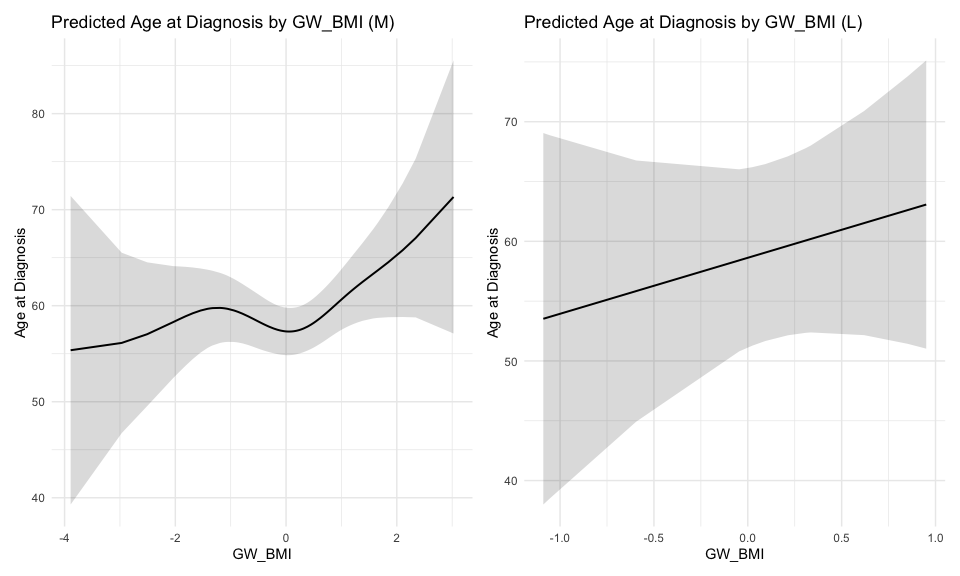

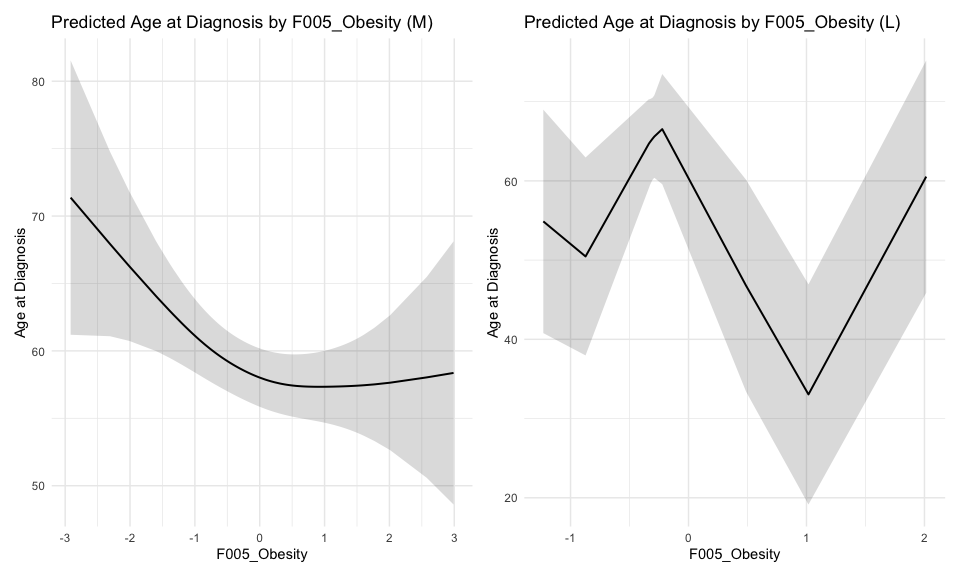

#### Age of CRC onset vs MRS using GAM - By RACE (AA:African American, NHW:NonHispanic White)

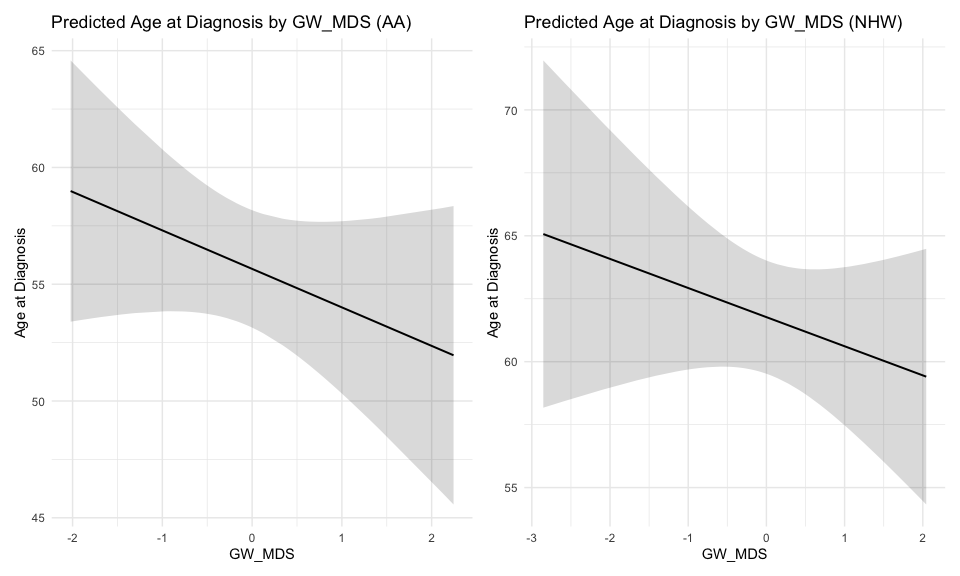

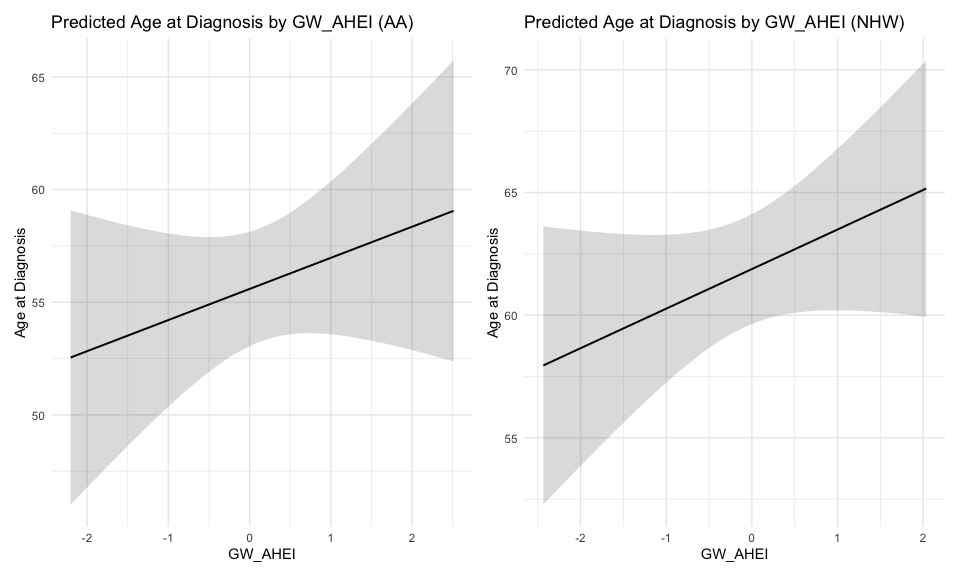

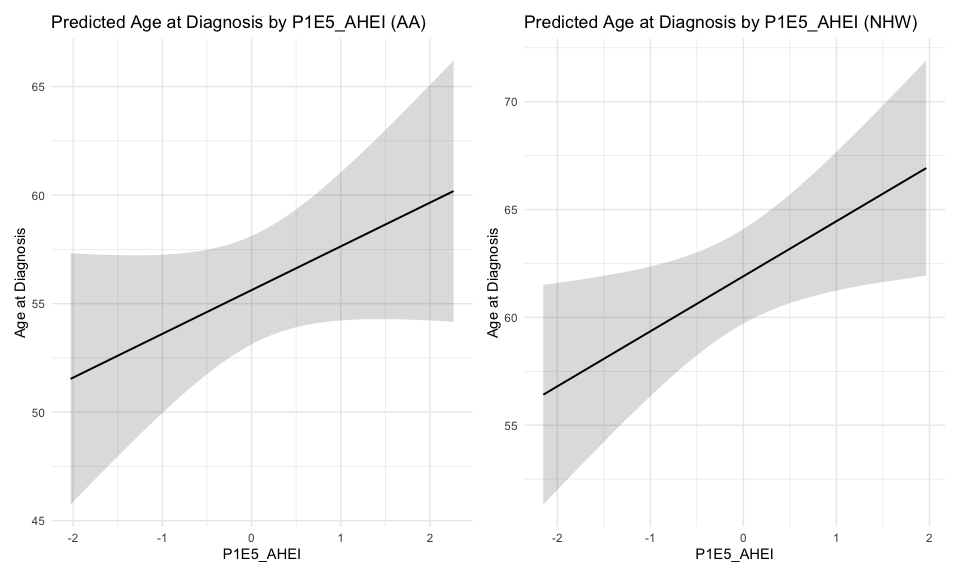

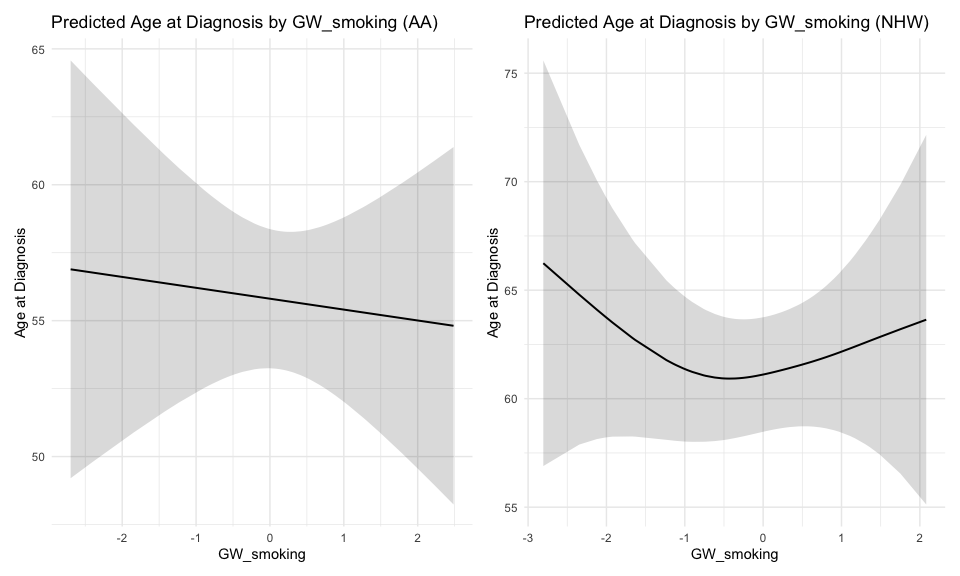

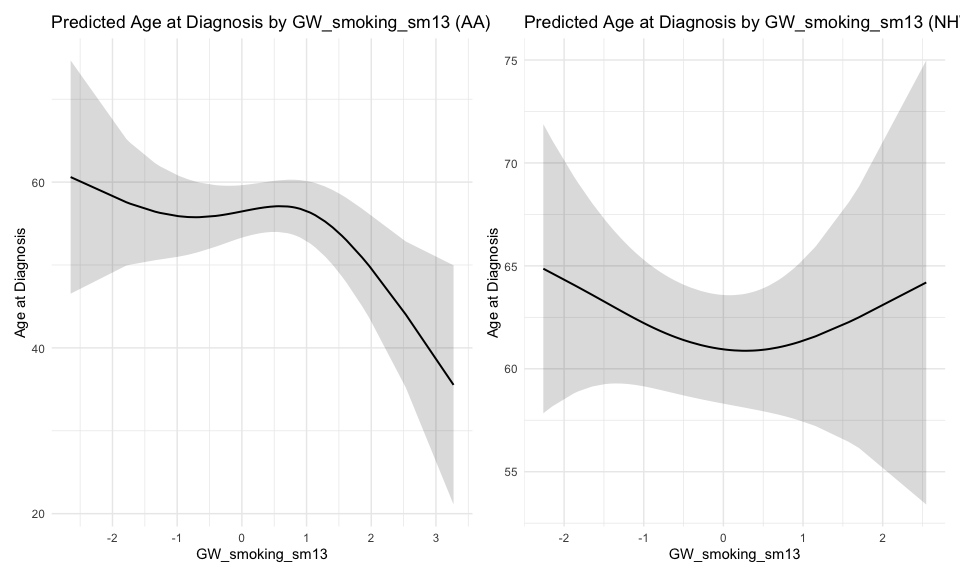

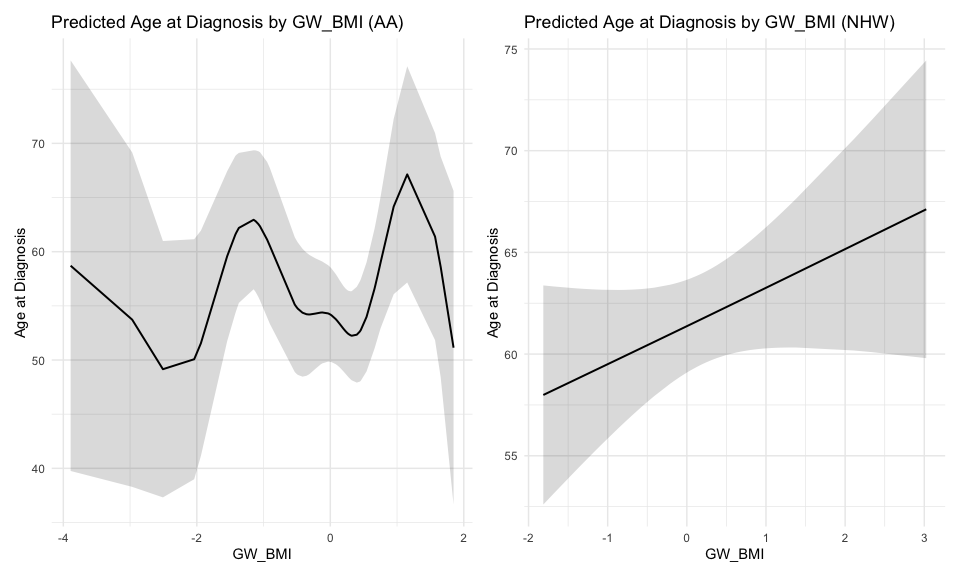

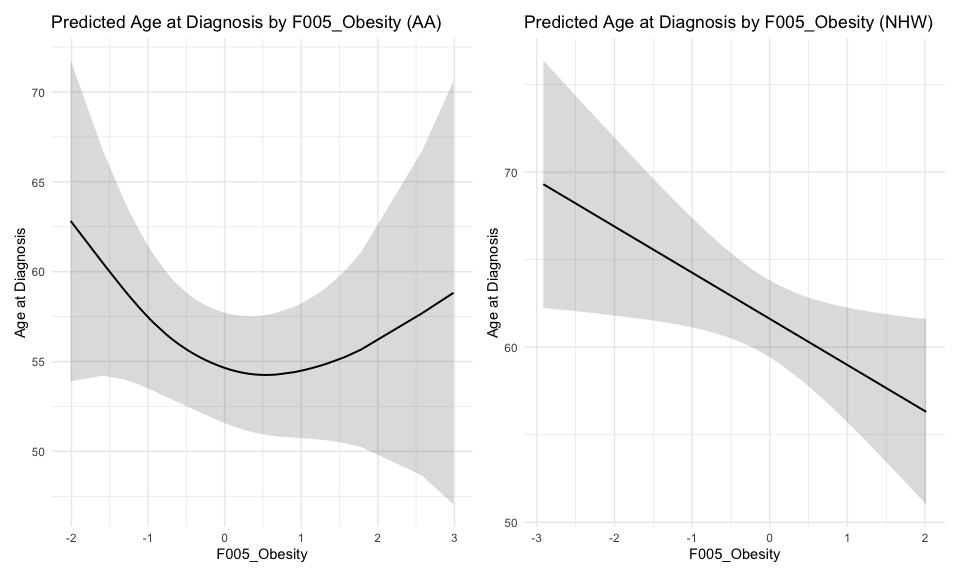

#### Age of CRC onset vs MRS using GAM - By MEEI (L:Low, MEEI <=2, H:High, MEEI >2)

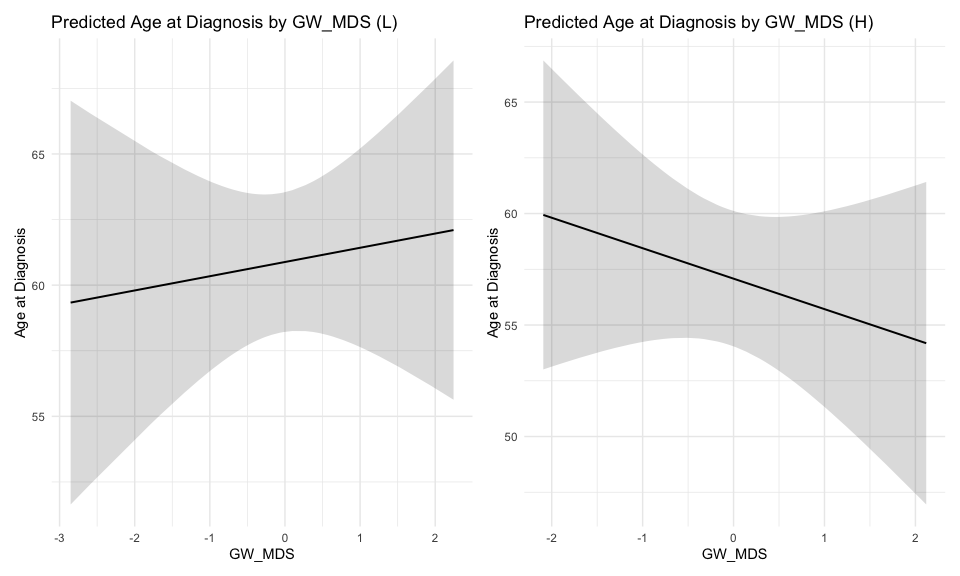

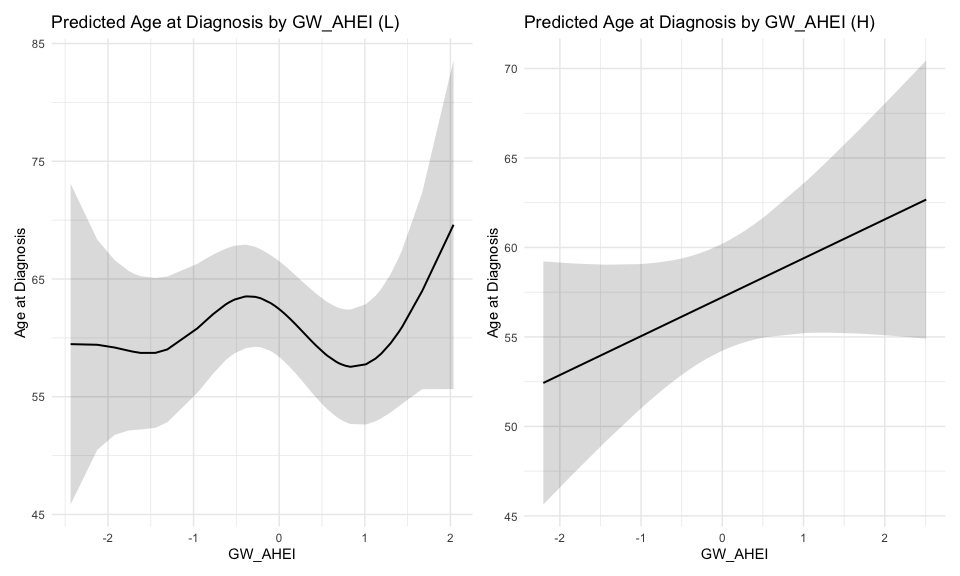

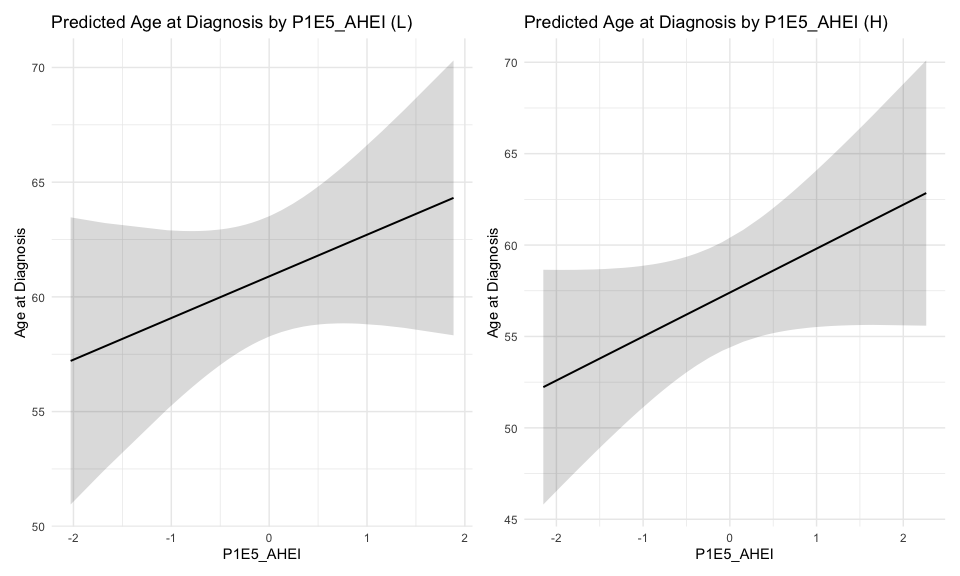

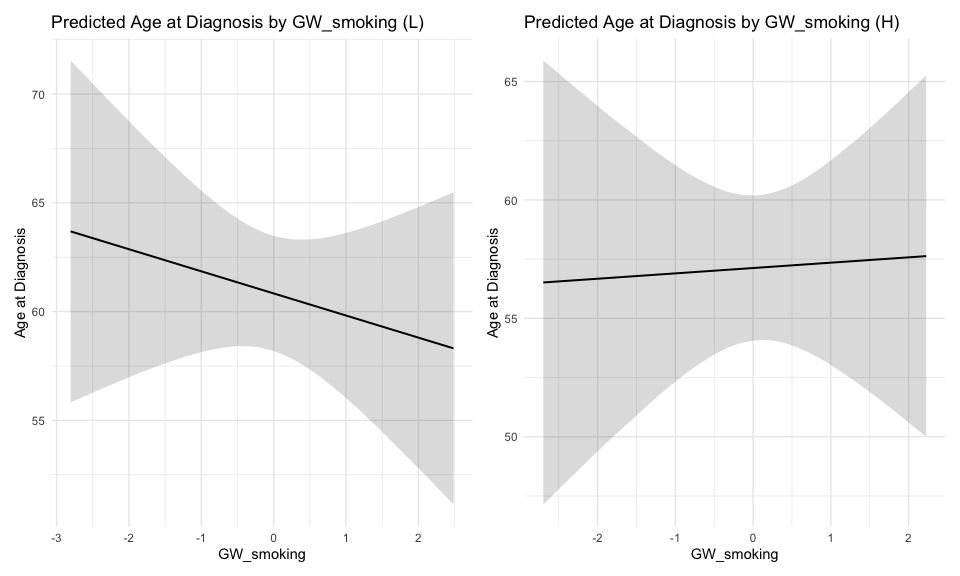

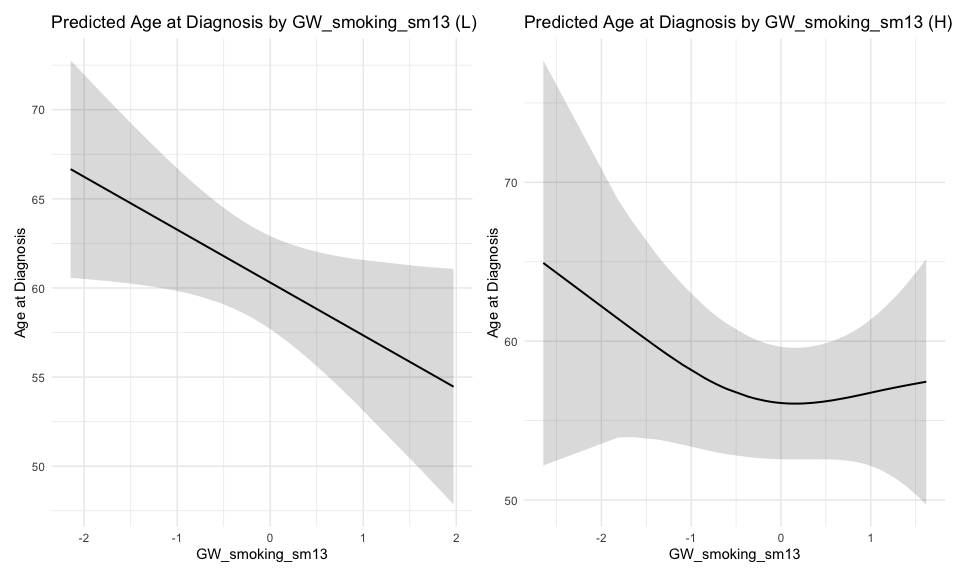

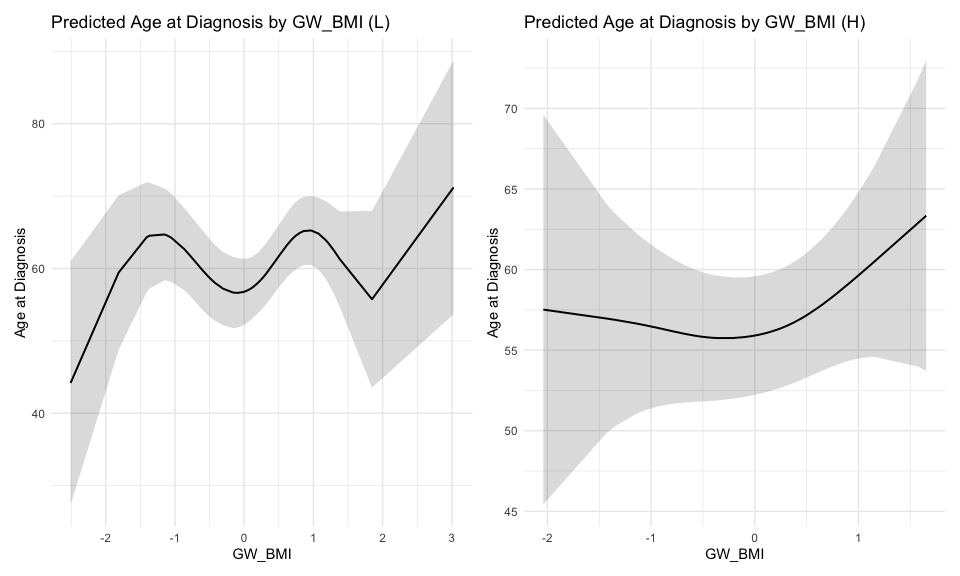

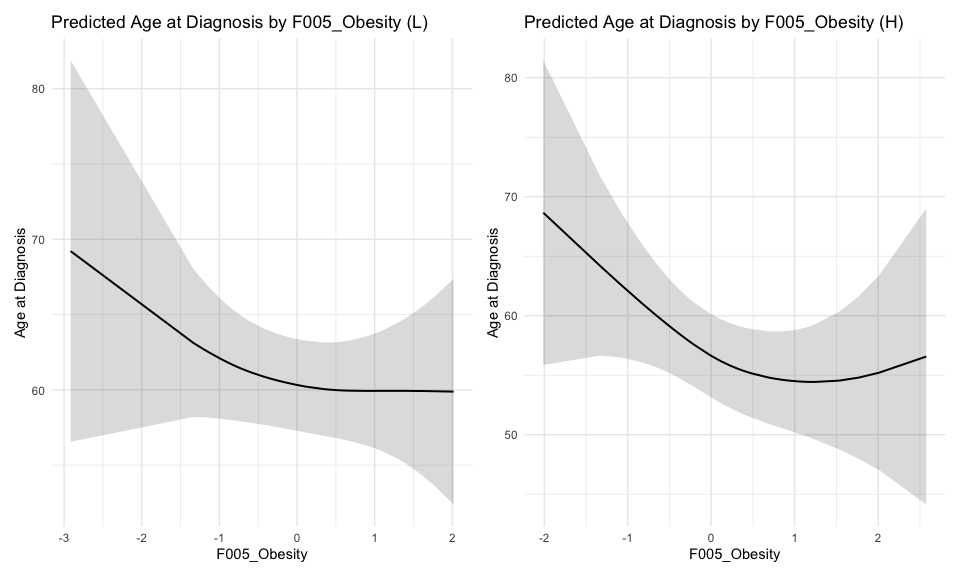

#### Age of CRC onset vs MRS using GAM - By 2015 National ADI (L:Low, ADI <=50, H:High, ADI >50)

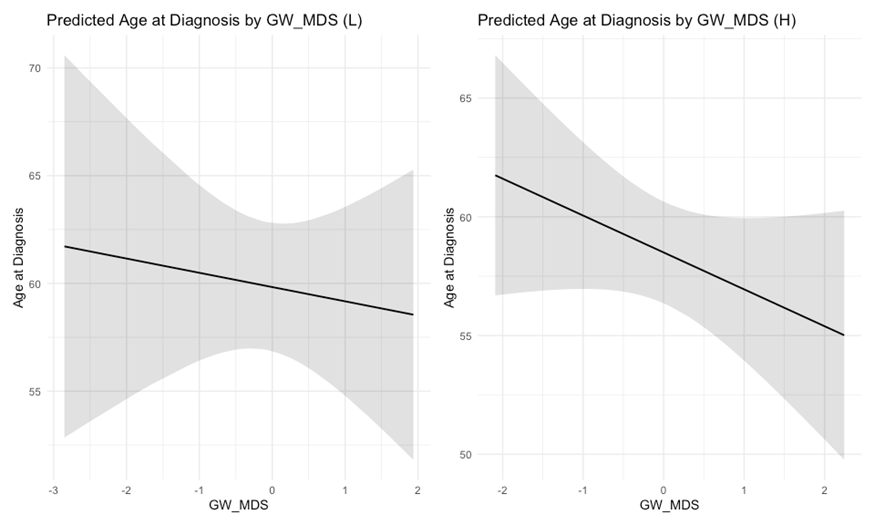

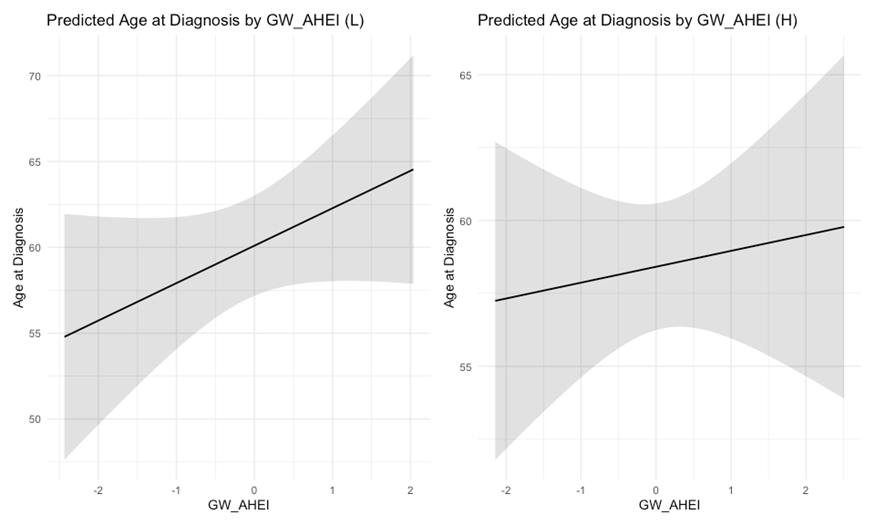

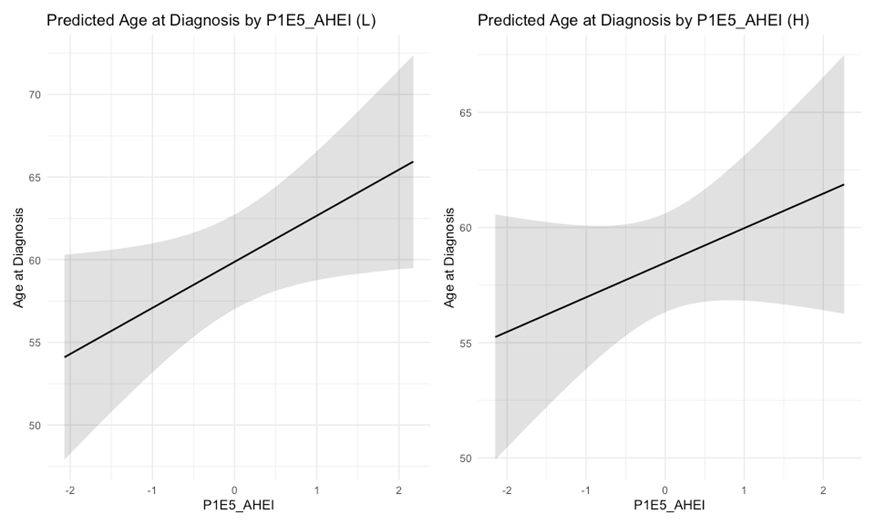

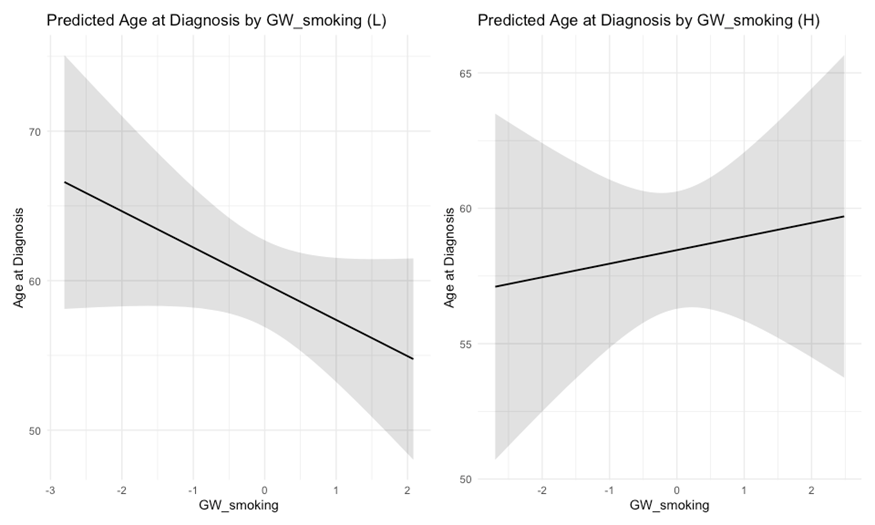

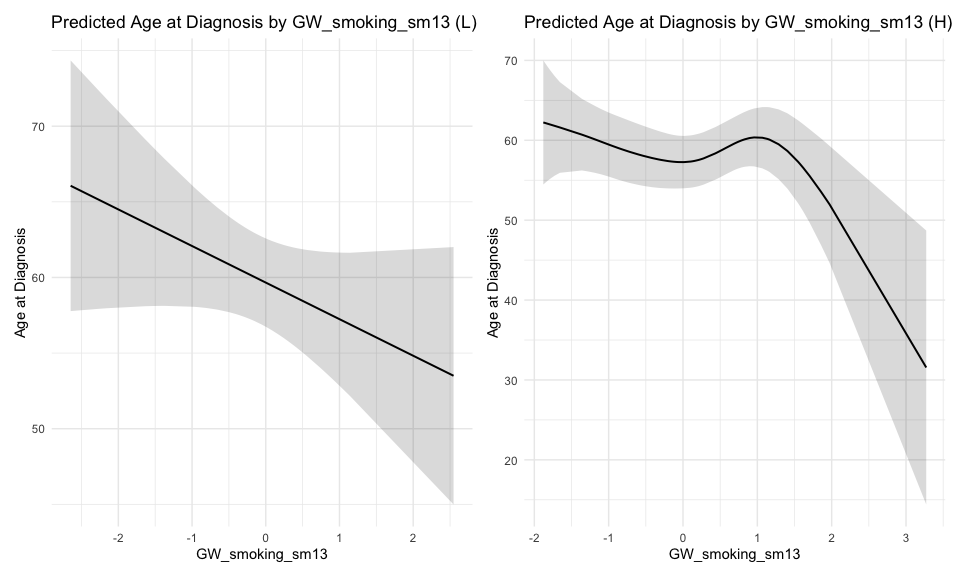

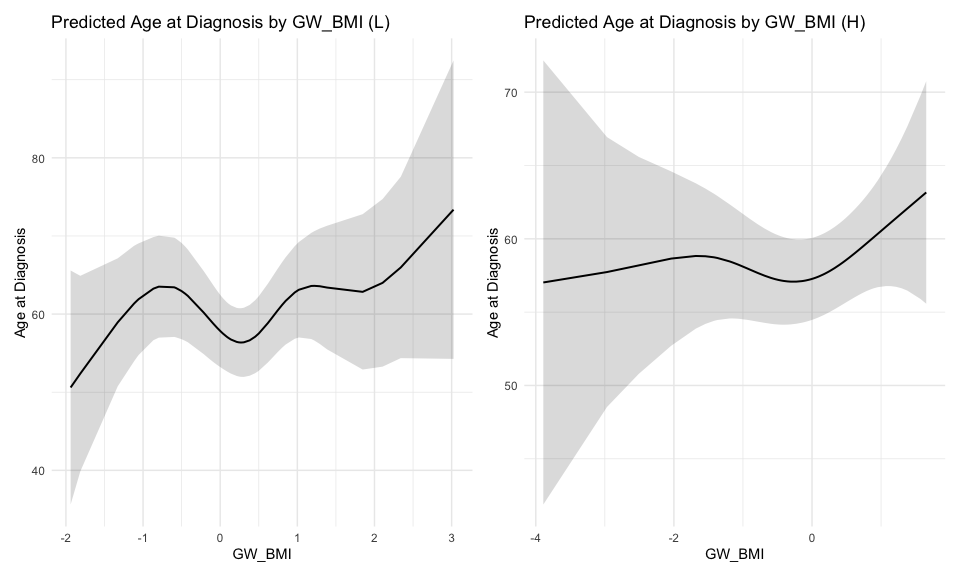

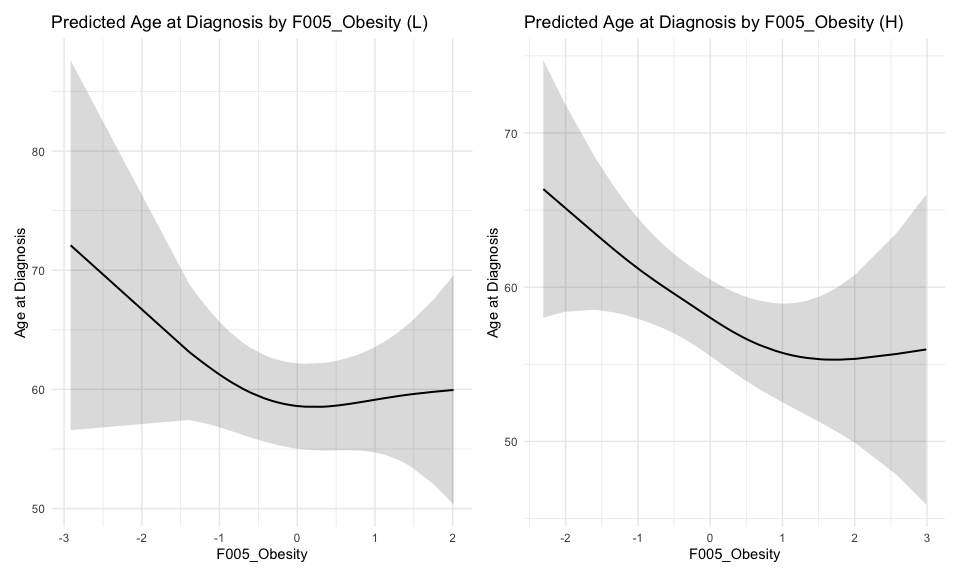
