## Supplemental Material 3 for "Epigenetic Exposure Signatures Distinguish Early-Onset from Later-Onset Colorectal Cancer in a Racially Diverse US Cohort: Findings from the Disparities and Cancer Epidemiology (DANCE) Study"

| **Supplemental Table 3a. Linear regression results adjusted for Benjamini-Hochberg FDR.** | | | | |
| --- | --- | --- | --- | --- |
| MRS (per SD-change) | Crude model | Original | Benjamini-Hochberg FDR adjusted | |
|  | $\beta$ (95% CI) | *P* | *P, 10% FDR* | *P, 20% FDR* |
| ***Pesticides*** |  |  |  |  |
| **Metolachlor (F01)** | 2.57 (0.87, 4.27) | **0.0033** | **0.011** | **0.021** |
| Metolachlor (P1E5) | 1.77 (0.05, 3.50) | **0.0442** | **0.047** | 0.095 |
| **Lindane (P1E5)** | 3.08 (1.41, 4.76) | **0.0004** | **0.005** | **0.011** |
| Picloram (GW) | 0.06 (-1.69, 1.81) | 0.9441 | 0.100 | 0.200 |
| **2,4-D herbicide (P1E5)** | -2.47 (-4.17, -0.76) | **0.0048** | **0.021** | **0.042** |
| Heptachlor (F005) | -2.13 (-3.85, -0.41) | **0.0154** | **0.026** | 0.053 |
| Mesotrione (F005) | 1.63 (-0.10, 3.36) | 0.0651 | 0.068 | 0.137 |
| Mesotrione (F01) | 1.74 (0.01, 3.47) | **0.0485** | 0.063 | 0.126 |
| Atrazine (P1E5) | 1.96 (0.23, 3.68) | **0.0262** | **0.037** | 0.074 |
| ***Pollutants*** |  |  |  |  |
| **Coarse PM2.5-10 (P1E5)** | -2.51 (-4.21, -0.80) | **0.0042** | **0.016** | **0.032** |
| Fine PM2.5 (P1E5) | -1.77 (-3.50, -0.04) | **0.0446** | 0.053 | 0.105 |
| NO_2_ (GW) | -2.03 (-3.75, -0.31) | **0.0212** | **0.032** | 0.063 |
| ***Lifestyle / body composition*** | | | |  |
| MDS (GW) | -1.45 (-3.19, 0.29) | 0.1008 | 0.084 | 0.168 |
| AHEI (GW) | 1.31 (-0.42, 3.05) | 0.1373 | 0.089 | 0.179 |
| AHEI (P1E5) | 2.00 (0.23, 3.68) | **0.0264** | **0.042** | 0.084 |
| Smoking (GW) | -0.98 (-2.73, 0.76) | 0.2672 | 0.095 | 0.189 |
| Smoking-Maas (GW) | -1.76 (-3.49, -0.03) | **0.0462** | 0.058 | 0.116 |
| BMI (GW) | 1.51 (-0.22, 3.25) | 0.0871 | 0.074 | 0.147 |
| Obesity (F005) | -1.49 (-3.22, 0.25) | 0.0923 | 0.079 | 0.158 |
| *MRS thresholds: GW = genome wide, F005 = 5% false discovery rate, F01 = 10% false discovery rate, P1E5 = p-value<10^-5^.* | | | | |

| **Supplemental Table 3b. Polytomous regression results adjusted for Benjamini-Hochberg FDR.** | | | | | |
| --- | --- | --- | --- | --- | --- |
| MRS (per SD-change) | Outcome group | Crude model | Original | Benjamini-Hochberg FDR adjusted | |
|  |  | OR (95% CI) | *P* | *P* at 10% FDR | *P* at 20% FDR |
| ***Pesticides*** |  |  |  |  |  |
| 2,4-D herbicide (P1E5) | EOCRC | **1.67 (1.03, 2.72)** | **0.0378** | **0.026** | 0.053 |
| 2,4-D herbicide (P1E5) | IOCRC | **1.53 (1.04, 2.25)** | **0.0314** | **0.018** | **0.037** |
| Atrazine (P1E5) | EOCRC | 0.66 (0.41, 1.06) | 0.0863 | **0.037** | 0.074 |
| Atrazine (P1E5) | IOCRC | 0.73 (0.49, 1.08) | 0.1130 | **0.042** | 0.084 |
| Heptachlor (F005) | EOCRC | 5.19 (1.00, 26.9) | 0.0501 | **0.029** | 0.058 |
| Heptachlor (F005) | IOCRC | 1.78 (0.40, 7.93) | 0.4481 | 0.087 | 0.174 |
| Lindane (P1E5) | EOCRC | **0.45 (0.27, 0.75)** | **0.0022** | **0.003** | **0.005** |
| Lindane (P1E5) | IOCRC | 0.92 (0.63, 1.35) | 0.6662 | 0.095 | 0.189 |
| Mesotrione (F005) | EOCRC | 0.71 (0.44, 1.13) | 0.1463 | 0.053 | 0.105 |
| Mesotrione (F005) | IOCRC | 0.80 (0.54, 1.17) | 0.2496 | 0.071 | 0.142 |
| Mesotrione (F01) | EOCRC | 0.71 (0.44, 1.14) | 0.1562 | 0.058 | 0.116 |
| Mesotrione (F01) | IOCRC | 0.82 (0.56, 1.19) | 0.2962 | 0.074 | 0.147 |
| Metolachlor (F01) | EOCRC | **0.53 (0.33, 0.87)** | **0.0125** | **0.011** | **0.021** |
| Metolachlor (F01) | IOCRC | **0.62 (0.42, 0.92)** | **0.0172** | **0.013** | **0.026** |
| Metolachlor (P1E5) | EOCRC | 0.67 (0.42, 1.09) | 0.1072 | **0.039** | 0.079 |
| Metolachlor (P1E5) | IOCRC | **0.63 (0.42, 0.93)** | **0.0216** | **0.016** | **0.032** |
| Picloram (GW) | EOCRC | 1.10 (0.70, 1.73) | 0.6763 | 0.097 | 0.195 |
| Picloram (GW) | IOCRC | 0.84 (0.57, 1.22) | 0.3470 | 0.079 | 0.158 |
| ***Pollutants*** |  |  |  |  |  |
| Coarse PM2.5-10 (P1E5) | EOCRC | **1.94 (1.16, 3.24)** | **0.0111** | **0.008** | **0.016** |
| Coarse PM2.5-10 (P1E5) | IOCRC | 1.29 (0.89, 1.88) | 0.1801 | 0.061 | 0.121 |
| Fine PM2.5 (P1E5) | EOCRC | 1.42 (0.88, 2.30) | 0.1535 | 0.055 | 0.111 |
| Fine PM2.5 (P1E5) | IOCRC | 1.12 (0.77, 1.62) | 0.5524 | 0.092 | 0.184 |
| NO_2_ (GW) | EOCRC | 1.57 (0.97, 2.55) | 0.0657 | **0.032** | 0.063 |
| NO_2_ (GW) | IOCRC | 1.37 (0.92, 2.03) | 0.1177 | **0.045** | 0.089 |
| ***Lifestyle / body composition*** | | | | | |
| AHEI (GW) | EOCRC | 0.82 (0.51, 1.31) | 0.409 | 0.084 | 0.168 |
| AHEI (GW) | IOCRC | 0.74 (0.51, 1.09) | 0.1252 | **0.047** | 0.095 |
| AHEI (P1E5) | EOCRC | 0.73 (0.45, 1.17) | 0.1895 | 0.063 | 0.126 |
| AHEI (P1E5) | IOCRC | 0.71 (0.49, 1.04) | 0.0797 | **0.034** | 0.068 |
| BMI (GW) | EOCRC | 0.70 (0.44, 1.12) | 0.1381 | 0.050 | 0.100 |
| BMI (GW) | IOCRC | 0.80 (0.54, 1.17) | 0.2484 | 0.068 | 0.137 |
| MDS (GW) | EOCRC | **1.73 (1.04, 2.86)** | **0.035** | **0.024** | **0.047** |
| MDS (GW) | IOCRC | 0.85 (0.58, 1.24) | 0.405 | 0.082 | 0.163 |
| Obesity (F005) | EOCRC | 1.37 (0.84, 2.24) | 0.208 | 0.066 | 0.132 |
| Obesity (F005) | IOCRC | **1.84 (1.22, 2.78)** | **0.0037** | **0.005** | **0.011** |
| Smoking (GW) | EOCRC | 1.16 (0.73, 1.85) | 0.5253 | 0.089 | 0.179 |
| Smoking (GW) | IOCRC | 1.20 (0.83, 1.74) | 0.3411 | 0.076 | 0.153 |
| Smoking-Maas (GW) | EOCRC | **1.71 (1.05, 2.79)** | **0.0328** | **0.021** | **0.042** |
| Smoking-Maas (GW) | IOCRC | 1.00 (0.68, 1.46) | 0.9895 | 0.100 | 0.200 |
| *MRS thresholds: GW = genome wide, F005 = 5% false discovery rate, F01 = 10% false discovery rate, P1E5 = p-value<10^-5^.* | | | | | |
